# ALADYNOULLI: A Bayesian approach to disease progression modeling for genomic discovery and clinical prediction

**DOI:** 10.1101/2024.09.29.24314557

**Authors:** Sarah M. Urbut, Yi Ding, Tetsushi Nakao, Satoshi Koyama, Xilin Jiang, Achyutha Harish, Leslie Gaffney, Whitney Hornsby, Jordan W. Smoller, Alexander Gusev, Pradeep Natarajan, Giovanni Parmigiani

## Abstract

Understanding how disease patterns evolve over a lifetime remains a key challenge in medicine. While electronic health records provide rich longitudinal data, existing models typically analyze each disease in isolation, missing the complex interplay between conditions and genetic factors. Here, we present `ALADYNOULLI`, a dynamic Bayesian framework that integrates longitudinal health records with genetic data to identify latent disease signatures while modeling individual-specific trajectories. Applied in three biobanks with up to 52 years of follow-up, our model discovers clinically interpretable disease signatures that show remarkable cross-population consistency (median 80% composition preservation) and reveal distinct biological subtypes within traditional diagnostic categories, with large effect sizes for signature differences between patient clusters (Cohen’s *d* up to 4.25, *p* ≤ 1 × 10^−8^ for 95% of comparisons). Genetic validation demonstrates biological relevance through multiple complementary approaches: enrichment in known risk populations (familial hypercholesterolemia carriers, clonal hematopoiesis carriers), 151 genome-wide significant loci including novel cardiovascular associations in our dataset, rare variant associations with established disease genes (*LDLR*, *TTN*, *BRCA2*), and heritability exceeding component diseases. We also include a non-specific low-incidence signature which captures resistance across many disease conditions. The model’s explicit likelihood formulation enables principled corrections for selection bias through inverse probability weighting while preserving biological signal. For clinical prediction, `ALADYNOULLI` substantially outperforms established risk scores (PCE, PREVENT, GAIL) across 28 conditions over both 1-year and 10-year horizons. By jointly modeling genetics and longitudinal diagnoses, `ALADYNOULLI` achieves enhanced biological discovery and improved disease prediction through a unified, interpretable framework. Code and interactive results are available https://surbut.github.io/aladynoulli2/index.html with application at http://aladynoulli.hms.harvard.edu.

## Introduction

The risk of disease varies substantially between individuals and throughout life, with complex interactions between genetic predisposition, environmental factors, and accumulated comorbidities. Understanding these dynamic risk patterns could transform early detection, prevention, and personalized treatment strategies (*1–3*). The increasing availability of large-scale electronic health records (EHRs) linked to genetic data provides unprecedented opportunities to model these complex disease trajectories at a population scale (*4–6*). However, extracting meaningful patterns from these rich, longitudinal datasets remains challenging due to patient population heterogeneity, the temporal nature of disease progression, and intricate relationships between diverse conditions.

Traditional approaches to analyzing EHR data often focus on isolated diseases or simple pairwise associations, failing to capture how multiple conditions evolve together over time ((*7*)). Recent unsupervised methods have attempted to identify disease clusters or trajectories (*8*), but typically do not account for temporal dynamics of disease risk, biological variability underlying the same disease, or the influence of genetic factors on disease progression (*9,10*). Consider a patient who develops rheumatoid arthritis at age 45, followed by hypertension at 48, and eventually suffers a myocardial infarction at 52. Traditional approaches may treat these as separate events or simple comorbidities, missing the underlying metabolic-inflammatory process that drives this progression. In addition, they typically do not use information from patients with similar patterns to improve prediction for rare diseases, where limited data make traditional disease-specific models less reliable.

We present `ALADYNOULLI`, a generative model that integrates germline genetic data with longitudinal EHRs to identify latent disease signatures to model individual-specific health trajectories over time. `ALADYNOULLI` addresses these limitations by identifying shared disease signatures that capture biological processes common across multiple conditions, enabling more accurate prediction even for rare diseases through information sharing with related, more common conditions. Our approach offers several key advantages over existing methods: (1) **Interpretability**: it generates disease signatures which are not only computational representations, but correspond also to clinically meaningful biological processes; (2) **Temporal modeling**: it captures how disease risk evolves dynamically over the life course; (3) **Genetic integration**: it directly and interpretably incorporates genetic information into the model architecture; (4) **Unified framework**: it simultaneously models the majority of prevalent conditions in the EHR, sharing information across related conditions, and improving prediction even for diseases with limited data (*11*); (5) **Individual-specific trajectories**: it provides personalized risk profiles that adapt as new clinical information becomes available; and (6) **Principled bias adjustment**: is grounded in an explicit likelihood specification, supporting principled adjustment for selection bias through inverse probability weighting. In summary, jointly modeling multiple diseases and their genetic determinants, `ALADYNOULLI` enables both improved prediction of future disease risk and enhanced discovery of genetic architecture underlying complex phenotypes, while revealing meaningful patient subgroups with distinct biological mechanisms that could inform personalized interventions.

## Results

### ALADYNOULLI captures temporal disease signatures and individual trajectories

Disease patterns among individuals vary by onset, progression speed, and composition, reflecting different underlying biological mechanisms. `ALADYNOULLI` models the probability of each disease for an individual by integrating across multiple latent signatures (**Figure 1**).

**Figure 1:**
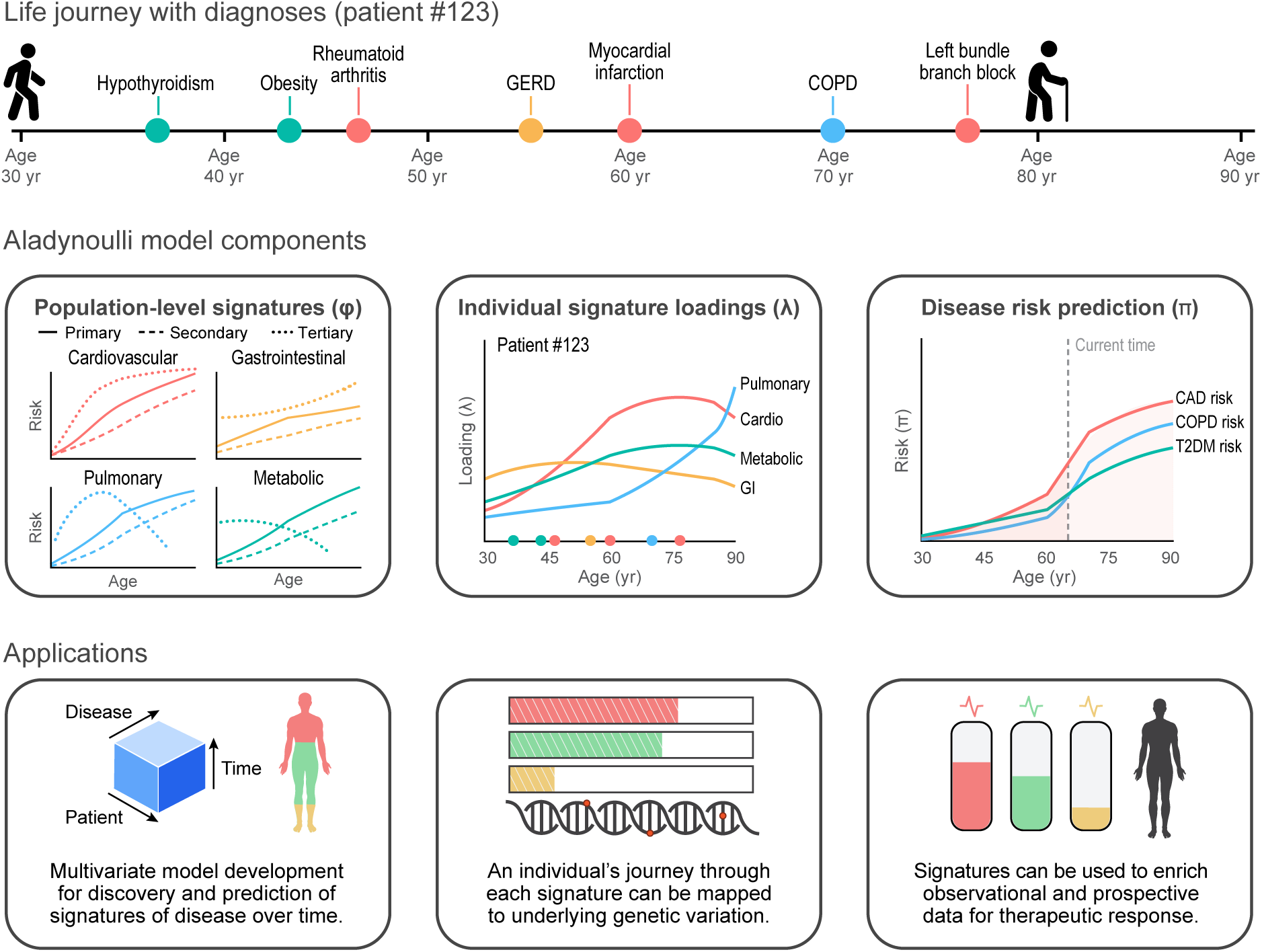
ALADYNOULLI model overview and applications. **Top:** Example patient timeline showing the sequence and timing of major diagnoses over the life course. **Middle:** Key model components. *Left:* Population-level disease signatures (*φ*), with each line representing the age-dependent risk trajectory for a specific disease within a signature. *Center:* Individual signature loadings (*λ*) transformed to *θ*via softmax, for a representative patient, showing how contributions from different signatures evolve over time. *Right:* Disease risk prediction (*π*) for selected diseases, integrating population-level signatures and individual loadings to generate personalized risk trajectories. **Bottom:** Applications of the model, including genomic discovery, therapeutic targeting, and patient matching (e.g., digital twin identification or stratification of patients with the same diagnosis but different risk profiles).

For each individual *i*, disease *d*, and time point *t*, *π_idt_* is the hazard of disease occurrence, that is, the probability of disease occurrence assuming the individual is still at risk for disease *d*. In `ALADYNOULLI` this is a weighted combination of signature-specific probabilities, where each signature captures patterns of diseases that tend to occur together (**Table S1)**:

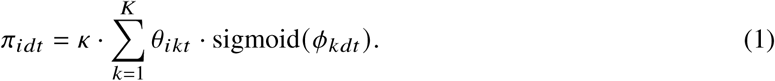

Here sigmoid(*ϕ_kdt_*) = 1/(1 + *e*^−^ *^ϕkdt^*), *κ* is a global calibration parameter, *θ_ikt_* is individual *i*’s normalized and time-varying association with signature *k* at time *t*, and *ϕ_kdt_* captures the relationship between signature *k* and disease *d* over time *t*. The normalized individual-signature associations (loadings) *θ_ikt_* are derived from latent variables *λ_ikt_* through a softmax transformation:

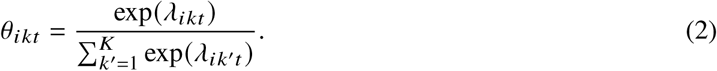

These latent variables *λ_ikt_* follow a Gaussian process (*12*) prior wherein we model the effects of genetic factors and time (see **Methods; Figure S3**). Specifically:

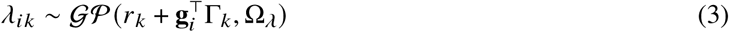

where *r_k_* is a signature-specific reference level in the population, Γ*_k_* captures genetic effects on signature predisposition, **g***_i_* represents individual genetic and demographic factors (36 polygenic risk scores, sex, and 10 genetic principal components; 47 features total) affecting the mean of *λ_ik_*, and Ω*_λ_* is a temporal covariance kernel modeling smooth trajectories for *λ_ikt_* over time.

Similarly, the disease-signature associations follow a Gaussian process prior:

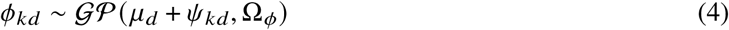

where *μ_d_* is a disease-specific baseline, or the logit of the population prevalence, *ψ_kd_* represents the overall strength of association between signature *k* and disease *d*, and Ω*_ϕ_* allows for temporal variation in these associations. Neatly, Ω*_ϕ_* and Ω*_λ_* govern the temporal covariance of deviations from the mean trajectory and are by construction independent from the mean. By specifying Gaussian process priors as offsets of mean parameters, our model encourages temporal fluctuations to be distinct from systematic effects like genetic predisposition or disease-signature associations.

A key innovation of our approach lies in its formulation as a mixture of probabilities rather than a probability of a mixture, as in traditional sparse factor analysis approaches (*13*). We also differ from allocation-based topic models that conditionally assign diseases to individuals *after* the event has necessarily occurred (*8*). In contrast `ALADYNOULLI` directly models the probability of disease occurrence as a weighted combination of signature-specific disease probabilities.

This crucial distinction allows our model to: (1) predict future disease onset rather than merely explain observed diagnoses; (2) accommodate multiple contributing disease processes simultaneously rather than forcing competitive allocation to a single signature; and (3) accurately model chronic conditions that persist over time rather than treating each diagnosis as an independent event. The combination of softmax-transformed individual loadings (*θ*) and sigmoid-transformed disease probabilities (*ϕ*) ensures proper probability scaling. Importantly, given the signature loadings *θ_ikt_* and associations *ϕ_kdt_*, diseases are conditionally independent—all disease correlations are mediated through shared signatures.

#### Terminology clarification

The factor analysis literature exhibits inconsistent terminology that can be confusing. In some traditions (e.g., sparse factor analysis (*13, 14*)), “loadings” refer to individual-specific weights (our *λ* parameters), while “factors” or “coefficients” refer to feature importance (our *ϕ* parameters). In other traditions, “loadings” refer to feature importance (our *ϕ* parameters), while individual components are called “scores” or “weights” (our *λ*parameters). Throughout this work, we use “loadings” to refer to individual-specific signature associations (*λ* and *θ*) and “signature-disease associations” to refer to feature importance (*ϕ*), consistent with the sparse factor analysis convention.

#### Two complementary applications

`ALADYNOULLI` serves two distinct but complementary purposes, each requiring different analytical approaches. For biomedical discovery, `ALADYNOULLI` operates with complete hindsight, leveraging entire patient trajectories to maximize our ability to identify biological patterns and mechanisms. This retrospective analysis transforms our understanding of disease patterns, progression speed, genetic relationships, and disease associations by using all available longitudinal data to characterize disease signatures, quantify genetic influences, and reveal patient heterogeneity within diagnostic categories. For clinical prediction, we operate under strict temporal constraints that mirror real-world clinical decision making (see **Figure S3** for distinction). We employ a rigorous temporal validation framework that uses only information available up to a prediction time point (see **Figure S10**). This prospective approach reflects real-world clinical use, where physicians must predict future risk based solely on a patient’s history to date, and ensures our performance metrics reflect true predictive capability rather than retrospective explanation. This is consistent with a well-established body of work on validation and clinical use of risk stratification ((*15*), landmarking approaches (*16*) and survival analysis (*17*)).

### Applying ALADYNOULLI identifies consistent signature patterns across diverse populations

We applied `ALADYNOULLI` to three independent biobank: UK Biobank (UKB, n=427,239), Mass General Brigham (MGB, n=48,069), and All of Us (AoU, n=208,263) (**Table S2, Figure S14**). We obtained ICD-10 codes from hospitalization diagnoses in each (*4, 18*) and transformed these to pheCodes (*19*), following established approaches for EHR phenotyping (*20*) (see **Methods**). A set of 348 pheCodes were selected representing diseases with at least 1000 unique occurrences in UK Biobank hospitalization episode statistics (*21*) as in (*8*). These same pheCodes were also present in AoU and MGB(see **Methods**). Despite differences in population characteristics, healthcare systems, and data collection methodologies across these cohorts, our model identified remarkably consistent signature patterns (**Table S3; Figures S5, S15, S16, S17**).

We set the number of signatures for training to *K* = 21 comprising 20 disease signature and one health signature. The signatures we estimated are stable both across cohorts and across data subsets (batches) within a cohort (**Figure S1**). They capture distinct and recognizable disease processes including cardiovascular, metabolic, pulmonary, psychiatric, musculoskeletal, and oncologic conditions (see **Table S3** for full characterization). Each signature demonstrates characteristic temporal patterns, with disease probabilities evolving dynamically with age. These are stable across biobanks (**Figure 2A; Figures S15, S16, S17**). For example, the non-ischemic cardiovascular signature shows steadily increasing probabilities for conditions like atrial fibrillation and heart failure after age 55 years, while the malignancy signature displays a sharp rise in metastatic disease probabilities between ages 60-75 years. The *ψ_kd_* capture the strength of association between diseases and signatures and vary within signatures in a way that is empirically determined during training (**Figure 2B**). The model also captures clinically expected temporal disease progression: hypercholesterolemia precedes myocardial infarction within the ischemic cardiovascular signature, while primary malignancies precede metastatic disease within the cancer signature (**Figure 2E**). These nuanced temporal relationships emerge directly from the model without explicit encoding, demonstrating `ALADYNOULLI`’s ability to learn clinically meaningful disease trajectories.

**Figure 2:**
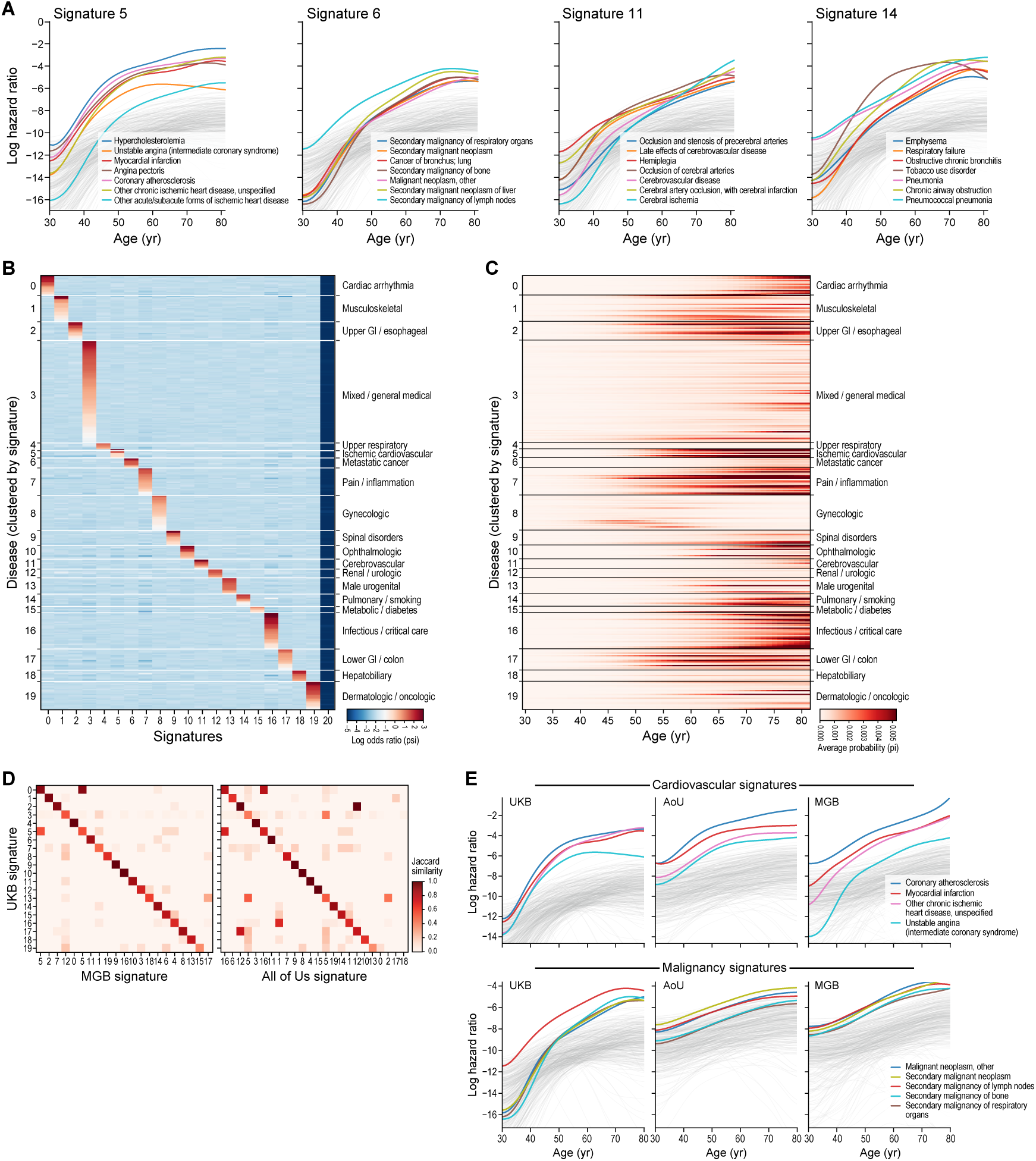
Population-level disease signatures inferred by ALADYNOULLI. (**A**) Age-dependent log hazard ratios (*ϕ_kdt_*) for four representative disease signatures (Ischemic Cardiovascular, Signature 5; Metastatic Cancer, Signature 6; Pulmonary/Smoking, Signature 14; and Cerebrovascular, Signature 11) from UK Biobank (UKB). Parameters shown are posterior estimates of time-varying disease-signature associations (*ϕ_kdt_*) pooled across 40 batches. Each colored line represents the log hazard ratio trajectory for a specific disease within the signature, with grey lines showing background diseases.(**B**) Heatmap of posterior static signature-disease association parameters (*ψ̂_kd_*) averaged across 40 UKB batches. Diseases are ordered by their signature assignment (determined by maximum posterior *ψ*value, argmax*_k_ ψ_kd_*), then sorted within each signature by descending posterior *ψ* value (strongest associations at top). (**C**) Model-predicted age-specific disease probabilities (*π̄_dt_*) averaged over individuals (*N* = 400, 000). For each disease *d* and age *t*, 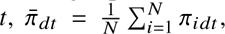 where *π_idt_* is computed from Equation 1 using posterior pooled *ϕ* parameters and posterior individual-specific *λ* loadings. Disease ordering matches Panel B. (**D**) Heatmaps showing composition preservation probability between UKB and two validation cohorts (Mass General Brigham (MGB), left; All of Us (AoU), right). Disease-to-signature assignments are determined using maximum posterior *ψ* values (argmax*_k_ ψ_kd_*). For each UKB signature *k*, the composition preservation probability is calculated as the proportion of diseases in that signature that also belong to its best-matching signature in the comparison cohort. Median composition preservation probability: 0.838 (83.8%) for MGB and 0.782 (78.2%) for AoU. (**E**) Cross-cohort comparison of signature trajectories for Ischemic Cardiovascular (Signature 5 in UKB/MGB, Signature 16 in AoU) and Metastatic Cancer (Signature 6 in UKB/MGB, Signature 11 in AoU) signatures. Parameters shown are posterior estimates of time-varying disease-signature associations (*ϕ_kdt_*) from three independent cohorts: UK Biobank (posterior pooled *ϕ* from 40 batches), All of Us (posterior pooled *ϕ* from 25 batches), and MGB (posterior *ϕ* from single trained model). Only diseases present in all three cohorts are shown.

The average age-specific hazard for a wide range of diseases are visualized in **Figure 2C**, highlighting temporal risk patterns. Furthermore, the model’s tensor structure (**Figure S13**), enables rapid disease hazard calculation using the average loadings (**Equation 3**) and population-level *ϕ_kd_*.

These signature patterns show strong consistency across the three independent biobanks (**Figure 2D; Figures S15, S16, S17**). For example, when comparing the membership of diseases within signatures between any two biobanks (with disease-to-signature assignments determined by posterior fitting using the signature with maximum posterior association strength, argmax*_k_* (*ψ_kd_*)), we observed high concordance (median composition preservation probability index = 0.80, IQR = 0.667, 0.964) across all pairwise comparisons between biobanks, **Figure S5**). **Figure 2E** illustrates this consistency for two important signatures: cardiovascular disease and malignancy. Despite differences in healthcare system and coding practices, the temporal patterns of key diseases within these signatures remain remarkably consistent, supporting the biological validity of the patterns discovered.

### Personalized trajectories reveal heterogeneity within disease categories

Beyond population-level signatures, `ALADYNOULLI` provides individual-specific trajectory information through the time-varying *λ_ikt_* parameters that reveal distinct disease progression patterns.

Figure 3A illustrates how individual disease journeys reveal biological differences among patients sharing the same diagnosis, reflecting heterogeneity—i.e., the presence of distinct subgroups with different underlying disease signature distributions—within the diagnostic category. This example patient (Patient Encoded Identifier 148745) demonstrates a complex trajectory with multiple diseases across diverse signatures. The figure shows the patient’s signature loadings (*θ*) over time, their disease timeline, and the time-varying disease probabilities (*π*) for diagnosed conditions, demonstrating how `ALADYNOULLI` integrates multiple data types to provide a comprehensive view of disease progression. Additional examples (**Figure S6**) demonstrate the diversity in temporal loadings that would be missed by an approach that considered only average loading. Our model also illustrates how individual-level trajectories and population phenomena combine to produce time-varying personalized disease probabilities (**Figure S25**): overall risk can be decomposed into the contributions of an individual’s time-varying population level signature loadings, demonstrating the complex interplay between multiple signatures in determining disease risk.

**Figure 3:**
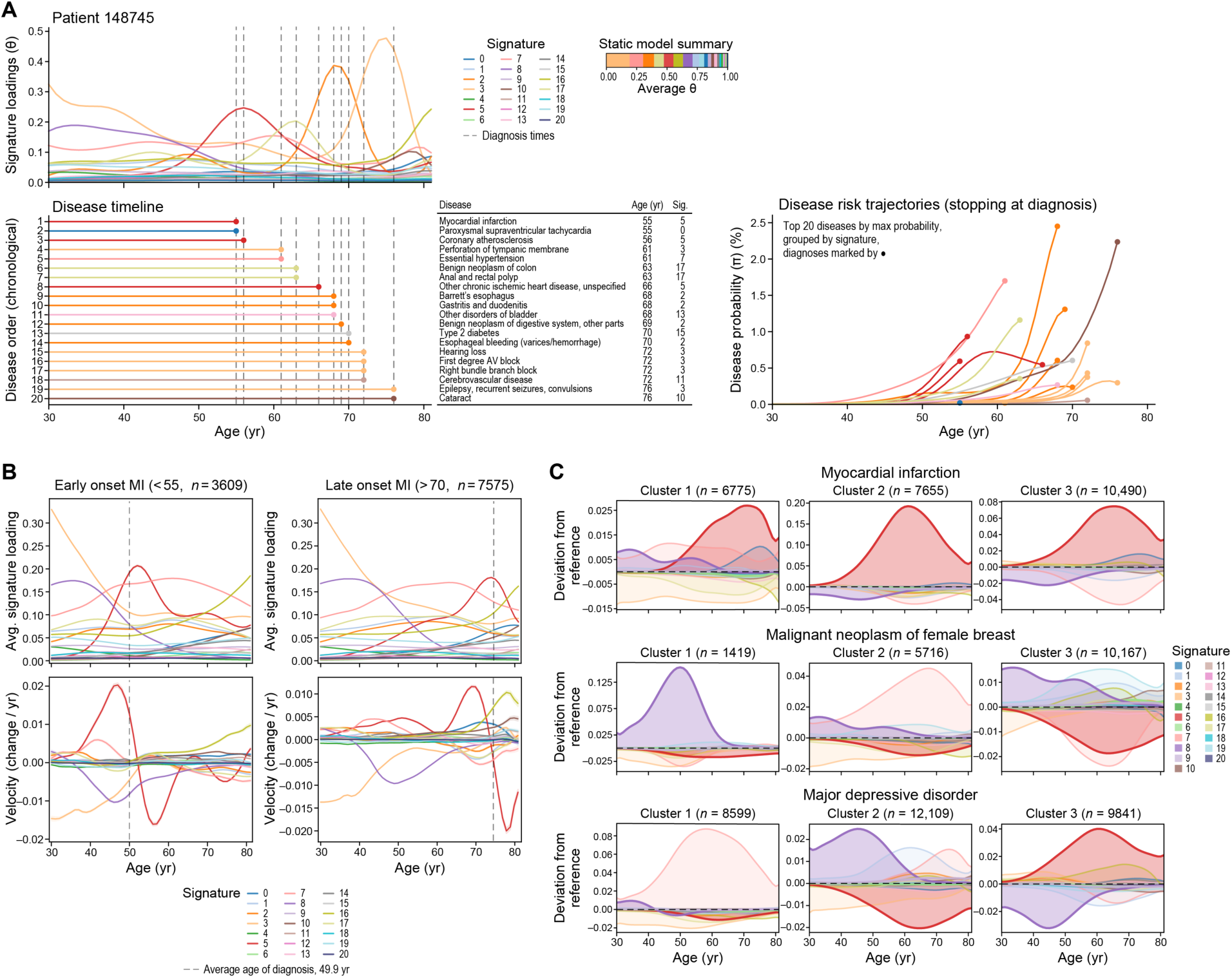
Individual-level trajectories and dynamic risk profiles. (**A**) Patient-specific posterior normalized signature loadings (*θ_ikt_* = softmax *λ_ikt_*) and disease probabilities (*π_idt_*) for a representative individual (Patient 148745, 20 diseases, ages 55-76). This patient was selected as a representative example demonstrating multiple diseases spanning a substantial time period with diverse signature involvement. **Top panel:** Posterior signature loadings (*θ_ikt_*) calculated as softmax of posterior individual signature loadings (*λ_ikt_*) from trained model, showing temporal evolution of 21 signatures. Inset shows average *θ* across time (stacked bar). **Middle panel:** Disease timeline showing chronological order of diagnoses with associated signatures. Diagnoses are marked by black circles. **Bottom panel:** Posterior disease probabilities (*π_idt_*) calculated from posterior pooled *ϕ_kdt_*, posterior individual *λ_ikt_*, and posterior *κ* using the model’s nonlinear transformation, showing top 20 diseases by maximum probability grouped by signature. **X-axis:** Age (years, 30-80). **Y-axis (top):** Normalized signature loading *θ* (0-1). **Y-axis (bottom):** Disease probability *π*(%). (**B**) Comparison of early-onset myocardial infarction (MI) (*<*55 years, n=3,609) and late-onset MI (≥70 years, n=7,575) showing posterior signature loadings and their temporal velocities. **Left panels:** Average posterior signature loadings (*θ̄kt*) calculated as mean of softmax(*λ_ikt_*) across patients in each group, where *λ_ikt_* are posterior individual signature loadings from trained models. Vertical dashed line indicates average age of diagnosis for each group. **Right panels:** Temporal velocity (change per year) calculated as gradient of mean signature loadings over time: 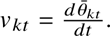 **X-axis:** Age (years, 30-80). **Y-axis (left):** Average signature loading (0-1). **Y-axis (right):** Velocity (change/year). (**C**) Signature heterogeneity within disease subtypes: Deviations from population reference for three diseases (Myocardial infarction, Malignant neoplasm of female breast, Major depressive disorder). For each disease, patients with the disease were clustered into 3 groups using k-means clustering on time-averaged posterior signature loadings 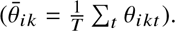 For each cluster at each timepoint, mean signature loadings were calculated and deviations from population reference (*θ_kt_*^ref^) were computed: 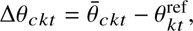 where *θ̄_ckt_* is the mean posterior *θ* for cluster *c*at time *t*and *θ_kt_*^ref^ is the population average posterior *θ* for signature *k*at time *t*. Each row shows one disease with three clusters (n shown in parentheses). **X-axis:** Age (years, 30-80). **Y-axis:** Deviation from reference (difference in normalized signature loading, Δ*θ*).

Aggregating these individual patterns reveals distinct group-level differences. In a retrospective analysis, the comparison of early-onset (≤ 55 years, mean age of event 49.9 years) and late-onset (≥ 70 years, mean age of onset 74.6 years) MI in Figure 3B shows that early-onset patients exhibit a higher and earlier peak in their Signature 5 contribution, as well as a more rapid increase in signature loading prior to the event, compared to late-onset cases. These quantitative differences in trajectory characteristics suggest that early- and late-onset MI, while sharing the same clinical diagnosis, may result from distinct disease processes requiring different preventive strategies.

To intuitively illustrate differences in signature composition among patients with the same clinical diagnosis for three representative diseases (myocardial infarction, breast cancer, and major depressive disorder), we applied a descriptive k-means clustering to patients’ time-averaged signature loadings for each disease (Figure 3C). This two-step approach first identifies patient subgroups based on their average signature associations, then visualizes temporal deviations from population reference trajectories for each cluster, providing clinically interpretable patient stratification. A complementary deviation-based clustering approach confirmed these patterns across biobanks (**Figure S32**).

We then calculated cluster-specific Cohen’s effect sizes (*22*) *C_ck_*^SIG^ as follows (**Figure S18; Extended Data)**. For cluster *c* and signature *k*, *C_ck_*^SIG^ is the standardized difference in mean time-averaged signature loadings between individuals in cluster *c* and those in all other clusters (see **Figure S18**). This is a measure of how distinct each cluster is with regard to each disease signature.

This analysis revealed that the majority of signature differences between clusters were not only large in magnitude (with many *C_ck_*^SIG^ values exceeding 0.8, and some as high as 2.5–4.2), but also highly statistically significant (p ≤ 1×10^−8^ for nearly all clusters). The largest effect size occurs in major depressive disorder, for the pain/inflammatory/metabolic signature 7 (characterized by asthma, migraine, osteoporosis, depression, and obesity) in cluster 1, which showed *C*_1,7_^SIG^ = 2.76 (p ≈ 0), revealing a depression subgroup with strong inflammatory comorbidities. In myocardial infarction, signature 5 (encompassing coronary atherosclerosis, ischemic heart disease, and hypercholesterolemia) shows *C*_2,5_^SIG^ = 2.82 (p ≈ 0) in cluster 2, indicating substantial heterogeneity between patient subgroups. In breast cancer, the gynecologic signature 8 shows the strongest differentiation (*C*_1,8_^SIG^ = 4.25, p ≈ 0), while the pain/inflammatory/metabolic signature (Signature 7) achieved near-complete patient separation in breast cancer cluster 2 (*C*_2,7_^SIG^ = 2.53, p ≈ 0).

We quantified PRS variability across patient clusters using Cohen’s effect sizes *C_cp_*^PRS^, revealing substantial differences in polygenic risk scores between patient subgroups that parallel the biological variation observed in signature loadings (Figure 4B**; Extended Data**). These clusters show elevated disease-specific PRS patterns (e.g., cardiovascular PRS enrichment in myocardial infarction clusters), confirming the biological meaningfulness of the identified patient subgroups. Taken together, these results demonstrate that the observed heterogeneity is both quantitatively substantial and statistically robust, supporting the biological relevance of the patient subgroups we identified (see **Extended Data S1-S3**).

**Figure 4:**
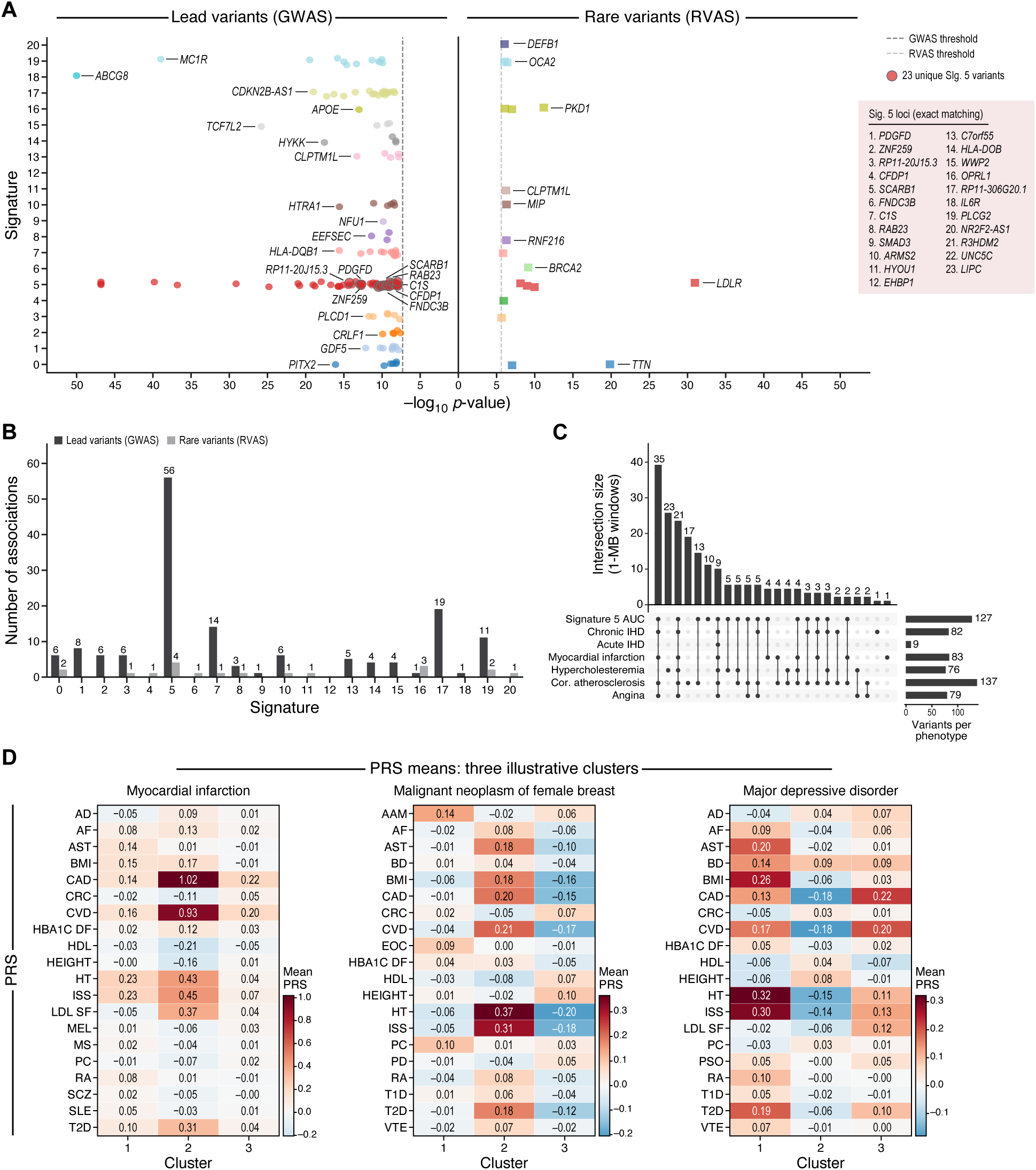
Genetic architecture and polygenic risk stratification of ALADYNOULLI disease signatures. (**A**) Lead variants from GWAS (left, 151 lead variants across our 21 signatures) and rare variants from RVAS (right, mask3 LoF variants, 59 associations, 18 unique genes). Signature 5: 23 unique loci (green; exact lead-SNP matching vs constituent-trait GWAS; 10 unique loci by ±1MB window overlap). **X-axis:** -log10 p-value. **Y-axis:** Signature (0-20). (**B**) Number of genetic associations per signature: lead variants (GWAS, dark grey) and rare variants (RVAS, light grey). Signature 5 shows 56 total associations. (**C**) UpSet plot showing overlap of genome-wide significant loci between Signature 5 AUC and cardiovascular traits (1-MB window overlap). Largest intersection: 35 variants shared across all six cardiovascular traits plus Signature 5 AUC. (**D**) Mean PRS values by cluster for three diseases (Myocardial infarction, Breast cancer, Major depressive disorder). Patients clustered as in Figure 3C; top 20 PRS traits by absolute Cohen’s d shown. Red: positive PRS (higher genetic risk); blue: negative PRS (lower genetic risk). Cohen’s d values in Supplementary Data.

### Integration of ancestry differences and selection biases

We also investigated how genetic ancestry influences signature loadings across the life course, revealing that ancestry-specific signature enrichment patterns vary with age (**Figure S22**). Genetic ancestry is now explicitly modeled through principal components (PCs) of genotype matrix. For South Asian ancestry, cardiovascular Signature 5 shows strong enrichment that peaks around ages 50-60 (deviation 0.025-0.03 when PCs are excluded), and the effect of ancestry is larger when PCs are included (peak deviation 0.06 at age 60, remaining elevated at 0.04 by age 80). East Asian ancestry shows enrichment in Signature 8 and Signature 9, with patterns that become more pronounced when PCs are included. These temporal patterns demonstrate that ancestry effects on signatures evolve across the life course, with peak deviations occurring at different life stages for different ancestries.

### Genetic factors influence signature trajectories

A key innovation of `ALADYNOULLI` is its integration of genetic information directly into the model through the Γ*_k_* parameters, which capture how polygenic risk scores (PRS) influence signature loadings. Using a comprehensive collection of 36 externally validated PRS developed independently of our signature analysis (*23*), we identified 116 significant PRS-signature association coefficients out of 756 candidate combinations (FDR *<* 0.05 using Benjamini-Hochberg correction on two-sided posterior tail probabilities) (**Supplementary Figure S27**; see Extended Data S0). We observed the strongest genetic effects for signatures with known heritable components: coronary artery disease PRS with the cardiovascular signature (Signature 5, *γ* = 0.153, Z = 27.2, p *<* 10^−15^), LDL cholesterol PRS with Signature 5 (*γ* = 0.071, Z = 22.7, p *<* 10^−15^), and type 2 diabetes PRS with the metabolic signature (Signature 15, *γ* = 0.154, Z = 58.3, p *<* 10^−15^). Highly significant Z statistics (Extended data *Annotation*/*gamma_a_ ssociations.csv*) demonstrate that genetic factors directly influence disease signature trajectories, consistent with the high heritability of these disease categories.

For the two genetic association analyses that follow, we trained a modified model to systematically identify genetic loci associated with signature trajectories. We defined a phenotype of ‘lifetime signature exposure’ for each signature, computed as the area under each individual’s signature loading curve over their follow-up period (**Figure S4; Methods**). Crucially, to avoid circularity, we removed the PRS term (**g***_i_*^⊤^ Γ*_k_* in Equation 3) from the prior on *λ* and re-trained signature loadings. We performed common variants genome-wide association analyses (GWAS) on the ‘lifetime signature exposure’ phenotype, testing 6,418,439 imputed variants (MAF ≥ 0.01, INFO ≥ 0.7) that passed quality control (**Methods**).

Linkage disequilibrium score regression (LDSC) (*24*) on 930,186 variants with available LD scores revealed significant SNP-based heritability for multiple signatures (Table S11), with the strongest signal observed for the cardiovascular signature (*ℎ*^2^ = 0.0414, SE = 0.003), followed by musculoskeletal (*ℎ*^2^ = 0.035, SE = 0.002) and pain/inflammation signatures (*ℎ*^2^ = 0.027, SE = 0.002). Critically, because our signature phenotypes are *continuous* (integrated area under the signature loading curve), heritability is inherently reported on the observed scale—there is no liability-scale transformation. For valid comparison with reported results, we therefore computed observed-scale heritability for component binary traits using the same LDSC procedure on the same UK Biobank data; signature heritability met or exceeded that of constituent traits on this comparable scale (**Table S12**).

Common variant GWAS identified 151 genome-wide significant loci across our 21 signature GWAS, with the cardiovascular signature (Signature 5) alone accounting for 56 lead loci. The strongest associations revealed signature-specific genetic architectures: Signature 5 showed strong signals at LPA (rs10455872), APOE (rs7412), and PCSK9 (rs11591147), all established lipid metabolism genes (*25,26*). The metabolic/diabetes signature identified TCF7L2 (rs7903146), the strongest known type 2 diabetes risk variant (*27*), while the musculoskeletal signature identified GDF5 (rs143384), a well-established osteoarthritis gene (*28*), the ophthalmologic signature (Signature 10) captured HTRA1 and CFH, key age-related macular degeneration gene; while the hepatobiliary signature (Signature 18) showed the strongest overall signal at ABCG8 (rs11887534), a gallstone risk gene, while also revealing additional loci warranting further investigation. Cross-signature connections also revealed shared genetic architecture: APOE appeared in both the cardiovascular signature (Signature 5) and the infectious/critical care signature (Signature 16), reflecting its dual role in cardiovascular and neurodegenerative disease risk (Extended Data GWAS).

We found that 23 of Signature 5’s 56 annotated loci were not found as lead loci in our own single-trait constituent cardiovascular GWAS using exact lead-SNP matching, including associations near genes with established roles in cardiovascular disease (*29*) (*IL6R* (rs6687726), *SCARB1* (rs11057839), *SMAD3* (rs56375023), *PDGFD* (rs1384705), and others) (**Table S4**, Extended Data). Using ±1MB window overlap instead of exact lead-SNP matching, 10 of the 56 Signature 5 lead loci have no constituent-trait lead locus within ±1MB in our analyses. Importantly, within our study and analysis pipeline applied to the same UK Biobank data, joint multi-disease signature modeling enables detection of associations that were not detected as lead loci in the corresponding single-trait constituent GWAS (angina pectoris, myocardial infarction, coronary atherosclerosis, hypercholesterolemia, acute IHD, chronic IHD), consistent with distributed pleiotropic effects that are too weak to detect individually in our sample but become detectable when aggregated across related phenotypes. While some of these loci (e.g., *IL6R*) have been identified in large-scale external CAD GWAS meta-analyses, they were not detected in our single-trait analyses, demonstrating that signature-based modeling enhances discovery power even within the same dataset. Among these, loci annotated to *WWP2*, *C1S*, *HYOU1*, *EHBP1*, *R3HDM2*, and *OPRL1/LKAAEAR1* (and *ARMS2*) appear among the more plausible candidates for follow-up given more limited prior evidence as CAD loci. Accordingly, when associating significant loci in each signature with component trait genotype dosage, we found similar improvements across signatures (**Figure S7**), supporting that our latent signatures reproduce constituent-trait associations and in many cases identify additional variants, demonstrating enhanced genetic discovery power of latent signatures.

We also performed gene-based rare variant association studies (RVAS) using aggregated rare variants within genes, testing whether gene-level rare variant burden is associated with signature exposure. This analysis tested 18,464 genes per signature using REGENIE gene-based association tests with loss-of-function and deleterious missense variants (Mask3: LoF, DelMis09, DelMis08), identifying 18 unique genes with genome-wide significant associations (p *<* 2.5 × 10^−6^ after Bonferroni correction) across 21 signatures. The ischemic cardiovascular signature (Signature 5) demonstrated association with four significant genes after false discovery correction - *LDLR*, *APOB*, *LPA* and *CDH26* - all with established roles in lipid metabolism and cardiovascular disease (*30, 31*). Signature 0 (heart failure/arrhythmia) showed a highly significant association with *TTN* (p = 1.0 × 10^−21^), encoding the cardiac structural protein titin (*32*), while Signatures 16 (critical illness/inflammation) was associated with *TET2*, *PKD1*, and *BRCA2* (Extended data, RVAS; S15). *TET2* is highly relevant to this signature’s component diseases (sepsis, infections, neutropenia, thrombocytopenia, anemias), as *TET2* mutations cause clonal hematopoiesis of indeterminate potential (CHIP), which is associated with chronic inflammation, increased infection risk, and blood cell dysfunction (*33*). *PKD1* mutations relate to kidney dysfunction and could contribute to acute renal failure and electrolyte disorders that characterize this signature (*34*). The ophthalmologic signature (Signature 10) was associated with *MIP*, encoding major intrinsic protein of lens fiber, which is essential for lens transparency and cataract formation (*35*), validating the signature’s capture of age-related eye disease pathways. These rare variant associations were robust across multiple variant masks (tested across six progressively inclusive functional impact categories) (**Figure S28C**; supplementary data). Finally, for each signature-gene pair identified through RVAS, we computed correlations between rare variant burden and disease presence across all diseases. We then plotted gene-disease correlations (x-axis) against the corresponding disease-signature loadings (y-axis) for diseases with statistically significant gene-disease correlations (p *<* 0.05). The y-axis displays the maximum signature-disease association over time (max*_t_ ϕ_kdt_*) transformed via sigmoid to probability scale, representing peak signature-disease association. We computed the correlation between these two vectors (S29).

To further demonstrate the biological relevance of our signatures, we examined signature enrichment in specific high-risk populations with known genetic predispositions (**Figure S23**). For example, familial hypercholesterolemia (FH) is caused by mutations in LDLR, APOB, or PCSK9, leading to severely elevated LDL cholesterol and premature cardiovascular disease (*36*), making FH carriers an ideal validation population for cardiovascular signature enrichment. We compared the proportion of FH carriers versus non-carriers who showed a pre-event rise in cardiovascular Signature 5 loadings (defined as an increase over the 5 years preceding their first cardiovascular event). FH carriers showed significantly higher rates of pre-event Signature 5 activation compared to non-carriers (odds ratio = 1.63, Fisher’s exact test p = 0.017), providing evidence that Signature 5 captures cardiovascular risk pathways.

We also validated signature associations using clonal hematopoiesis of indeterminate potential (CHIP) carriers, demonstrating enrichments that provide independent support for the biological interpretability of signatures. CHIP mutations cause chronic inflammation and are associated with increased risk of hematologic malignancies, cardiovascular disease, and other inflammatory conditions (*37*). Our analysis reveals that CHIP carriers show strong enrichment in Signature 16 (Critical Care/Inflammation) before multiple outcomes: DNMT3A carriers show 1.97-fold enrichment (OR=1.97, p=0.0007) for Signature 16 before Leukemia/MDS events, with 81.1% of carriers showing rising trajectories compared to 68.5% of non-carriers (**Figure S23B**). TET2 carriers similarly showed enrichment in Signature 16 before cardiovascular and inflammatory outcomes, consistent with CHIP’s known association with chronic inflammation and increased cardiovascular risk (*37*).

#### Inverse probability weighting enables principled bias correction

As the UK Biobank may be enriched in healthy individuals ((*38*)), we assessed the impact of such potential bias on the model with inverse probability weighting (IPW). A key advantage of `ALADYNOULLI`’s model structure is its ability to adjust for this selection bias by weighting individual terms of the likelihood function inversely to their probability of participation. This preserves interpretation of all model components, including biological disease-signature relationships—a capability that distinguishes our approach from black-box machine learning methods that cannot easily incorporate such principled corrections.

We validated IPW through two complementary approaches. First, we performed a controlled experiment by artificially creating a biased population, randomly dropping 90% of women from the full UK Biobank sample (reducing the sample from 400,000 to 204,287 individuals, with women comprising only 10.6% instead of 54.4%). To isolate the effect of selection bias on parameter estimation, we retrained all model parameters (*ϕ*, *λ*, *γ*, *ψ*) while holding *μ_d_* fixed across all three scenarios (full cohort, biased cohort without IPW, and biased cohort with IPW). We successfully recovered the full population patterns using IPW: signature-disease associations (*ϕ*) remained stable, while disease probabilities (*π*) and *empirical prevalence* trajectories (observed disease rates calculated from the data, not the model parameter *μ_d_*) for diseases with expected sex bias (e.g., Breast and Prostate cancer) matched those trained in the full population (**Figure S20**). Second, we applied IPW to correct for real-world UK Biobank participation bias using a published (*39*) lasso-regularized logistic regression model trained on demographic variables (region, birth year, sex, household characteristics, employment status, tenure, ethnicity, self-reported health, and education) to estimate probabilities of UK Biobank participation. We calculated weights as the ratio of the general population selection rate (4.34%) to each individual’s predicted selection probability, trimming at the 1st and 99th percentiles (weight range: 0.17-6.63, mean: 0.93, median: 0.59). These weights were incorporated into the `ALADYNOULLI` likelihood function by multiplying each individual’s contribution by their IPW (Eqn. 7), with cross-biobank validation across UK Biobank, Mass General Brigham, and All of Us demonstrating that signature patterns remain consistent across selection mechanisms.

In both validation experiments, the signature-disease associations (*ϕ*) remain stable (correlation *>* 0.999, mean absolute difference *<* 0.002), demonstrating that core biological patterns are preserved, while individual loadings (*λ*) adapt to reflect the reweighted population distribution. This structural separation enables principled bias correction while maintaining interpretability.

### Dynamic risk assessment improves disease prediction

A primary motivation for modeling longitudinal disease patterns is to improve prediction of future disease events. To rigorously evaluate `ALADYNOULLI`’s predictive performance, we implemented comprehensive, leakage-free validation strategies that mimic real-world clinical follow-up (**Table S5**).

#### Primary evaluation: Dynamic 1-year predictions

Our primary evaluation uses a temporal validation approach (evaluating predictions at multiple fixed timepoints) (*16*), where we compute 1-year risk predictions at 10 distinct time points during follow-up (enrollment + 0, 1, 2, …, 9 years). At each time point, we retrain models using only data available up to that point and evaluate performance on 1-year outcomes. We report the median AUC across these 10 dynamic 1-year predictions, which captures how predictive accuracy evolves as patients accumulate new diagnoses over time and reflects how the model would be used in clinical practice.

All individuals contribute EHR data from age 30+ onwards (or first EHR access), providing decades of longitudinal disease history. Individuals enrolled in UK Biobank between 2006-2010 at ages 40-70 years (median age 54, see **Figure S10**) and were followed for a median of 14.4 years until 2023/2024 (see **Figure S11**).

All predictions were generated using leave-one-out cross-validation (excluding each batch from training when generating its predictions) to ensure robust, generalizable performance estimates. This primary evaluation demonstrates robust performance across diseases (ASCVD: 0.879, Breast Cancer: 0.867, Atrial Fibrillation: 0.801, Heart Failure: 0.811, Parkinson’s: 0.796, **Table S6**).

#### Comparison with existing benchmarks: 1-year and 10-year predictions

For comparison with traditional clinical risk scores, we also evaluated predictions made at recruitment (using models trained with data up to enrollment) against both 1-year and 10-year outcomes (ALADYNOULLI 1-year, 10-year). This comparison between `ALADYNOULLI` and traditional risk scores is only feasible at enrollment (i.e., 2006-2010 in UKB, **Fig S10**) because of availability of the necessary input. At enrollment, 1-year predictions show good performance (ASCVD: 0.881, Breast Cancer: 0.782, Atrial Fibrillation: 0.797, Heart Failure: 0.769). 10-year predictions, as expected, are less precise, but we maintain predictive power (ASCVD: 0.733, All Cancers: 0.674, Diabetes: 0.651, **Table S6**). It is important to note that for the vast majority of the 348 conditions considered, there is no established lifetime risk model in current clinical practice. All analyses were performed strictly prospectively, ensuring that only data available up to prediction time was used for each individual predicted. Individuals with prevalent disease at prediction time were excluded (see **Methods**).

We also evaluated a dynamic 10-year rolling interpolation approach that demonstrates the model’s capabilities with time-dependent information: by updating predictions annually and aggregating risk over a 10-year horizon (similar to time-dependent covariates, **Table S8**), this approach shows how the model’s probability estimates evolve as new information becomes available. Note that this approach uses information at each year and is therefore not directly comparable to standard prediction evaluations, but provides insight into how model performance improves when incorporating time-dependent covariates (see **Extended Methods; Table S5**). Finally, we compared with traditional cox modeling (*17*) using age as a time scale, also on ten-year outcomes, using sex and family history when available as a predictor.

Comprehensive calibration analysis demonstrates calibration across the full range of predicted risks (MSE = 4.67 × 10^−7^), with mean predicted rates (5.55 × 10^−4^) closely matching mean observed rates (5.45 × 10^−4^) across 722,346,330 patient-time observations (at-risk only) (Figure 5D). The calibration plots show alignment between predicted and observed rates.

**Figure 5:**
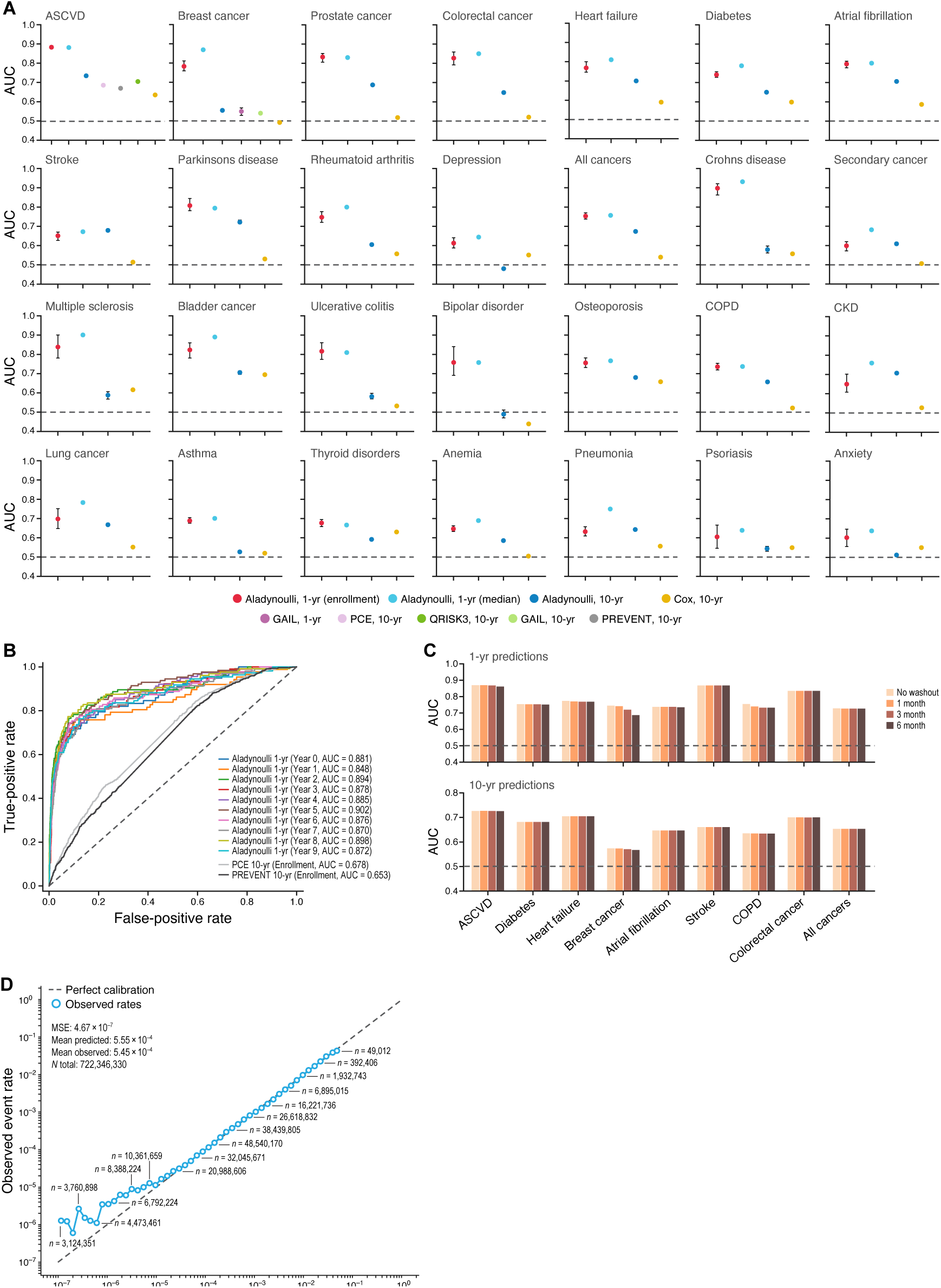
Multi-Disease Risk Prediction Performance and Model Interpretation. (**A**) AUC for 1-year and 10-year predictions across 28 diseases. Methods: Aladynoulli 1-year (enrollment, red); Aladynoulli 1-year (median, light blue, median AUC across Years 0-9); Aladynoulli 10-year (enrollment, dark blue); Cox 10-year (orange, age/sex/family history); PCE/PREVENT/QRISK3 10-year (ASCVD only); GAIL 1-year/10-year (Breast cancer female only). All analyses exclude prevalent disease and use only data available at prediction time. (**B**) ROC curves for Aladynoulli 1- year predictions (Years 0-9, colored lines; Year 0: AUC=0.881, Years 1-9: AUCs 0.848-0.902) versus PCE (AUC=0.678) and PREVENT (AUC=0.653) for ASCVD in this 10,000-person held-out test set (PCE 0.683 and PREVENT 0.667 in the overall cohort). (**C**) Washout period impact: AUC for 1-year (top) and 10-year (bottom) predictions with 0, 1, 3, or 6 month exclusions before prediction timepoint. Bars: no washout (light brown), 1 month (medium brown), 3 months (darker brown), 6 months (darkest brown). (**D**) Calibration plot (log-log scale): predicted versus observed event rates. Posterior *π_idt_* calculated from pooled *ϕ_kdt_*, individual *λi kt*, and *κ* at enrollment. 50 bins in log space (min 10,000 observations/bin). MSE = 4.67 × 10^−7^, mean predicted = 5.55 × 10^−4^, mean observed = 5.45 × 10^−4^, *N* = 722,346,330 observations.

We further evaluated `ALADYNOULLI`’s performance for ASCVD (atherosclerotic cardiovascular disease) risk prediction, first in the general population (**Fig S12**) and then in specific high-risk subgroups. In the overall cohort, `ALADYNOULLI` outperformed both PREVENT (AUC: 0.667) and PCE (AUC: 0.683) (Figure 5D), with performance in sex-based analyses (males: `ALADYNOULLI` 0.711 vs PREVENT 0.615; females: `ALADYNOULLI` 0.710 vs PREVENT 0.665, **Figure S8**). We also evaluated the GAIL model (*40*) for breast cancer, using the reported family history data for comparison in the UKB. Of note, many disease-specific clinical scores require information not available on biobank level interviews, though the UK Biobank did provide these variables. `ALADYNOULLI` compared to the GAIL model showed 1-year AUCs of 0.782 vs 0.549 (difference +0.233) for women only (**Figure S19**).

We then specifically evaluated `ALADYNOULLI`’s performance in patients with pre-existing rheumatoid arthritis (RA) and breast cancer (BC) for 10-year ASCVD risk prediction. This analysis investigates whether `ALADYNOULLI` maintains predictive accuracy in the presence of confounding comorbidities that can mask cardiovascular risk signals, a common challenge in clinical practice. For 10-year ASCVD outcomes, we used the static version of our leakage-free prediction approach to compute baseline risk for each individual, as the number of ASCVD events per year in these high-risk subgroups was too small to allow stable estimation of dynamic 1-year AUCs (see Methods). `ALADYNOULLI` achieved AUCs of 0.694 (RA) and 0.689 (BC) for 10-year ASCVD prediction in these high-risk subgroups, outperforming PREVENT (RA: 0.659, BC: 0.544) in these high risk populations where traditional models often underperform, portrayed also in distinct signature trajectories (**Figure S33**).

#### Temporal leakage assessment

We conducted three sensitivity analyses to evaluate the robustness of our predictions to temporal misalignment and diagnostic cascades.

##### (1) Reverse causation assessment

We excluded events occurring within 1, 3, or 6 months before enrollment from predictions at enrollment. This investigates whether predictions rely on diagnostic cascades immediately preceding enrollment rather than genuine predictive signals. Performance remained essentially unchanged across both time horizons: for ASCVD, 1-year prediction AUC was 0.870 at baseline and 0.862 with 6-month exclusion (drop of 0.9%); 10-year prediction AUC was 0.726 at baseline and 0.726 with 6-month exclusion (no measurable drop). The negligible impact of excluding pre-enrollment events indicates that diagnostic cascades are not driving our predictions (see extended data Washout horizons; main text Figure 5d).

##### (2) Time horizon analysis

We compared 10-year predictions at enrollment with and without excluding the first year of outcomes from evaluation. This demonstrated substantial residual predictive power after removing early events that could reflect diagnostic workup effects (ASCVD AUC = 0.723 with 1-year exclusion vs 0.733 baseline, a drop of only 1.4%; see extended data Washout horizons).

##### (3) Washout period analysis across timepoints

We evaluated 1-year predictions at fixed timepoints (enrollment+1 through enrollment+9 years) with varying washout periods (0, 1, or 2 years). At each timepoint, models were trained using only data available up to the prediction time minus the washout period, providing a comprehensive assessment of washout effects across the full follow-up window (see extended data Washout Pivot and Figure S30).

## Discussion

We presented `ALADYNOULLI`, a novel Bayesian framework for modeling dynamic disease signatures and individual health trajectories from longitudinal health records and germline genetic data. By integrating these two data modalities, `ALADYNOULLI` provides a unified framework for understanding disease comorbidities, predicting future disease events, and discovering genetic architecture underlying complex phenotypes. This work addresses a critical gap in precision medicine (*41–43*), where the integration of diverse data sources remains challenging despite the promise of personalized approaches to disease management (*44*). Unlike traditional disease-specific predictive models that require separate development for each condition, `ALADYNOULLI`’s unified framework simultaneously captures risk for multiple diseases, enabling information-sharing across related conditions, improved prediction for diseases with sparse data, and comprehensive decision support across clinical disciplines. Importantly, `ALADYNOULLI` achieves this using only routinely collected diagnostic codes from standard clinical care in the EHR, without requiring the specialized laboratory values, biomarkers, or questionnaire data that established risk scores (PCE, PREVENT, GAIL) depend upon. This implies that future work combining `ALADYNOULLI` with additional data sources can further improve its predictive ability in specific domains.

Our model’s identification of consistent disease signatures across three independent cohorts supports their biological validity and clinical relevance. These signatures capture meaningful disease relationships that align with known pathophysiological processes while highlighting connections between conditions that may share underlying mechanisms. The temporal dynamics of these signatures further enhance our understanding of how disease risk evolves throughout the life course, addressing the need for more sophisticated approaches to understanding disease progression beyond static risk assessment (*45*).

The genetic discovery through GWAS and RVAS analyses represents a significant advance. The identification of genetic variants that associate more strongly with signature loadings than with individual diseases demonstrates that our approach can uncover pleiotropic mechanisms with weaker effects on individual diagnoses—these may represent biologically important pathways and potential targets for therapeutic interventions that are difficult to detect with traditional single-disease GWAS approaches.

Beyond risk prediction, `ALADYNOULLI`’s signature-based framework provides unique capabilities that distinguish it from traditional machine learning approaches. The interpretable, time-varying signature loadings serve as biological patient descriptors that enable fundamentally new types of analyses and applications.

### Signature-based patient matching and stratification

Unlike black-box models that provide only risk scores, `ALADYNOULLI`’s signature loadings enable patient matching based on shared biological mechanisms rather than disease labels. Patients with similar signature profiles, even if they have different diagnoses, may share underlying pathophysiological pathways and could benefit from similar interventions. This signature-based stratification addresses a critical limitation of traditional diagnostic categories, which often mask underlying biological heterogeneity (*46*). For example, individuals with strong metabolic signature contributions to their coronary disease may benefit more from intensive glucose management, while those with inflammatory signature patterns might respond better to anti-inflammatory approaches, regardless of their specific cardiovascular diagnosis.

### Dynamic biological profiling

The model’s ability to detect changing signature profiles in real-time as patients accumulate new diagnoses enables dynamic adjustment of preventive strategies. **Figure 3A–C** demonstrates this capability, showing how patients’ signature loadings are updated following new clinical information, providing a continuously evolving biological profile that reflects the patient’s current health state. This dynamic approach aligns with the emerging paradigm of digital medicine (*47, 48*), where continuous monitoring and real-time biological assessment enable more responsive and personalized care. Unlike machine learning approaches that require retraining to incorporate new information, `ALADYNOULLI`’s signature framework naturally updates patient descriptors as new diagnoses accumulate.

### Enhanced clinical trial design

Signature-based patient stratification has strong potential to enhance clinical trial efficiency by identifying more homogeneous patient populations and more appropriate controls. By enrolling patients with similar signature profiles, trials might achieve greater treatment effects and identify responder subgroups more effectively. This approach could mitigate the high failure rates in clinical trials by ensuring more biologically appropriate study populations (*49*), while also advancing our understanding of treatment response heterogeneity. The cross-cohort consistency we observed in signature-based patient heterogeneity further suggests that signature profiles could serve as generalizable biomarkers for patient stratification across diverse populations and healthcare settings.

Our disease-based approach fundamentally differs from approaches such as Delphi-2M ((*50*)) and (**Figure S26**) that predict individual ICD codes without aggregating to clinically meaningful disease entities. Our model predicts interpretable diseases defined through Phecode aggregation (**Figure S24**). This principled comparison—using the same Phecode-to-ICD mappings that define our disease phenotypes—reveals that disease-level predictions provide superior clinical interpretability while maintaining competitive predictive performance. By predicting diseases rather than codes, our model enables direct clinical translation: physicians can understand and act on predictions for “myocardial infarction” or “colorectal cancer” rather than interpreting abstract ICD code probabilities that may not correspond to meaningful clinical entities. Delphi-2M model parameters are not publicly available for external validation, precluding direct model comparison or individual-level prediction replication. Our comparison is therefore based exclusively on the published performance metrics reported by Shmatko et al. (*50*), using careful Phecode-to-ICD-10 mapping to ensure fair comparison between our disease-level predictions and Delphi’s ICD code-level predictions.

### Comparison with transformer-based foundation models

In addition to differences in phenotypes, our model provides interpretable latent dimensions for both prediction and discovery of novel genetic effects without sacrificing explanatory power. Recent transformer-based foundation models for EHRs have demonstrated impressive scalability and zero-shot prediction capabilities through large-scale pre-training on EHR sequences (*50, 51*). While these approaches excel at general-purpose prediction tasks, `ALADYNOULLI` provides complementary advantages that address different clinical needs. Unlike foundation models that require extensive fine-tuning for new cohorts and lack interpretable biological structure, `ALADYNOULLI`’s signaturebased framework enables fast transfer learning by fixing population-level parameters (*ϕ*, *ψ*) learned from large cohorts and fitting only individual-level signature loadings (*λ*) for new patients—a computationally efficient approach that preserves interpretability. More importantly, `ALADYNOULLI` uniquely combines strong predictive performance with biological discovery capabilities: the interpretable signature structure enables direct genetic discovery (151 GWAS loci, rare variant associations), biological validation (FH, CHIP enrichment), and uncertainty quantification through full Bayesian posteriors—features that foundation models’ black-box architectures cannot provide. While foundation models excel at large-scale general-purpose prediction, `ALADYNOULLI` offers a unified framework for both discovery and prediction, enabling clinicians to not only predict risk but also understand the underlying biological mechanisms driving individual trajectories, making it uniquely suited for precision medicine applications where interpretability and biological insight are paramount.

### Natural handling of competing risks

A critical advantage of our signature-based framework is its natural accommodation of risks for 348 diseases simultaneously. Patients remain at risk for remaining conditions after developing one, and hazards adjust accordingly. This approach reflects clinical reality where patients can and do develop multiple serious conditions sequentially, and the model continues to provide actionable risk estimates for subsequent diseases even after initial diagnoses. For example, a patient who develops myocardial infarction at age 54 remains at risk for heart failure, stroke, and other cardiovascular complications, with the model updating their signature loadings to reflect the new clinical information while maintaining predictions for all other diseases (**Figure S21**).

Several limitations should be acknowledged. First, our model relies on EHR data, which may contain biases related to healthcare access, diagnostic coding practices, and incomplete capture of disease history. These limitations are common to all EHR-based studies and highlight the importance of validating findings across multiple healthcare systems, as we have done here. However, a key methodological advantage of our framework is its ability to handle selection bias through inverse probability weighting: the separation of biological disease-signature relationships (*ϕ*) from population composition effects (*λ*/*π*) enables principled bias correction while preserving core biological patterns, a capability that distinguishes our approach from black-box machine learning methods that cannot easily incorporate such corrections (**Figure S20**). Second, while we incorporate genetic factors, we do not explicitly model environmental exposures or lifestyle factors that significantly influence disease risk (*45*). Third, our use of established PRS may miss genetic effects that act directly on signatures but weakly on individual diagnoses, as our signature-based GWAS identified loci not captured by traditional single-disease approaches. Fourth, our model makes several important assumptions including linearity in genetic effects and additivity in signature contributions, which may not capture all complex interactions. Fifth, our model uses age (time since birth) rather than calendar year as the temporal axis, which does not explicitly capture cohort effects (changes in disease prevalence over calendar time). Because UK Biobank participants all enrolled during a narrow window (2006-2010), we are unable to explore cohort effects. Explicit modeling of calendar year trends would represents an important extension for future work. Future work could integrate these additional data sources and relax these assumptions to further enhance predictive performance and biological insight, addressing the complex interplay between genetic and environmental factors that shape the risk of disease (*52*).

Despite these limitations, `ALADYNOULLI` represents a significant advance in longitudinal health modeling with important implications for precision medicine (*41, 42*). By capturing the complex interplay between genetic predisposition and time-varying disease patterns, our approach provides a framework for more personalized risk assessment and potential therapeutic targeting. Our model’s ability to identify meaningful patient subgroups within traditional disease categories, coupled with enhanced genetic discovery power, moves beyond simple risk prediction to provide deeper insights into disease biology and patient heterogeneity. These capabilities could inform more targeted clinical trials and intervention strategies, ultimately leading to more effective personalized prevention and treatment approaches.

As healthcare increasingly moves toward data-driven precision approaches (*43, 44, 47, 53*), a method like `ALADYNOULLI` that can integrate diverse data sources and model complex temporal relationships will become increasingly valuable for improving patient outcomes. The integration of longitudinal EHR data with genetic information represents a powerful approach to understanding disease biology and improving clinical decision-making, addressing key challenges in modern medicine including the need for more accurate risk prediction, better patient stratification, and enhanced therapeutic targeting (*46*). This work contributes to the broader vision of precision medicine where individual biological profiles guide clinical decision-making, moving beyond the limitations of traditional diagnostic categories toward more nuanced and personalized approaches to disease management.

## Materials and Methods

### Model

We recapitulate the model’s formulation and provide a full account of model assumptions and implementation details.

#### Mathematical Formulation

The `ALADYNOULLI` model represents the probability of disease occurrence for patient *i*, disease *d*, at time *t* via a mixture of probabilities as follows:

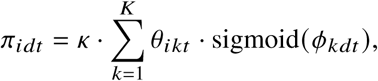

where *κ* is a global calibration parameter, *θ_ikt_* represents patient *i*’s time-varying association with signature *k*, and *ϕ_kdt_* captures the relationship between signature *k* and disease *d* over time.

The patient-signature associations *θ_ikt_* are parameterized as a softmax function of latent variables *λ_ikt_* as:

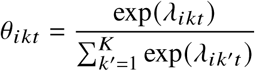

The `ALADYNOULLI` model is specified hierarchically, via distributional assumptions on many of these components, which we refer to as priors.

Indicating by *λ_ik_* the function of time describing the evolution of the latent variable over time for patient *i* and signature *k*, we specify this as a Gaussian process:

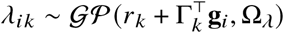

where *r_k_* is a signature-specific baseline, Γ*_k_* captures how genetic/demographic factors **g***_i_* influence patientsignature associations, and Ω*_λ_* is a kernel function controlling temporal smoothness. The covariate matrix **G** contains 36 polygenic risk scores plus sex and 10 genetic PCS (47 features total), providing genetic and demographic information for each individual. The kernel function Ω*_λ_* is defined as:

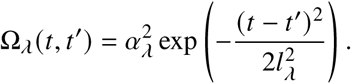

In our implementation, the amplitude parameter *α_λ_* is set to 100 and the length-scale parameter *l_λ_* is set to *T* /4. Similarly, the disease-signature association temporal functions follow a Gaussian process:

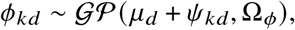

where *μ_d_* is a disease-specific baseline derived from the logit of the population prevalence, *ψ_kd_* represents the the time invariant component of the association between signature *k* and disease *d*, and Ω*_ϕ_* is a kernel function defined as:

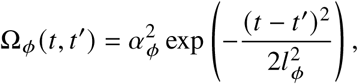

where the amplitude is fixed to *α_ϕ_* = 100 and the length-scale is set to *l_ϕ_* = *T* /3 in our implementation.

The genetic effect parameters Γ*_k_* form a vector of length *P* representing the effects of *P* genetic, sex and ancestral covariates on signature *k*, and are assigned independent zero-mean Gaussian priors:

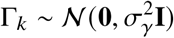

where *σ_γ_*^2^ is the prior variance. This prior encodes the assumption that genetic effects are expected to be small in magnitude.

#### Censored Data and Detailed E Matrix Formation Methodology

A critical aspect of `ALADYNOULLI` is its careful handling of censored observations, which is part of what allows it to function as a **generative model of disease progression** rather than a retrospective analysis tool. The likelihood function, defined below, incorporates time-to-event information through a standard survival analysis encoding of binary disease-specific event indicators in *Y* and censoring times in *E*. Element *E_id_* in *E* represents, in summary, the minimum of the disease-specific event time and the patient’s maximum follow-up time, ensuring the likelihood only includes timepoints where the patient was actually observed. This formulation properly accounts for competing risks by using censoring times derived from actual data.

More specifically, the matrix *E* is defined through the following steps:

##### Step 1: Identify maximum follow-up time for each patient

For each patient *i*, we determine their maximum follow-up age from ICD-10 data: max censor*_i_* = max(last diagnosis date*_i_,* administrative censor date*_i_*). This represents the last timepoint at which patient *i* could have been observed, accounting for death, loss to follow-up, emigration, or end of study period.

##### Step 2: Convert to timepoint scale

We convert the maximum censoring age to the model’s timepoint scale: max timepoint*_i_* = max censor*_i_* − 30, where timepoint 0 corresponds to age 30.

##### Step 3: Cap event/censor times in E matrix

For each patient *i*and disease *d*, the *E* matrix entry is as follows:

- If disease *d* is not observed in patient *i*, then *E_id_* = max timepoint*_i_*
- If disease *d* is observed in patient *i*, then *E_id_* is the age of the diagnosis of disease *d*for patient *i* in years, minus 30.

##### Step 4: Apply to prevalence calculation

When computing prevalence at each age *t*, we only include patients who are still at risk:

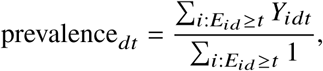

where the denominator includes only patients who have not yet been censored at timepoint *t*. This ensures prevalence estimates reflect realistic at-risk periods.

#### Likelihood Function

The likelihood is constructed to respect this censoring structure (*17, 54*). Consider first ℒ*_N LL_*, the negative log of the likelihood, that is the negative log of the probability of observed disease histories conditional on the disease probabilities *π*’s:

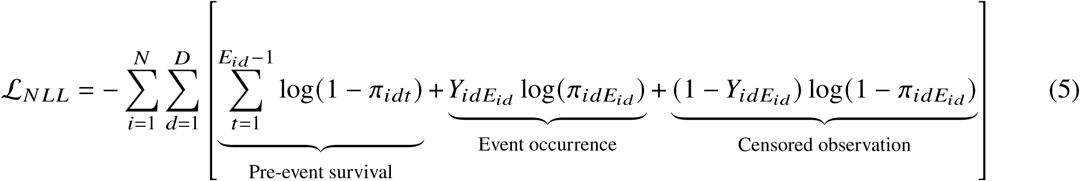

The contribution of individual i to this expression has three components:

1. **Pre-event survival** (*t < E_id_*): For all time points before the event/censoring time, we know that the individual did not have disease *d*, contributing log(1 − *π_idt_*) to the likelihood at each time.
2. **Event occurrence** (*t* = *E_id_*, *Y_idEid_* = 1): If disease *d* occurred at time *E_id_*, we observe the event. The corresponding contribution is log(*π_idEid_*).
3. **Censored observation** (*t* = *E_id_*, *Y_idEid_* = 0): If the individual was censored without disease at time *E_id_*, we only know they were disease-free at that time, contributing log(1 − *π_idEid_*).

#### Objective Function for Maximum a Posteriori (MAP) Computation

The computational derivation of the MAP estimate of the unknown parameters in the model proceeds by optimising the function ℒ*_total_* which includes the negative log-likelihood ℒ*_N LL_* as well as terms arising from the gaussian process prior.

The negative logs of the Gaussian process terms, as previously specified are:

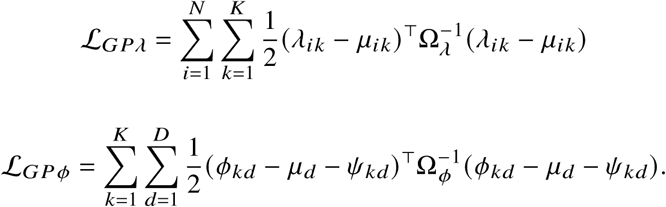

Combining these terms, and assuming diffuse near uniform priors on *κ* and Γ’s, the negative log posterior (the “loss” for short) is:

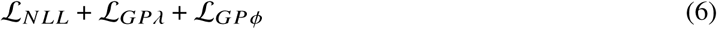

#### Training, Validation and Testing Architecture

The genetic and performance results in this paper are based on training in the UKB dataset; we also trained the model in the MGBB and AOU datasets to establish consistency as in (**2**; model fit for all three data sets including *ϕ* parameters, disease prevalence *μ*and primary *time*− *fixed*signature-disease associations *ψ_k_* are evaluable in exported parameters (Extended data)for all three data sets. Our analytical approach includes two complementary goals: 1) retrospective analysis for model training and cross-cohort validation; 2) prospective analysis for prediction evaluation, assummarized in **Figure S3**.

##### Retrospective Analysis (Figures 2-4)

For computational efficiency and uncertainty quantification, we divided each cohort into non-overlapping subsets: UK Biobank into *B* = 40 subsets of approximately 10,000 individuals each (reserving each subset of 10,000 individuals in turn as a held-out test set), All of Us into 30 subsets, and Mass General Brigham into 4 subsets (reflecting the smaller dataset sizes). For each subset *b* within each cohort, we jointly estimated the disease-signature associations (*ϕ*^^(^*^b^*^)^) and individual loadings (*λ*^^(^*^b^*^)^) using all available observed data up to age 81 or censoring time, whichever came first. This retrospective approach utilized the complete disease trajectory for each individual, enabling us to characterize the full spectrum of disease signatures and their temporal dynamics. For initialization, each subset uses the same pre-computed initial clusters and *ψ* parameters, facilitating consistent signature interpretations across all subsets within each cohort.

For UK Biobank, the final disease-signature parameters used for explanatory analyses were computed as the average across the 40 training subsets (excluding the held-out test set):

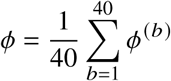

This subset-averaging approach provides robust parameter estimates while maintaining computational tractability. Additionally, it facilitates assessment of estimation robustness. The AOU and MGB cohorts serve as external validation datasets, demonstrating the replicability of disease signatures across different populations and healthcare systems. No prediction tasks were performed on these two cohorts.

This subset-averaged *ϕ*was used to generate the disease signature visualizations in Figure 2, including the temporal patterns and cross-cohort consistency analyses. Individual trajectory analyses (Figure 3) utilized the subset-specific *λ*^(^*^b^*^)^ estimates, and subsequent clustering of patients by their time-averaged signature loadings was performed within each disease category. The genetic analyses (Figure 4) were performed exclusively in UK Biobank, employing the area-under-the-curve of individual signature trajectories as quantitative phenotypes for genome-wide association studies.

##### Prospective Analysis (Figure 5)

For prediction evaluation, we implemented a strictly prospective framework using only UK Biobank data with leave-one-out cross-validation to ensure robust, generalizable performance estimates. We used a subset of 400,000 individuals (the first 400,000 eIDs, which are random, in 40 batches of 10,000 each) from the full UK Biobank cohort of 427,239 individuals for computational efficiency in the cross-validation framework. For each of 40 batches, we trained models on the remaining 39 batches to learn population-level parameters: disease-signature associations (*ϕ*), signature offsets (*ψ*), genetic effects (*γ*), and the global calibration parameter (*κ*). These parameters were then fixed, and predictions for the held-out batch of 10,000 individuals were generated by learning only individual-specific trajectories (*λ̂*) using only data available up to specific prediction time points.

This approach ensures no information leakage from the held-out batch. It also reflects the intended realworld clinical scenarios where population-level disease patterns are known from prior research, but individual risk trajectories must be estimated from available clinical history up to the prediction time. All prediction performance metrics reported in this paper are based on this leave-one-out cross-validation approach.

#### Model Initialization

To ensure computational feasibility and parameter stability we initialize training as follows. We perform the spectral clustering of diagnoses described below and use it to derive starting values for *ψ*. We do this once on the entire data set to establish stable disease clusters and signature-disease associations. These initial *ψ*values are then reused in all subsequent subset analyses for *b*= 1*, . . .,* 40.

Spectral clustering is performed using Scikit on disease co-occurrence patterns. We compute a disease co-occurrence matrix *C* where *C_dd_*′ represents the frequency with which diseases *d* and *d*^′^ co-occur in the same patient. We apply spectral clustering to this matrix to identify *K*disease clusters.

We then initialize the *ψ_kd_* parameters based on cluster membership: for diseases in cluster *k*, we set *ψ_kd_* = 1.0+*ϵ* where *ϵ* is a small random noise. For diseases not in cluster *k*, we set *ψ_kd_* = −2.0+*ϵ*. The choice of initial *ψ* was initialized at values of 1 and −2, which on the log scale correspond to a 20-fold difference in odds (i.e., 10^−9^ vs 10^−11^), thereby spanning a broad range of plausible values for disease risk. For the timevarying parameters *λ_ikt_* and *ϕ_kdt_*, we initialize by drawing a single sample from the corresponding Gaussian process prior with reduced variance to preserve the structured mean initialization. Specifically, we initialize *λ_ikt_* via a random draw from the Gaussian process prior with mean *r_k_* + Γ*_k_*^⊤^**g***_i_* and covariance kernel scaled by amplitude *α* = 0.1, where Γ*_k_* is initialized using regression on disease occurrences. We initialize *ϕ_kdt_* via a random draw from the Gaussian process prior with mean *μ_d_* + *ψ_kd_* and the same reduced amplitude, where *μ_d_* is derived from the logit of disease prevalence. The reduced amplitude ensures that random deviations do not bias the parameters arbitrarily away from the informative mean structure while maintaining temporal smoothness. We generate the GP samples via Cholesky decomposition of the scaled kernel matrices. This approach provides a structured and plausible initialization that reflects our prior smoothness assumptions. The number of latent signatures *K* was chosen to provide a parsimonious balance between model complexity and interpretability, based on prior experience and exploratory analysis.

#### Optimization Details

We trained the model using gradient descent to minimize the loss (6. The solution can be interpreted as approximating a Maximum a Posteriori (MAP) estimate of the parameters, assuming dispersed priors on *λ*’s, Γ’s, *μ*’s *ψ*’s and *ϕ*’s. We minimize the loss (6) over a fixed maximum number of epochs (i.e. complete passes through the entire training dataset). At each epoch, we compute gradients via backpropagation and update parameters using the Adam optimizer in PyTorch, a deep learning framework that allows efficient computation of gradients through automatic differentiation. The model was trained using a learning rate of 0.01. The model is optimized at a time scale of one year and thus trained to provide the most accurate 1-year risk predictions. Longer-term risk (e.g., 10-year) can also be derived by simple manipulations of the estimated *π*’s.

For computational efficiency, we used the Cholesky decomposition to compute the Gaussian process contributions and to sample from the Gaussian process prior during initialization. We also used a jitter term of 1^−6^ to ensure numerical stability when computing the inverse of the kernel matrices. In practice, for our subsets of 10000 individuals, the model converged after 200 epochs. For inference on new patients, the vectorized implementation enables rapid risk prediction: generating predictions for a single individual requires approximately 0.05 second (or 8 minutes for 10000 individuals), making the model suitable for real-time clinical decision support.

#### Computation of Probabilities of Future Events

To ensure that our model provides prospective predictions without data leakage, we implement a strict censoring strategy that distinguishes between cohort recruitment and prediction time. This approach allows us to simulate real-world clinical scenarios where predictions are made based only on information available at a specific point in time (**S10**).

*Cohort enrollment time* refers to the time point when an individual entries `ALADYNOULLI`, which for our purposes is age 30. For example, in our UKB analysis, all individuals were followed in the EHR from 1980 forward (*55, 56*) and thus assigned an enrollment time in our study at young adulthood, age 30, or whichever comes later.

*Cohort recruitment time* refers to the time when individuals joined the Biobank. The UKB recruited individuals aged between 40 and 69 years in the time frame from 2006 to 2010 ((*4*)).

*Prediction time* refers to the time when we imagine making a clinical prediction, with knowledge of the health history up until that time.

*Prediction time for benchmarking with existing clinical risk scores* In practice, when comparing to existing clinical scores, this coincides with recruitment time as above.

In the UKB, an individual is observed in the EHR from adulthood and contributed data to our analysis from age 30 until the end of follow-up. However, for prediction analyses, we only make predictions attime points after the cohort recruitment time.

#### Simulation Study

To validate the ability of the `ALADYNOULLI` model to recover latent disease clusters and temporal dynamics, we conducted a simulation study using `ALADYNOULLI` itself as the generative model. This approach allows us to test whether the model can accurately recover known ground truth parameters from synthetic data that follows the exact same probabilistic structure as our proposed model (**Figure S2**).

We generated synthetic data with *N* = 10, 000 individuals, *D* = 20 diseases, *T* = 50 time points (ages 30-79), *K* = 5 latent disease signatures, and *P*= 5 genetic covariates. The data generation process follows `ALADYNOULLI`’s exact mathematical formulation. We first created *K* = 5 distinct disease clusters with 4 diseases per cluster, assigning strong positive associations within clusters (*ψ_kd_* = 1.0) and strong negative associations outside clusters (*ψ_kd_* = −2.0). Disease baseline trajectories (*μ_d_*) were generated on the logit scale with diverse prevalence patterns ranging from rare (logit prevalence ≈ −14) to common (logit prevalence ≈ −8), incorporating realistic age-dependent onset patterns with varying peak ages and slopes.

For individual trajectories, we generated genetic covariates **G** ∈ ℝ*^N^* ^×^*^P^* and genetic effect matrices Γ*_k_* ∈ ℝ*^P^*^×^*^K^*, then sampled individual signature loadings *λ_ikt_* from Gaussian processes with means **g***_i_*^⊤^ Γ*_k_* and temporal covariance with length scale *T* /4 = 12.5 years. Disease-signature associations *ϕ_kdt_* were sampled from Gaussian processes with means *μ_d_* + *ψ_kd_* and temporal covariance with length scale *T* /3 ≈ 16.7 years. Event probabilities were computed using the exact `ALADYNOULLI`formula: 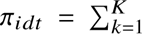 softmax(*λ_ikt_*) · sigmoid(*ϕ_kdt_*), and disease events were sampled from these time-varying probabilities.

When we applied `ALADYNOULLI` to these synthetic data, the model successfully recovered the composition of the signatures (Adjusted Rand Index = 0.843, Normalized Mutual Information = 0.943), and accurately reconstructed the temporal trajectories and genetic effects.

### Cohorts

Data are drawn from three distinct biobanks: Massachusetts General Brigham (MGB), UK Biobank (UKB), and All of Us (AoU). Each cohort is described in **Table S2** and below (**Figure S14**).

#### Massachusetts General Brigham Biobank (MGBB)

MGBB is an integrated research initiative based in Boston, Massachusetts (*18*). It collects biological samples and health data from consenting individuals at Massachusetts General Hospital, Brigham and Women’s Hospital, and local healthcare sites within the MGB network. Since July 1, 2010, the MGBB has enrolled more than 140,000 participants and extracted DNA from approximately 90,000 participants’ samples, and 53,306 participants were genotyped by Illumina Global Screening Array (Illumina, CA). All participants provided their informed written / electronic consent. EHR data are available on all participants from approximately 1990 (see S7). We used a subset of 48,069 for whom EHR and genetic data were available. **Cohort inclusion criteria:** All individuals with available EHR and genetic data were included; no restrictions were placed on the number of visits or age at enrollment, as the model accommodates variable follow-up times through its censoring framework.

#### UK Biobank (UKB)

The UKB is a large-scale, population-based cohort that recruited over 500,000 participants aged 40–69 years between 2006 and 2010 from across the United Kingdom (*4, 57*). The cohort includes extensive phenotypic data, biological samples, and longitudinal follow-up of health outcomes. Genotyping was performed using the UK BiLEVE array or the UKB Axiom array, with subsequent imputation to the Haplotype Reference Consortium (HRC) and UK10K reference panels. Participants were genotyped to investigate genetic contributions to various health and disease traits, with particular attention to the relationship between genetic variants and cardiometabolic diseases. Electronic health records are available on all participants from approximately 1980, and some as early as 1980 (*58*) and thus allow access to clinical diagnostic data prior to the recruitment date). We used the subset of 427,239 for whom genomic and EHR data were available. Polygenic risk scores (PRS) were obtained from an external set of controls (*23*).

#### All of US (AOU)

The AOU research program (*59*) is a large-scale cohort study designed to increase the representation of historically understudied populations in biomedical research. Since 2018, AOU enrolled adults (*age* ≥ 18) at more than 730 US sites. Of the 800,000+ consented participants, more than 560,000 have completed core enrollment requirements, including health questionnaires and biospecimen collection. Data from these participants are continuously linked to electronic health records (EHR), which capture ICD-9 / ICD-10, SNOMED, and CPT codes. Genetic data includes array-based genotyping from 315,000 participants and whole genome sequencing (WGS) from 245,394 participants who were then available to contribute polygenic risk scores for downstream analyzes.

#### Preprocessing and Disease Encoding

Following the approach of Jiang et al. (*8*), we initially analyzed 348 PheCode diseases from UK Biobank that were selected based on prevalence thresholds (≥1,000 occurrences) to ensure sufficient statistical power for comorbidity analysis. This threshold was chosen to balance disease coverage with statistical power: diseases with fewer than 1,000 occurrences would have insufficient power for reliable signature assignment. We focused on ICD-10 chapters A–N (disease chapters), excluding injury (S, T), poisoning, external causes of morbidity and mortality (V–Y), factors influencing health status (Z), and codes for special purposes (U). Disease records were mapped from ICD-10/ICD-10CM codes to PheCodes using a standardized three-step procedure: (1) direct ICD-10 to PheCode mapping using established PheCode definitions (*19*), (2) aggregation of related ICD codes to single PheCode categories, and (3) validation of mapping consistency across cohorts. To validate our findings across independent populations, we then applied the same disease selection strategy to All of Us (AOU) and Mass General Brigham (MGB) cohorts using their respective ICD coding systems. In AOU, we extracted ICD-9 and ICD-10 codes directly from the OMOP Common Data Model condition occurrence tables (*59*), successfully reproducing all 348 diseases from the UK Biobank selection. In MGB, we similarly used ICD-9 and ICD-10 codes, reproducing 346 of the 348 diseases for validation analyses. This multi-cohort approach enabled us to assess the generalizability of disease signatures across different healthcare systems and populations while maintaining consistency in the underlying disease definitions used for `ALADYNOULLI` model development and validation.

### Analysis

#### Stability Across Subsets and Cohorts

We empirically verified that *ϕ* estimates were highly stable across subsets. To assess robustness to natural variation in sample composition, we computed standard errors of *ϕ* parameters across all 40 training batches. Each batch represents 10,000 individuals with slight variation in composition across batches. For each *ϕ*parameter (signature-disease-time combination), we calculated the standard error of the mean as the standard deviation across batches divided by 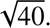 We observed small standard errors (mean SE = 0.0010, median SE = 0.0002, 95% of SE values ≤ 0.004), demonstrating the robustness of our disease-signature associations to batch-to-batch variation in sample composition (**Figure S1**).

To further assess the replicability of our disease signatures across different populations (shown in Figure 2C of the main paper), we performed cluster correspondence as follows (**Fig 2D**). We examined the correspondence between disease clusters identified in each biobank by creating normalized confusion matrices. For each pair of biobanks (UKB vs MGB and UKB vs AoU), we identified the set of diseases common to both biobanks, mapped each disease to its assigned cluster in each biobank based on posterior fitting (disease-to-signature assignments determined by the signature with maximum posterior association strength, argmax*_k_* (*ψ_kd_*)), created a cross-tabulation matrix showing the proportion of diseases in each UKB cluster that were assigned to each MGB/AoU cluster, and normalized the counts by row to show the distribution of cluster assignments.

We computed a composition preservation probability index to quantify cross-cohort correspondence. For each UKB cluster *k*, we identified its best-matching cluster in the comparison cohort (the cluster receiving the highest proportion of diseases from that UKB cluster). The composition preservation probability for cluster *k* is defined as:

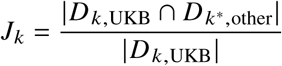

where *D_k,_*_UKB_ is the set of diseases in UKB cluster *k* (determined by posterior *ψ* values, argmax*_k_* (*ψ_kd_*)), *D_k_*∗,_other_ is the set of diseases in the best-matching cluster *k*^∗^ in the comparison cohort (also determined by posterior *ψ* values), and | · | denotes set cardinality. This metric represents the proportion of diseases in a UKB signature that also belong to its best-matching signature in the comparison cohort (i.e., the intersection size divided by the UKB signature size, rather than the traditional Jaccard intersection-over-union). The overall cross-cohort similarity is the median of these cluster-specific similarities: *J* = median(*J*_1_*, J*_2_*, . . ., J_K_*) across all UKB clusters. This metric ranges from 0 (no correspondence) to 1 (perfect correspondence), where higher values indicate stronger replicability of disease clustering patterns across populations.

This analysis revealed strong correspondence between clusters across biobanks (median composition preservation probability = 0.80), particularly for cardiovascular and malignancy signatures, suggesting robust biological patterns that transcend population differences.

For temporal pattern analysis, we performed a detailed comparison of the temporal patterns (*ϕ* trajectories) for diseases shared across all three biobanks, focusing on two key signatures: the cardiovascular signature (MGB: Sig 5, AoU: Sig 16, UKB: Sig 5) and the malignancy signature (MGB: Sig 11, AoU: Sig 11, UKB: Sig 6). For each signature, we identified diseases assigned to that signature in all three biobanks, plotted the temporal patterns (*ϕ* values) for each shared disease, overlaid the average pattern across all three biobanks (gray dashed line, **2**), and used consistent colors for each disease across biobanks to facilitate comparison. This analysis demonstrated consistency in the temporal patterns of disease risk across different populations, with shared diseases showing similar risk trajectories despite being modeled separately in each biobank.

#### Individual Patient Trajectory Visualization

To illustrate the complex interplay of disease signatures in individual patients (shown in Figure 2 **A** of the main paper), we analyzed detailed trajectories for patients with multiple conditions.

For each selected patient, we created a three-panel visualization showing temporal signature loadings, a disease timeline with diagnosis timing, and disease specific probabilities, and then contrasted with the time-averaged signature contributions.

#### Disease-Specific Trajectory and Heterogeneity Analysis

To systematically quantify differences in signature composition among patients with the same clinical diagnosis and understand disease progression heterogeneity and associated genetic architectures (Figures 3F, 4B-D), we performed trajectory clustering analysis using the `ALADYNOULLI` model. For each disease of interest (e.g., breast cancer, major depressive disorder, myocardial infarction), we implemented the following analysis pipeline:

##### Patient Selection and Temporal Averaging

For each disease, we identified all patients who developed the condition and computed their time-averaged normalized signature loadings:

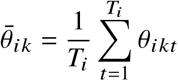

where *θ_ikt_* is the signature loading for individual *i*, signature *k*, and time *t*.

##### Patient Clustering

We applied k-means clustering (k=3) to the time-averaged signature loading matrix *θ̄_ik_* to identify illustrative distinct patient subgroups within each disease category.

##### Trajectory Visualization

We computed cluster-specific mean trajectories across individuals within the cluster 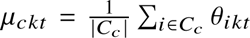 and visualized deviations from population reference as filled line plots for each time point: Δ*_ckt_* = *μ_ckt_* − ref*_kt_*, where ref*_kt_* represents the population-average signature loading.

##### Genetic Architecture Analysis

For each cluster, we computed mean polygenic risk scores (PRS) across individuals in the cluster, and created heatmaps showing cluster-specific values of these scores. To quantify variability of PRS scores among individuals with the same disease, we calculated Cohen’s *d* effect sizes for each PRS comparing in-cluster versus out-of-cluster distributions:

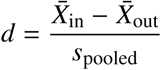

where *X̄*_in_ and *X̄*_out_ are the mean PRS values for patients within and outside each cluster, respectively, and *s*_pooled_ is the pooled standard deviation. Cohen’s *d* values of 0.2, 0.5, and 0.8 correspond to small, medium, and large effect sizes, respectively, providing a standardized measure of genetic differentiation between patient subgroups.

For cluster *c* and signature *k*, *C_ck_*^SIG^ is the standardized difference in mean time-averaged signature loadings between individuals in cluster *c* and those in all other clusters. This measures how distinct each cluster is with regard to each disease signature. Similarly, for cluster *c* and PRS *p*, *C_cp_*^PRS^ is the standardized difference in mean PRS values between individuals in cluster *c*and those in all other clusters.

Confidence intervals and p-values for Cohen’s *d* were estimated, and significance was assessed to determine whether the observed cluster differences were likely to be due to chance.

#### Deviation pathway Heterogeneity Analysis for Myocardial Infarction

As a supplementary analysis, to identify illustrative biological pathways to myocardial infarction using deviation-from-reference clustering, we performed the following supplementary analysis. For each individual with a myocardial infarction diagnosis, we extracted signature loadings (*θ_ik_* (*t*)) over the 10-year window preceding the MI event. We computed signature deviations by subtracting the population reference trajectory for each signature: *δ_ikt_* = *θ_ikt_* −*θ̄_kt_*, where *θ̄_kt_* is the population mean signature loading at age *t*. This deviation-based approach removes age-related confounding and focuses on disease-specific patterns relative to expected population trajectories. For each patient, we computed a 105-dimensional feature vector (21 signatures × 5 timepoints) representing mean signature deviations across the 10-year pre-MI window. Patients were clustered into pathways using K-means clustering (K=4) on these deviation vectors. Pathway assignment was validated through: (1) cross-cohort replication in Mass General Brigham using identical clustering procedures, (2) comparison of precursor disease prevalence patterns across pathways, and (3) signature trajectory visualization to confirm biological interpretability. Cross-cohort validation was performed by applying the UK Biobank cluster centroids to the MGB cohort and comparing pathway sizes and disease enrichment patterns.

#### Genetic Analysis of Signature Trajectories

For each individual *i*, we compute the temporal signature loadings *θ_ik_* (*t*) for each signature *k* and timepoint *t* using the softmax transformation:

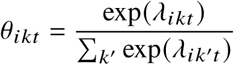

where *λ_ik_* (*t*) is the latent score for individual *i*, signature *k*, and time *t*. The softmax is computed across the signature dimension for each individual and timepoint. To summarize each individual’s overall exposure to a given signature, we integrate the signature trajectory over time:

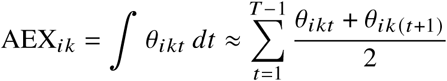

where *T* is the total number of timepoints. The resulting average signature exposure over time (AEX) for each signature is used as a quantitative phenotype for downstream genetic association analysis (**Figure S4**).

We perform GWAS using the AEX values as quantitative phenotypes. For each signature *k*, we test for association between the AEX phenotype and genome-wide SNP genotypes. Association testing is performed using the Regenie (*60*) software (described below), which implements a two-step ridge regression approach for computational efficiency and control of population structure.

##### Genetic ancestry handling

All GWAS analyses included individuals of all genetic ancestries (not restricted to European ancestry), with population structure controlled through inclusion of the first 20 principal components (PCs) of genetic ancestry as covariates. This approach allows us to leverage the full diversity of the UK Biobank while properly accounting for population stratification. The following covariates are included in all association models: sex, age at recruitment, and the first 20 principal components (PCs) of genetic ancestry (*4*).

#### GWAS details

We performed genome-wide association studies using Regenie (*60*), a two-step ridge regression approach designed for computational efficiency and robust control of population structure in large biobank-scale datasets.

##### Step 1: Whole-genome ridge regression

We fit a whole-genome ridge regression model using all genetic variants to account for polygenic effects, relatedness, and population structure. The model was fit using leave-one-chromosome-out (LOCO) cross-validation, where predictions for each chromosome exclude variants from that chromosome to prevent overfitting. This step produces polygenic predictions that capture the aggregate genetic background.

##### Step 2: Single-variant association testing

We performed single-variant association tests on the residuals from Step 1, testing each of 6,418,439 imputed variants (MAF ≥ 0.01, INFO ≥ 0.7) individually. By testing variants on pre-adjusted residuals rather than raw phenotypes, this approach provides well-calibrated association statistics that are robust to case-control imbalance, relatedness, and population stratification. All models included covariates for sex, age at recruitment, and the first 20 principal components of genetic ancestry. Genome-wide significance was set at *P <* 5 × 10^−8^. We identify nearby genes using both nearest gene and gene body analyses (Ensembl and GnomaAD).

In addition to our main analysis featured in 4, for each signature, we identify genome-wide significant SNPs (e.g., *P <* 5 × 10^−8^) and further analyze their relationships with individual disease phenotypes and nearby genes. The analysis proceeds as follows. First, we extract the lead SNPs from the GWAS summary statistics for each signature. For our supplementary analyses (S7), for each top SNP, we test its association with a panel of binary constituent disease phenotypes which comprise our signature inputs using logistic regression, controlling for sex and the first 20 PCs. Third, we visualize the matrix of SNP– phenotype association strength (− log_10_(*P*)) using heatmaps, marking associations significant at *P <* 5×10^−8^. This enables identification of signature-specific loci—SNPs associated with the signature but not with any individual constituent disease. Fourth, we use UpSet plots to visualize the overlap of significant variants across signatures and individual diseases.

#### LDSC Heritability Analysis

Heritability estimates (*ℎ*^2^) represent the proportion of phenotypic variance in signature trajectories that can be attributed to additive genetic variation. Because our signature phenotypes are continuous (integrated area under the signature loading curve), all heritability estimates are on the observed scale, that is, there is no liability-scale transformation for continuous traits.

Heritability estimates for signature trajectories were calculated using linkage disequilibrium score regression (LDSC) (*24*) on the GWAS summary statistics generated from Regenie. For each signature, we applied LDSC with standard parameters using the European ancestry LD reference panel (baseline LD v2.2), following standard practice for heritability estimation from GWAS summary statistics. The LDSC intercept was used to assess potential residual confounding and genomic inflation. For comparison with component diseases, we also performed GWAS for individual cardiovascular traits (myocardial infarction, coronary artery disease, angina pectoris, hypercholesterolemia, coronary atherosclerosis, acute ischemic heart disease, chronic ischemic heart disease, unstable angina) using the same Regenie workflow and covariates (sex, age at recruitment, first 20 genetic PCs), then computed LDSC heritability estimates for each trait. For valid comparison, we report observed-scale heritability for binary traits; liability-scale estimates are provided separately for reference against literature values that typically report on that scale(Table S12).

#### Rare Variant Association Studies (RVAS)

Gene-based rare variant association studies were performed using Regenie (*60*) gene-based association tests. For each signature, we tested associations between signature AEX values and aggregated rare variant burden within genes across six variant filtering masks (Mask1-Mask6), representing progressively inclusive functional impact categories (Mask1: most restrictive, Mask6: most inclusive). Each mask was further stratified by minor allele frequency (MAF) thresholds (singleton, 1×10^−5^, 0.0001, 0.001, 0.01, 0.1). Variants were annotated using Ensembl VEP (Variant Effect Predictor) and aggregated at the gene level as well as Encode using nearest gene. The gene-based test statistic aggregates variant-level association signals within each gene, accounting for linkage disequilibrium and variant effect sizes. Genome-wide significance was set at *P <* 2.5 × 10^−6^ after Bonferroni correction for the number of genes tested (18, 464 genes). Significant genes were identified across all masks to ensure robustness to variant filtering criteria, with genes discovered in multiple masks considered high-confidence associations.

#### Three-Way Validation of Rare Variant Associations

To validate rare variant associations through an independent pathway, we examined the three-way consistency between (1) signature-gene associations from RVAS, (2) gene-disease correlations from rare variant burden analysis, and (3) disease-signature loadings from the *ϕ* parameters. For each signature-gene pair identified through RVAS, we computed rare variant burden scores for each individual by summing variant counts (burden = (2 − *X*), where *X*is the genotype matrix with PLINK encoding. We then calculated Pearson correlations between rare variant burden and binary disease outcomes across all diseases. Finally, we tested whether diseases with high rare variant burden correlations also showed high signature-disease loadings (*ϕ* values) by computing the correlation between the vector of gene-disease correlations and the vector of disease-signature loadings. This three-way validation provides biological plausibility by demonstrating that: (1) rare variant carriers show disease enrichment consistent with signature associations, and (2) signature-disease loadings reflect the same underlying biology captured by rare variant-disease associations.

#### Clinical Validation: Familial Hypercholesterolemia and Clonal Hematopoiesis

To validate the biological meaningfulness of disease signatures, we performed targeted analyses of two well-characterized genetic conditions with known disease associations. **Familial Hypercholesterolemia (FH):** We identified FH carriers using genetic variants in LDLR, APOB, and PCSK9 following established criteria (***?***). For FH carriers and matched controls, we computed signature loadings and tested for enrichment of Signature 5 (cardiovascular signature) in the 5-year window preceding first ASCVD event (myocardial infarction, coronary artery disease, or stroke). For each patient with an ASCVD event, we computed the pre-event change in Signature 5 loading (ΔSig5) over the 5-year window preceding the event, defined as the difference between signature loading at 1 year before the event and at 5 years before the event. We then classified patients as showing a “pre-event rise” if ΔSig5 ≥ 0. Enrichment was quantified by comparing the proportion of FH carriers versus non-carriers who showed a pre-event rise using Fisher’s exact test on a 2×2 contingency table (rise/no-rise × carrier/non-carrier), with one-sided alternative testing for higher enrichment in carriers. Results were reported as odds ratios with p-values from Fisher’s exact test.

#### Model Evaluation and Comparison

Figure 5 presents a comprehensive evaluation of our multi-disease risk prediction model in the UKB, and comparisons with important single-disease models. Each is evaluated in a strictly prospective, leakage-free framework. In the testing data, all individual level parameters were estimated using only information available up to the time of prediction. Individuals with prevalent disease at prediction were excluded from the risk set for that disease. In UKB all individuals are followed for at least 10 years from recruitment. We considered the following prediction tasks and metrics.

##### Median AUC Aladynoulli Dynamic

This metric evaluates the model’s ability to make dynamic predictions at multiple time points during follow-up: it is derived by refitting the Aladynoulli model using fixed *ϕ̂*, *γ*^ and *κ* parameters, previously estimated from the full-history training data, and now applied to a series of one-year prediction tasks. Critically, while the fixed parameters above were estimated from the full training data, the *λ̂*’s for each prediction task are now estimated using data only up to the point of prediction. For each of the first 10 years after recruitment, the model is retrained using only data available up to that point for the held out test set (in orange in **Figure S3**). The median area under the receiver operating curve (AUC) across these ten dynamic one-year fits is reported. This captures how predictive accuracy evolves as patients accumulate new diagnoses and leverages the flexibility of our method to perform dynamic, prospective risk estimation at any time point. Individuals with prevalent disease at prediction time were excluded from the risk set.

##### Aladynoulli Recruitment (1-year)

This metric uses the Aladynoulli model’s predicted 1-year risk at the time of recruitment, evaluated against observed 1-year outcomes. The risk estimate is *π̃_idt_*, the predicted 1-year risk for individual *i* recruited at time *t* for disease *d*. In practice, any age of prediction could be chosen, but we use the age of recruitment to the UK Biobank because it is the only time at which we have the availability of additional clinical variables required for comparability with existing clinical benchmarks.

##### Aladynoulli Recruitment (10-year)

This metric uses the Aladynoulli model’s predicted 1-year risk at the time of recruitment, evaluated against observed 10-year outcomes. The risk estimate is *π̃_idt_*, as in the 1-year predictions.

##### Cox

For benchmarking purposes we employ a ‘baseline Cox model useing only family history and sex as covariates, with age as the time scale delineating start and stop of the interval, representing a minimal clinical model that does not require any model curation or disease-specific features. This highlights the fact that our approach does not rely on disease-specific risk factors or manual feature engineering.

When benchmarking AUC performance, we compared the `ALADYNOULLI` model not only to the benchmarking Cox proportional hazards model above, but also to established clinical risk scores: PREVENT (*61*), Pooled Cohort Equation (*62*) for ASCVD, and Gail (*40*) for breast cancer, models for diseases where these scores are available after specific and often expensive curation. Of note, these clinical risk scores require laboratory values and biomarkers that are either collected during targeted clinical visits (introducing selection bias) or, when extracted from routine EHR data, may be subject to measurement bias since sicker patients typically receive more frequent testing. In contrast, our approach leverages routinely collected diagnostic codes (ICD codes) that are systematically recorded for all patients regardless of disease severity, providing a more unbiased data source for risk prediction.

This approach ensured that all model comparisons were fair, prospective, and reflective of the information available at the time of risk assessment.

##### Dynamic 10-year Rolling

This approach demonstrates the model’s interpolation capabilities by evaluating how probability estimates evolve as new information becomes available. For each year of the 10-year horizon, we update the model’s predictions using information available up to that time point, then aggregate the cumulative 10-year risk as 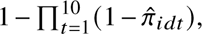 where *π*^*_idt_* is the predicted risk for individual *i* and disease *d* at year *t* after recruitment. While this rolling evaluation does not use knowledge of the future outcome of interest, it is not leakage-free. This is because it does incorporate future information about events that are potentially correlated with, or even resulting from, the event of interest, because the model’s probability estimates at year *t* are influenced by information available up to year *t*. Thus it is best understood as interpolation rather than extrapolation. While this metric cannot be used for prospective evaluation, it demonstrates the model’s technical capabilities for dynamic risk assessment and shows how probability estimates evolve over time.

##### Additional Methodological Challenges

We conducted a series of analyses to assess the robustness of `ALADYNOULLI` to selection bias, population stratification, temporal leakage, and reverse causation, complementing the primary analyses described in the main text. We provide technical details next.

##### Three key methodological features

(1) **Inverse probability weighting (IPW)** to correct for UK Biobank participation bias while preserving biological signal; (2) **Comprehensive washout analyses** (1-6 months and 1-2 years) to assess temporal leakage and reverse causation; and (3) **Patient-specific censoring** using realistic follow-up times to properly handle competing risks. These strengthen the validity and generalizability of our findings.

##### Selection Bias and Inverse Probability Weighting (IPW)

To evaluate robustness to participation bias in UK Biobank, we implemented inverse probability weighting (IPW) following Schoeler et al. ((*38*)). We fit a lasso-regularized logistic regression model for biobank participation using demographic and socioeconomic variables (region, birth year, sex, household characteristics, employment status, housing tenure, ethnicity, self-reported health, and education) and computed each individual’s predicted participation probability *P*^*_i_*.

Inverse probability weights were defined as

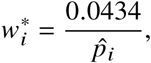

where 4.34% is the estimated participation rate in the source population, with *w_i_*^∗^ trimmed at the 1st and 99th percentiles to avoid undue influence of extreme weights. These raw weights are then normalized to sum to *N*:

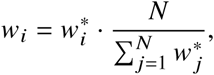

ensuring that the weighted sample size equals the observed sample size for proper loss scaling.

The weighted loss function modifies the data likelihood component of Equation **??** to incorporate individual weights. The weighted negative log-likelihood is:

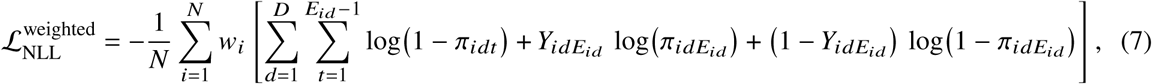

Where *w_i_* is the normalized IPW weight for individual *i*. The Gaussian process prior terms (L_GP_*_λ_* and L_GP_*_ϕ_*) remain unweighted, as they represent priors on model parameters rather than data contributions.

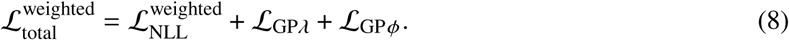

This formulation ensures that individuals with lower predicted participation probabilities (underrepresented groups) contribute proportionally more to parameter estimation, effectively rebalancing the training data toward the source population distribution.

##### Population Stratification and Genetic Ancestry

To assess the impact of population structure on signatures, we performed both PC-adjusted and ancestry-stratified analyses.^1^ First, we compared models fit with versus without genetic principal components in the Gaussian process mean for *λ* (Equation 3), holding all other settings fixed. Disease–signature associations (*ϕ*) were nearly identical between models (correlation = 1.000, mean absolute difference *<* 0.002), demonstrating that disease–signature relationships are robust to PC adjustment. In contrast, individual signature loadings (*θ*) showed ancestry-specific shifts, with cardiovascular Signature 5 exhibiting the largest deviation for South Asian (SAS) individuals, consistent with known elevated cardiovascular risk in this group. Second, we performed PCA on time-averaged signature loadings and examined clustering by ancestry (EUR, AFR, EAS, SAS), finding substantial overlap between groups with modest deviations along principal components for AFR, EAS, and SAS. Finally, we compared `ALADYNOULLI` predictions to Cox baseline models including age, sex, and genetic PCs, confirming that `ALADYNOULLI` retains a performance advantage across ancestry groups, indicating that its gains are not explained by ancestry covariates alone (Extended Data Figure S31).

##### Reverse Causation Assessment: Short-Term Washout

To test whether predictions are driven by diagnostic cascades immediately preceding events, we excluded events occurring within 1, 3, and 6 months of enrollment.^2^ Performance remained robust with minimal AUC degradation (mean drop = 0.0112 for 1-year predictions, 0.0010 for 10-year predictions), indicating that `ALADYNOULLI` captures predictive signal rather than reverse causation effects.

##### Temporal Leakage Assessment: Multi-Year Washout

To address temporal misalignment between ICD-10 dates and true disease onset, we conducted two complementary analyses. First, we compared 10-year predictions made at enrollment with versus without excluding the first year of outcomes following enrollment. This analysis (Panel B) demonstrates robustness to excluding the first year of outcomes, with minimal performance degradation (e.g., ASCVD AUC = 0.723 with 1-year exclusion vs 0.733 baseline). Second, we evaluated 1-year predictions at fixed timepoints (enrollment+1 through enrollment+9 years) using models trained with different washout periods (0, 1, or 2 years). For each prediction timepoint *t_Pred_* ∈ {1, 2*, …,* 9}, we trained models using data up to *t_Pred_* − *w* years, where *w* ∈ {0, 1, 2} represents the washout period. This analysis (Panel C) tests for temporal information leakage by varying the amount of training data available, demonstrating substantial residual predictive power even with 1–2-year washout periods across key diseases.

All analyses were performed using Python, with survival models implemented in lifelines and scikitsurvival, and validated in R (Version 4.0) using the Survival package, and calibration and discrimination metrics computed using standard epidemiological methods.

## Data Availability

All data produced in the present study are available upon reasonable request to the authors.

http://aladynoulli.hms.harvard.edu

## Author contributions

S.M.U., P.N. and G.P. conceptualized the study, developed the methodology, implemented the software, and wrote the original draft. Y.D. and X.J. contributed to methodology development and formal analysis. W.H. assisted with data curation and visualization. A.G., P.N., and G.P. provided supervision, resources, and critical review. All authors contributed to manuscript review and editing.

## Data and materials availability

All code for implementing `ALADYNOULLI` is publicly available at https://surbut.github.io/aladynoulli2/index.html, with comprehensive documentation and interactive analysis notebooks available at https://surbut.github.io/aladynoulli2/index.html and https://surbut.github.io/aladynoulli2/reviewer_responses/README.html.

1. **Complete code repository:** All model implementation, training scripts, and analysis code are publicly available at https://github.com/surbut/aladynoulli2 and outlined in detail at https://surbut.github.io/aladynoulli2/index.html#complete-workflow. This includes:
  - Detailed description of model implementation and discovery vs prediction overview: https://surbut.github.io/aladynoulli2/reviewer_responses/notebooks/framework/Discovery_Prediction_Framework_Overview.html
  - Suggested workflow: https://github.com/surbut/aladynoulli2/blob/main/pyScripts/dec_6_revision/new_notebooks/reviewer_responses/preprocessing/WORKFLOW.md
  - Detailed explanation of preprocessing: https://surbut.github.io/aladynoulli2/reviewer_responses/preprocessing/create_preprocessing_files.html
  - Training scripts for all three cohorts: https://github.com/surbut/aladynoulli2/blob/main/claudefile/run_aladyn_batch_vector_e_censor.py
  - Prediction and validation pipelines: https://github.com/surbut/aladynoulli2/blob/main/claudefile/run_aladyn_predict_with_master_vector_cenosrE_fullEtest.py
  - All analysis notebooks addressing reviewer concerns: https://surbut.github.io/aladynoulli2/index.html#reviewer-response-analyses
  - Vectorized real-time patient refitting implementation
2. **GWAS summary statistics:** Full summary statistics for all 21 signatures (SIG0-SIG20) will be made available via Zenodo (DOI to be assigned upon acceptance). Each file includes: SNP rsID, chromosome, position, effect allele, other allele, beta, standard error, p-value, minor allele frequency, and sample size. These summary statistics enable independent replication, meta-analysis, and genetic correlation analyses without requiring access to individual-level genetic data. Additionally provided:
  - Lead SNP summary statistics for each signature (SIG0-SIG20) with genome-wide significant associations
  - Variant annotation comparison file (variant annotation comparison.csv) containing annotations for all 151 GWAS hits with gene assignments, locus boundaries, and validation vs. novelty classifications
3. RVAS and genetic association results:
  - Canonical gene-signature associations across 6 variant filtering masks (canonical/ directory with 36 .tsv files)
  - PRS-signature association results (gamma prs.csv) with effect sizes, standard errors, and p-values for all 756 PRS-signature tests
  - Cohen’s d values for signature differences across patient heterogeneity clusters (signature cohens d *.csv for myocardial infarction, breast cancer, and major depressive disorder)
  - PRS Cohen’s d values across heterogeneity groups (prs cohens d *.csv) demonstrating genetic validation of patient subgroups
4. Trained model parameters: Due to UK Biobank data sharing policies, full trained model parameter matrices (*ϕ*, *ψ*) derived from individual-level data cannot be directly shared. However, we provide aggregated results that enable replication and extension:
  - Disease-signature association tables showing signature-disease relationships (aggregated *ϕ*values as tables/figures)
  - Age-specific prevalence estimates for all diseases across all three cohorts (aggregated population-level data)
  - Disease-signature loading matrices (*ψ_kd_*) showing which diseases load strongly on each signature (signature compositions, aggregated)
  - Signature temporal patterns (aggregated across population)
  Model parameters for MGB and All of Us cohorts may be shareable subject to their respective data use agreements (to be confirmed upon publication).
5. PheCode lists and disease mappings:
  - Complete list of 348 diseases with PheCode definitions (PheCode version 1.2)
  - ICD-10 to PheCode mappings used in our analysis
  - Disease prevalence estimates across all three cohorts
  - Available in CSV format in the GitHub repository (data/ directory)
  - Detailed aggregation analysis showing how PheCodes aggregate ICD-10 codes (see https://surbut.github.io/aladynoulli2/reviewer_responses/notebooks/R1/R1_Q3_ICD_vs_PheCode_Comparison.html for full analysis demonstrating 15x dimensionality reduction) and in the supplementary text of our main paper.
6. **Interactive results website:** http://aladynoulli.hms.harvard.edu provides interactive visualizations of:
  - Signature compositions and temporal patterns
  - Individual trajectory examples
  - Genetic loading examples
  - Individual risk calculation examples
  The website displays only synthetic data (slightly modified UK Biobank data for individual examples) and does not provide access to individual-level model parameters or allow reverse engineering of the UK Biobank model parameter matrices, which remain protected under data sharing policies.
7. **Permanent archive:** A versioned release of all code, summary statistics, and supplementary data files will be archived on Zenodo (DOI to be assigned upon acceptance) to ensure long-term accessibility.

**Biobank data access:** Individual-level data from UK Biobank, Mass General Brigham Biobank, and All of Us are available through their respective access procedures due to data use agreements and patient privacy protections. However, our publicly shared summary statistics, aggregated results, and code enable researchers to apply our methods to other cohorts and conduct meta-analyses without requiring access to the original biobank data.

We have updated the Data Availability statement in the manuscript to comprehensively reflect these commitments. All publicly shared resources contain only aggregated or de-identified information in compliance with institutional IRB approvals and data use agreements.

## Extended Data

### Extended Data Files

- **Exported parameters:** Population-level parameters (*ϕ*, *ψ*, *μ*) for UK Biobank, All of Us, and Mass General Brigham cohorts. Per cohort subdirectory (ukb/, aou/, mgb/):
  – phi master pooled.npy - Time-varying disease-signature associations (21 signatures × 348 diseases × 52 timepoints)
  – psi master.npy - Static signature-disease strength matrix (21 × 348)
  – mu baselines.npy - Disease baseline prevalence trajectories (348 × 52, UK Biobank only)
  – Summary tables (top diseases per signature, top signature per disease, signature statistics)
  – Verification plots (heatmaps, trajectory visualizations)
  – Metadata JSON files with parameter descriptions and README files
- **CohensD:** Cohen’s *d*and p-values for signature loading and PRS differences between disease clusters. Six CSV files quantifying which signatures and polygenic risk scores distinguish patient subgroups:
  – `signature cohens d *.csv` - Three files for major depressive disorder, breast cancer, and myocardial infarction signature loading differences
  – prs cohens d *.csv - Three files for PRS differences between disease subtypes for the same three diseases Columns include cluster ID, signature/PRS number, mean values within and outside clusters, Cohen’s *d*effect sizes, and p-values.
- **Annotation:** PRS-signature association statistics and variant annotation files:
  – gamma associations.csv- Effect sizes, standard errors, Z-scores, p-values, and FDR-corrected significance for 756 PRS-signature associations (36 PRS × 21 signatures), with 116 significant at FDR *<* 0.05
  – variant annotation comparison.csv - Comparison of Ensembl VEP vs original nearestgene annotations for all 151 genome-wide significant loci
  – all loci annotated.tsv - Comprehensive annotation of all significant loci with Ensembl VEP annotations
  – SIG5 AUC unique genes 1mbwindow.csv - Unique Signature 5 genes using 1MB window overlap analysis (10 genes)
  – `SIG5 AUC unique genes exact.csv` - Unique Signature 5 genes using exact SNP matching (23 genes, including PDGFD, SCARB1, SMAD3, C1S, HLA-DOB, WWP2, and others)
- **Additional heterogeneity comparison:** Pathway analysis for myocardial infarction heterogeneity and cross-cohort validation:
  – complete pathway analysis output/ - Underlying data for Extended Data Figure S28, including pathway discovery analysis with signature deviation trajectories, PRS patterns, and pathway comparisons (9 PDF visualizations in ukb pathway discovery/), cross-cohort validation comparing UK Biobank to Mass General Brigham (transition analysis visualizations), and text summaries
  – `mgb deviation analysis output/` - Mass General Brigham cohort validation results including statistical test results, visualizations, and pickled results for programmatic access
- **Supplementary tables.xlsx:** Comprehensive performance evaluation results. Multi-sheet workbook consolidating prediction performance comparisons. Each sheet contains AUC values with confidence intervals, event counts, and improvement metrics:
  1. **Washout analysis (short-term):** Comparing different exclusion periods (0, 1 month, 3 months, 6 months) for 1-year and 10-year predictions. Events occurring within the specified period before enrollment are excluded from outcome evaluation, while the model is trained using all available data up to enrollment.
  2. **Age-offset analysis:** Model trained at t=0, t=1, …, t=9 years since enrollment, with 1-year predictions evaluated at each timepoint.
  3. **Washout results by disease pivot:** Models trained at time horizons t = 0, 1, 2, …, 9 years post-enrollment. For each prediction timepoint *t_Pred_* ∈ {1, 2*, …,* 9}, performance evaluated using models trained at horizons *t_train_* = *t_pred_* − *w*, where *w* ∈ {0, 1, 2} represents the washout period in years. This design enables assessment of how prediction accuracy varies with the temporal distance between model training and prediction timepoints, simulating scenarios where predictions must be made using models trained with less recent data.
  4. **Washout results by disease summary:** Summary of 0-, 1-, and 2-year washout predictions over different time horizons for the above analysis.
  5. **Washout analysis (10-year):** Comparing 10-year predictions in which the model is trained with data at the prediction time and the first year of outcomes are excluded from measurement.
  6. **Delphi-2M comparison (PheCode-based mapping):** Comparison of 1-year AUC with Delphi- 2M using PheCode-based mapping (principled comparison using actual PheCode→ICD aggregation).
  7. **Delphi-2M comparison (simple mapping):** Comparison using simple mapping (manual dictionary/string matching approach for baseline comparison).
  8. **ICD-10-CM to PheCode mapping:** Mapping with Delphi and Aladynoulli 1-year predictions for 28 diseases (83 rows).
  9. **Summary of prediction horizons:** Summary of 1-, 5-, and 10-year predictions (static, using prediction at t=0 for 10-year outcomes) and dynamic AUC.
  10. **Baseline predictions:** Results for 1-year prediction at enrollment (0-year washout) for all individuals.
  11. **External clinical scores:** Comparison with external clinical scores (PCE, PREVENT, QRISK3, GAIL).
  12. **Cox proportional hazards baseline:** Comparison with Cox proportional hazards baseline.
  13. **Time horizon comparisons:** Comparisons across 1-, 5-, and 10-year prediction horizons.
  14. **ICD-10-CM to PheCode mapping (complete):** Complete mapping (14,048 rows mapping ICD-10-CM codes to standardized PheCode categories).
- **RVAS:** Gene-based rare variant association study (RVAS) results:
  – `significant/` - Results across 6 variant filtering masks (Mask1-Mask6) with multiple minor allele frequency thresholds per mask (singleton, 1 × 10^−5^, 0.0001, 0.001, 0.01, 0.1), including both all significant results and canonical transcript results (suffix canonical.tsv). Also includes summary CSV tables in gene based csv/ and LaTeX-formatted tables in rvas tables latex.tex
  – ss pub/ - Compressed summary statistics files: twenty-one gzipped files (one per signature) containing variant-level summary statistics used for gene-based RVAS analysis
- **GWAS:** Lead SNPs for each disease signature and Signature 5 component trait analysis:
  – `signature lead snps/Supplementary data S*.txt` - Twenty-one files (one per signature, SIG0-SIG20) containing genome-wide significant lead variant summary statistics. Each file includes chromosome, position, variant ID, effect/other alleles, effect allele frequency, beta, standard error, − log_10_(p-value), sample size, rsID, locus boundaries, and overlap classification (e.g., whether the locus was detected as a lead locus in constituent-trait GWAS within our study). Used for Manhattan plots and genetic discovery analysis.
  – signature5 componenttrait lead snps/ - Lead SNPs from constituent cardiovascular trait GWAS used for Signature 5 uniqueness analysis, containing lead variant summary statistics for 6 cardiovascular traits (angina pectoris, coronary atherosclerosis, hypercholesterolemia, myocardial infarction, and other ischemic heart disease variants) and visualization of variant overlap (cardiovascular variants upset plot 1mb windows.pdf)

**Figure S1:**
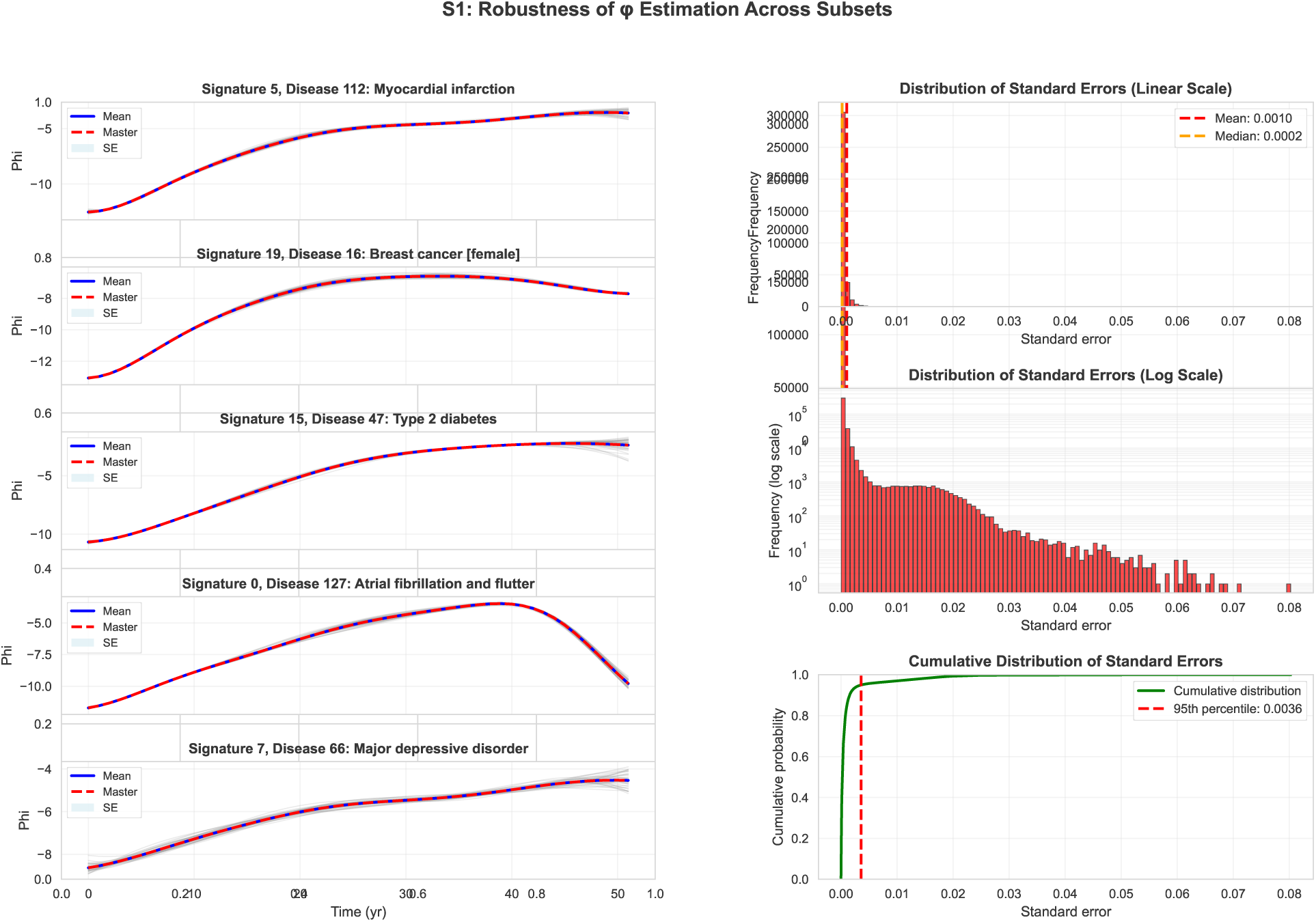
Robustness of *ϕ* estimation across subsets. (**A**) Phi trajectories for five representative diseases across all 40 training subsets. For each disease-signature pair (myocardial infarction–Signature 5, breast cancer–Signature 19, type 2 diabetes–Signature 15, atrial fibrillation–Signature 0, depression–Signature 7), the plot shows trajectories from all subsets (gray lines), the mean across subsets (blue line), the master checkpoint estimate (red dashed line), and standard error bands (light blue shading). (**B**) Distribution of standard errors across all *ϕ* parameters: (top) histogram on linear scale with mean and median indicated, (middle) histogram on log scale, (bottom) cumulative distribution with 95th percentile marked. The very small standard errors (mean SE = 0.0010, median SE = 0.0002, 95% of values ≤ 0.004) demonstrate the stability of *ϕ*estimation across subsets and confirm that disease signatures represent replicable biological patterns rather than subset-specific noise.

**Figure S2:**
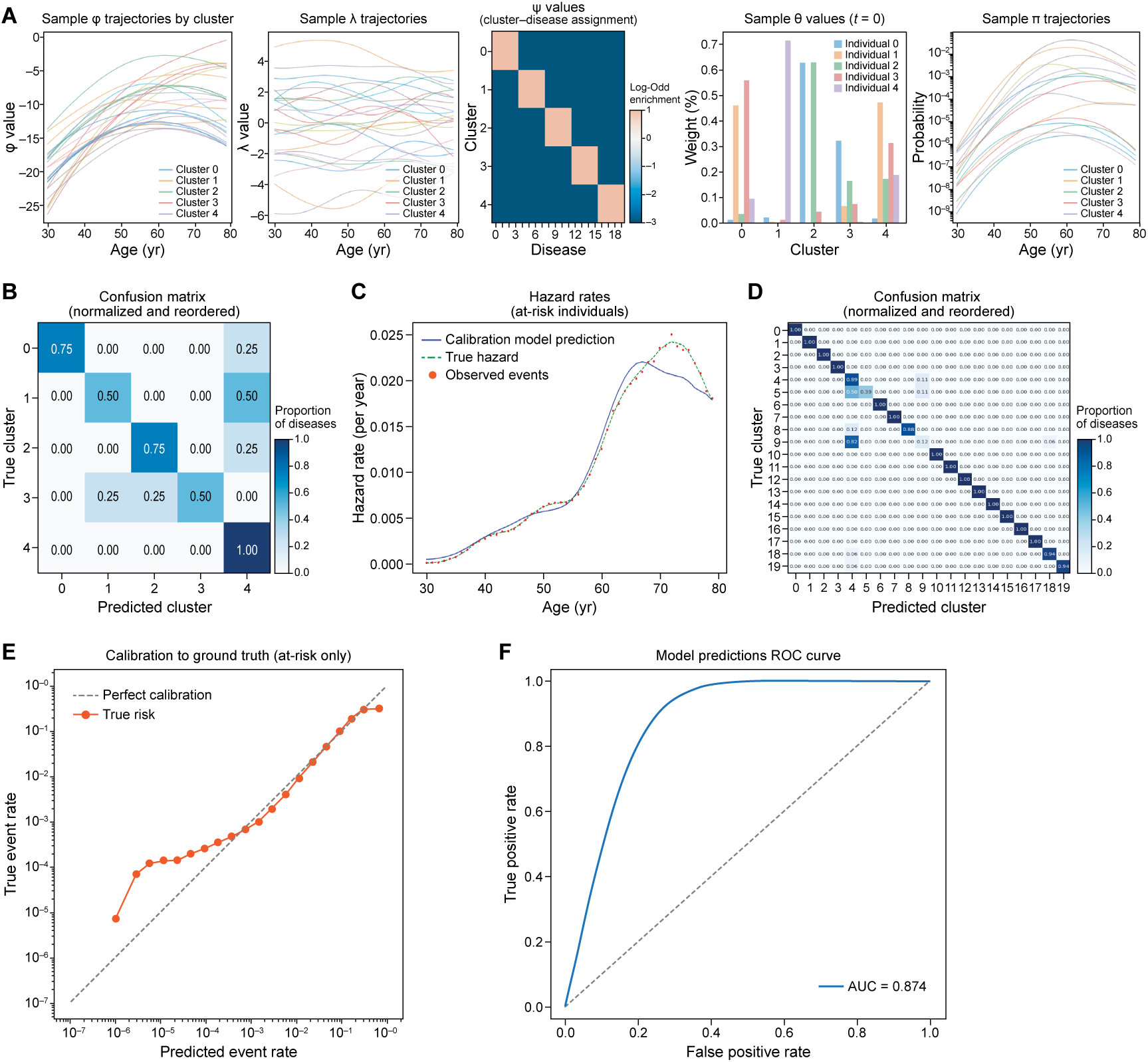
Simulation study demonstrates accurate recovery of latent disease clusters and temporal dynamics. (**A**) Simulated disease baseline trajectories on the logit scale, showing diverse prevalence and onset patterns. (**B**) Example latent signature trajectories for individual patients, illustrating temporal smoothness and genetic heterogeneity. (**C**) True and inferred disease cluster assignments visualized as a confusion matrix. (**D**) Correlation matrix comparing true and inferred cluster assignments. (**E**) Comparison of true and model-inferred hazard rates over time. (**F**) ROC curve for simulated disease prediction, demonstrating high discriminative performance. Together, these results confirm that the `ALADYNOULLI` model can accurately recover both the cluster structure and temporal risk dynamics from complex, realistic simulated data.

**Figure S3:**
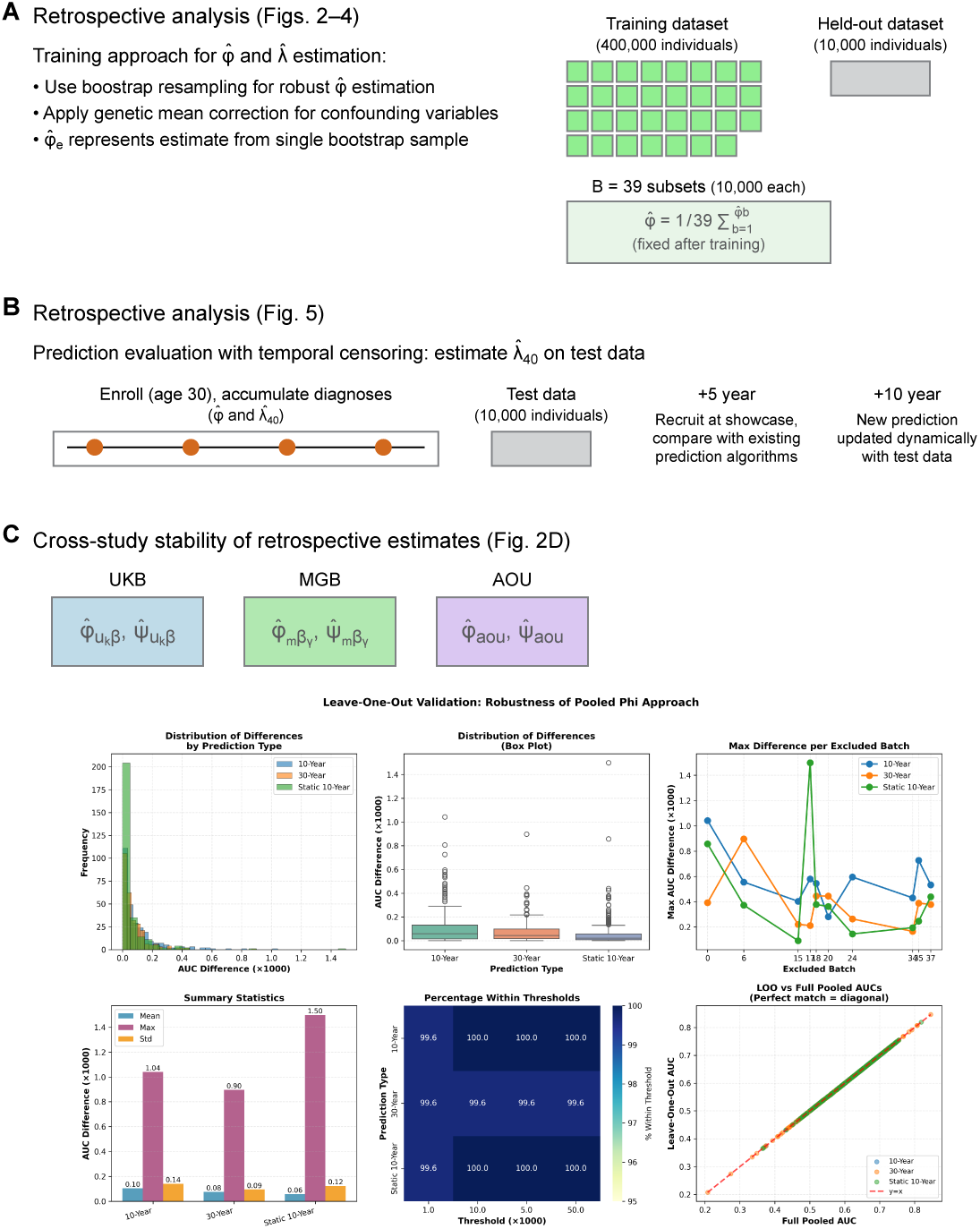
ALADYNOULLI. (**A**) *Joint analysis (Figures 2-4):* UK Biobank training data (400k individuals) divided into 40 subsets of 10k each, with one subset held out for testing. Disease-signature associations (*ϕ̂*) and individual loadings (*λ̂*) estimated on each of the 39 training subsets using complete disease trajectories, then averaged: 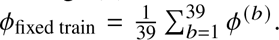 The held-out test set is never used for *ϕ* estimation, ensuring no data leakage. Genetic validation performed using models with genetic mean effects removed (Γ*_k_* = 0). (**B**) *Prospective Analysis:* Rigorous prediction evaluation using the held-out test set (10k individuals) with fixed *ϕ*_fixed train_ parameters. Individual loadings (*λ̂*_test_) re-estimated using only data available up to each prediction time point through temporal censoring. This approach prevents data leakage and simulates real-world clinical scenarios where population-level disease patterns are known but individual risk trajectories must be estimated prospectively from available clinical history. (**C**) *Cross-Population Validation:* Independent estimation of disease signatures in Mass General Brigham (MGB) and All of Us (AOU) cohorts demonstrates reproducibility across different populations and healthcare systems. Each cohort yields cohort-specific *ϕ* and *ψ* parameters, with strong correlation between cohorts confirming biological validity rather than population-specific artifacts. This cross-population validation strengthens confidence in the universal applicability of discovered disease signatures (see Figure S5). (**D**) *Leave-One-Out Cross-Validation:* Robustness validation of the pooled *ϕ*approach by excluding one batch at a time and comparing predictions using leave-one-out *ϕ* estimates versus full pooled *ϕ* estimates. The analysis evaluates static 10-year and dynamic 10-year AUC comparisons across all 40 batches. Results demonstrate excellent robustness: predictions showed near-perfect correlation (r = 0.995) with master predictions, mean AUC differences were ≤ 0.0001, maximum differences were ≤ 0.0015, and 99.6-100% of comparisons fell within a 0.01 AUC threshold. The scatter plot (bottom-right) shows LOO AUCs versus full pooled AUCs clustering tightly along the y=x line, confirming that excluding any single batch has negligible impact on predictive performance. This validation confirms that no single batch dominates the pooled *ϕ* estimates and that the full pooled approach generalizes well across all batches.

**Figure S4:**
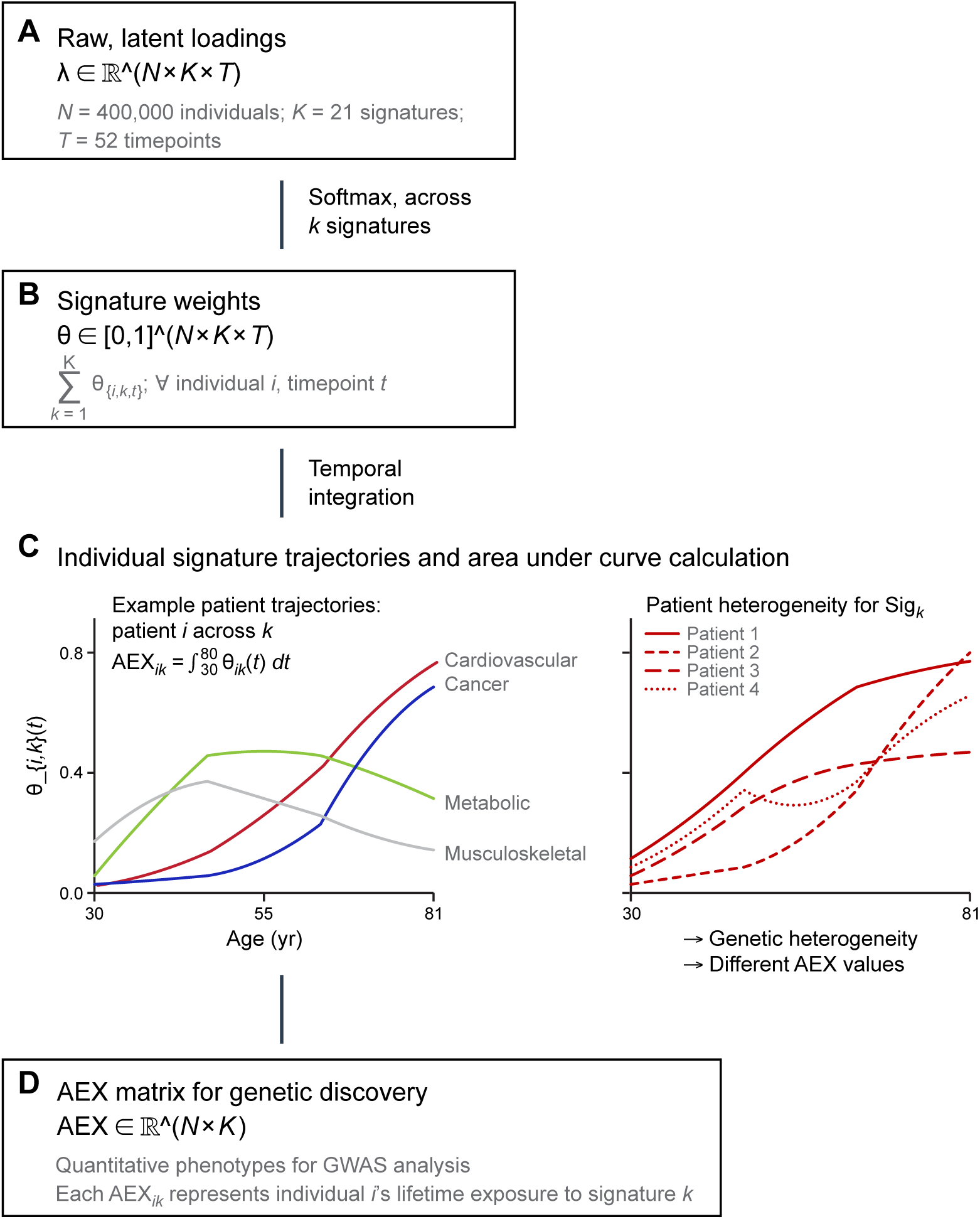
Average Exposure over time (AEX) calculation for genetic discovery. (**A**) *Raw Latent Loadings:* The model estimates individual-specific loadings *λ̂* ∈ ℝ*^N^* ^×^*^K^*^×^*^T^* for N=400,000 individuals across K=21 signatures and T=52 timepoints (ages 30-81). (**B**) *Signature Weights:* Raw loadings are transformed via softmax to obtain normalized signature loadings *θ*0, 1 *^N^* ^×^*^K^* ^×^*^T^*, where *_k_ θ_ik_ t* = 1 for each individual and timepoint, representing the probability distribution across signatures. (**C**) *Individual Signature Trajectories:* Left panel shows example temporal trajectories for different signatures (cardiovascular, cancer, metabolic, musculoskeletal) illustrating distinct age-related patterns. Right panel demonstrates patient heterogeneity within a single signature, showing how genetic and environmental factors lead to different AEX values across individuals. (**D**) *AEX Matrix:* The area under each individual’s signature trajectory is computed as AEX*_ik_* = ^80^ *θ_ik_ t dt*, yielding a quantitative phenotype matrix AEX ℝ*^N^* ^×^*^K^* where each entry represents individual *i*’s lifetime exposure to signature *k*. This matrix serves as the input for genome-wide association studies to identify genetic variants influencing signature-specific disease risk patterns.

**Figure S5:**
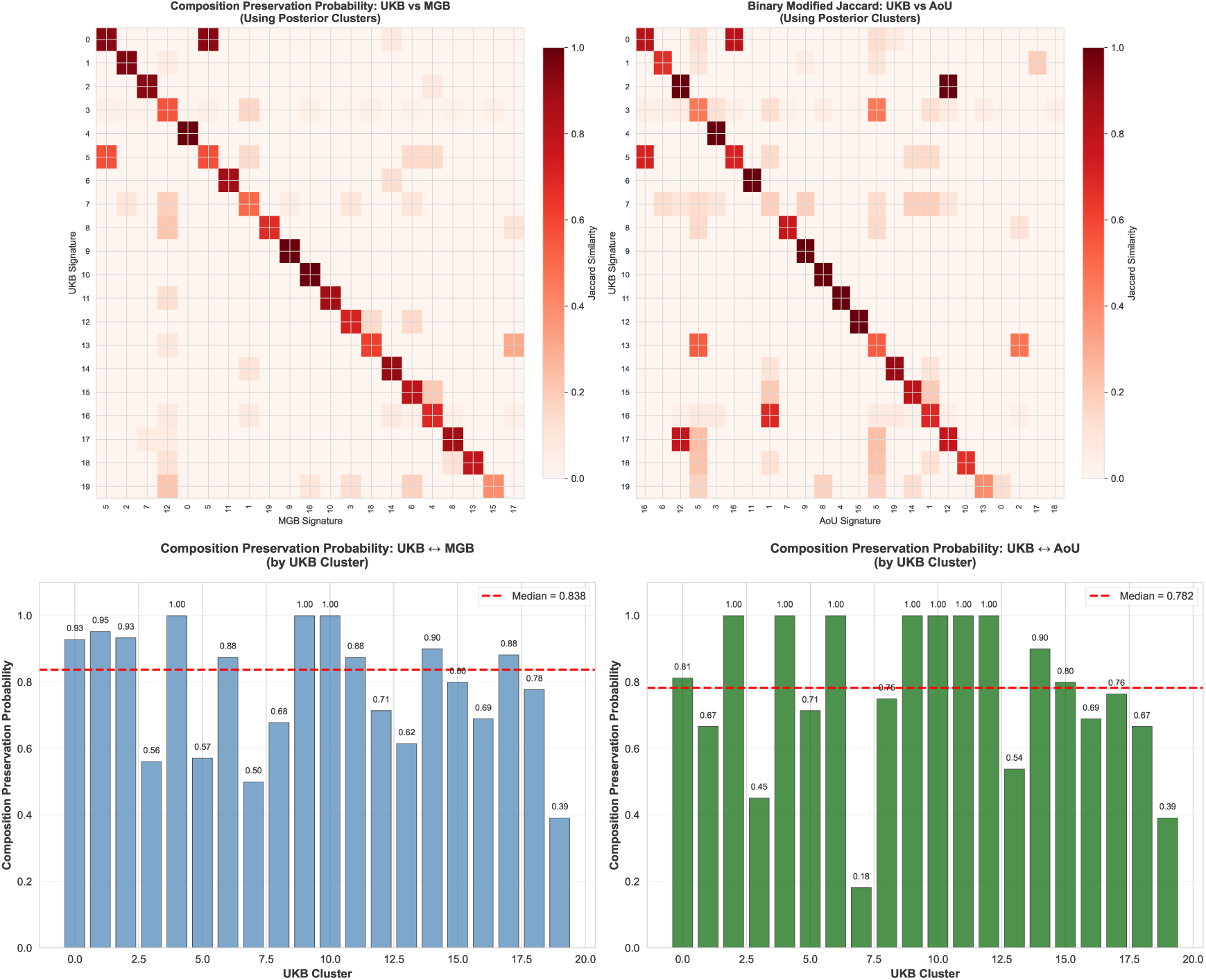
Cross-cohort validation demonstrates robust disease signature replicability. (**A**) Heatmaps showing the correspondence between `ALADYNOULLI` disease signatures (clusters) across cohorts, focusing on diseases common to all three biobanks. Each cell represents the proportion of diseases from a UK Biobank (UKB) signature that map to the corresponding signature in Mass General Brigham (MGB, left panel) or All of Us (AoU, right panel). Darker red indicates stronger correspondence. UKB clusters are ordered by their best-matching cluster in each validation cohort to highlight the diagonal pattern of correspondence. (**B**) Composition preservation probability calculation for each UKB signature. Disease-to-signature assignments are determined using posterior fitting (the signature with maximum posterior association strength, argmax*_k_ ψ_kd_* for each disease). For each UKB cluster, we identified the best-matching cluster in the comparison cohort (MGB or AoU) by finding the cluster with maximum intersection size relative to the UKB cluster size. The composition preservation probability index is calculated as the proportion of diseases in a UKB signature that also belong to its best-matching signature in the other cohort (i.e., the intersection size divided by the UKB signature size, rather than the traditional Jaccard intersection-over-union). The analysis reveals high cross-cohort replicability, with a median composition preservation probability of 0.80 (IQR: 0.667, 0.964) across both validation cohorts, indicating that 80% of diseases within each UKB signature are consistently grouped together in the corresponding signature in independent healthcare systems. This demonstrates that disease signatures identified in UK Biobank are consistently reproduced in independent populations, confirming biological validity rather than population-specific artifacts.

**Figure S6:**
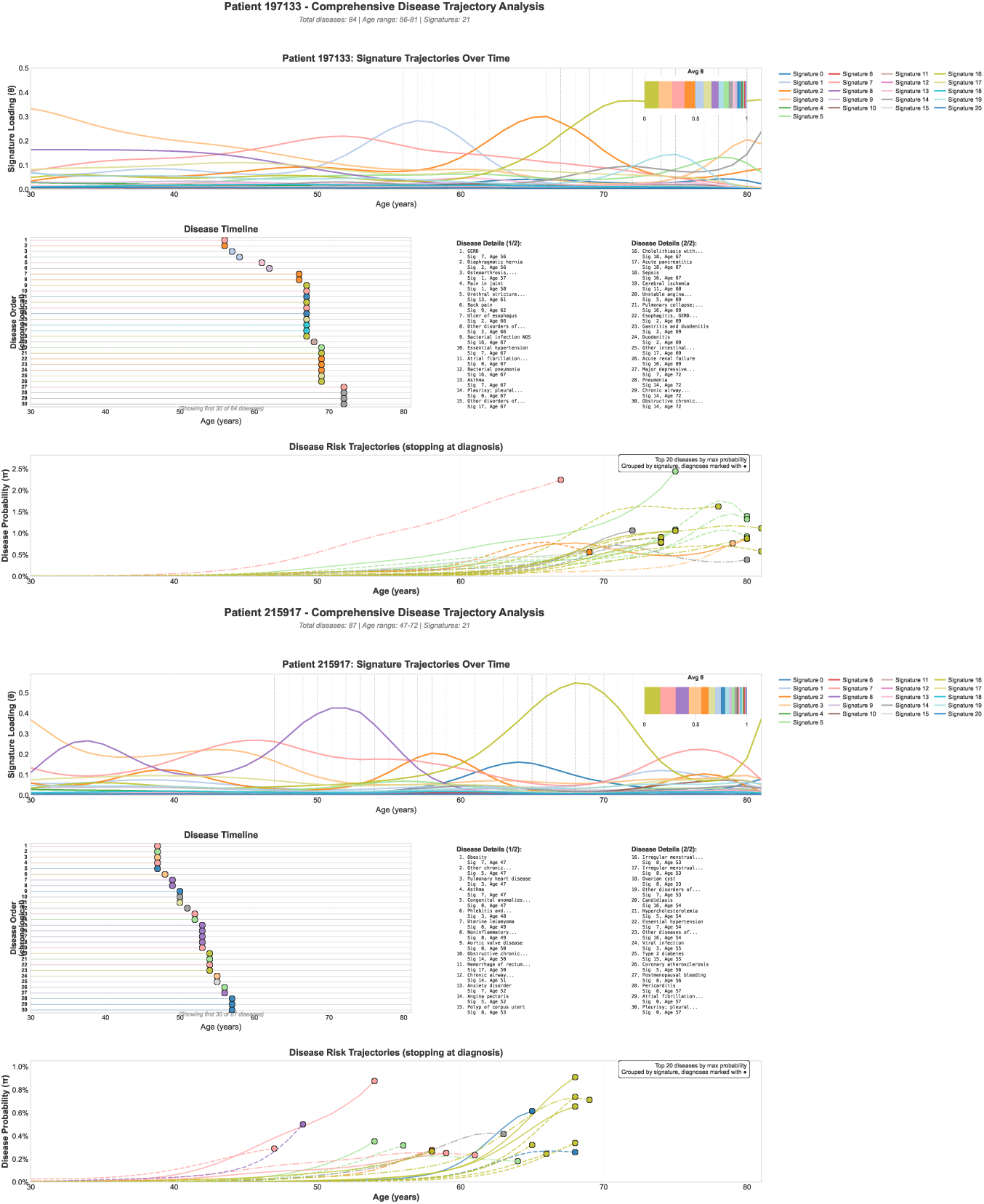
Individual patient trajectories reveal distinct patterns of disease progression. For each of two representative patients, the figure displays comprehensive disease trajectory analysis: (**Top panel**) Normalized signature loadings (*θ*) over time, showing how contributions from different disease signatures evolve throughout the patient’s life course, with vertical dotted lines indicating the timing of each disease diagnosis. (**Middle panel**) Chronological disease timeline displaying all diagnosed conditions with colors matching their primary signature assignments, providing a visual representation of disease co-occurrence patterns and temporal sequencing. (**Bottom panel**) Time-averaged signature contributions showing the overall signature profile for each patient, summarizing the relative importance of different biological pathways in their disease history. Patients are selected to demonstrate increasing complexity of disease profiles, from simpler single-signature patterns to complex multi-signature interactions involving 30+ diseases across multiple biological pathways. Colors are consistent across panels and represent the primary signature assignment for each diagnosed condition, enabling visual tracking of how signature loadings change in response to new diagnoses and how multiple signatures can contribute simultaneously to a patient’s disease risk.

**Figure S7:**
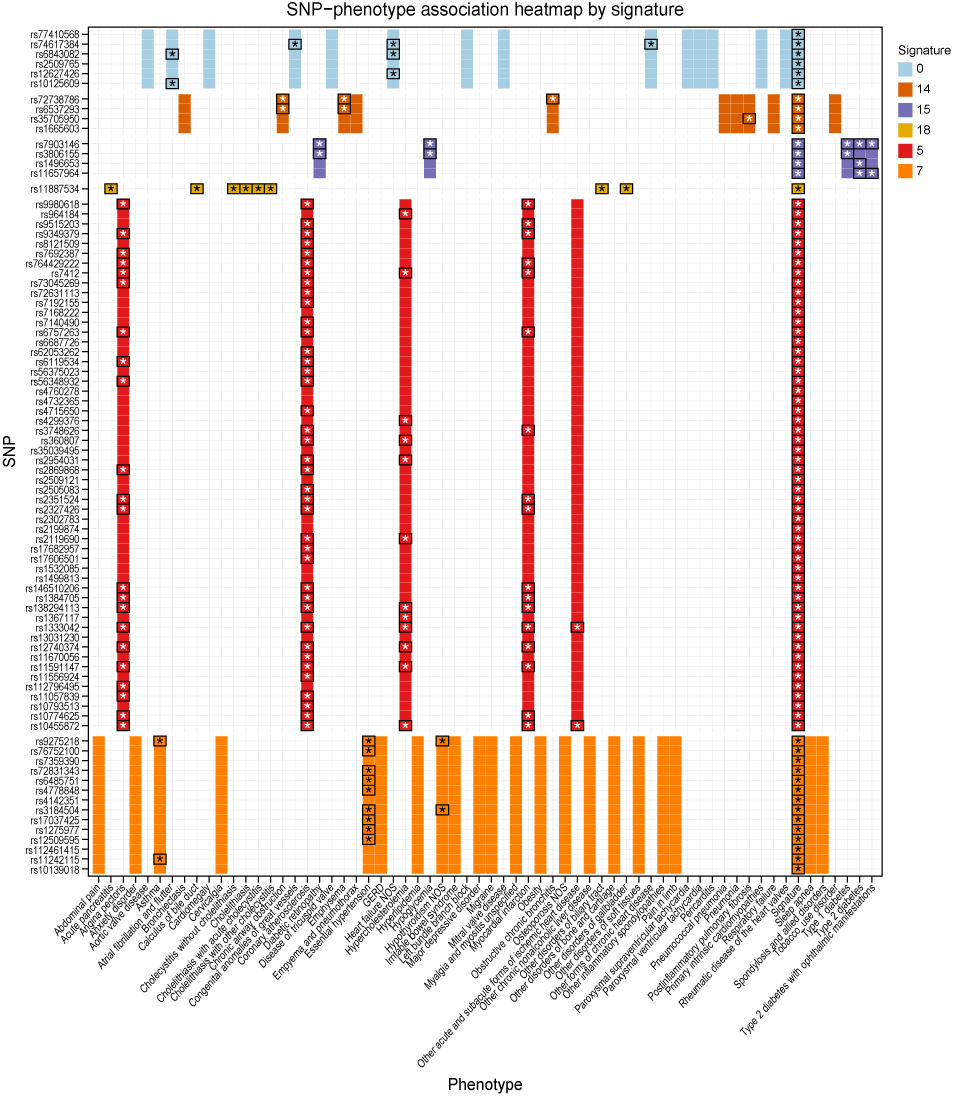
Signature-specific SNP associations reveal pleiotropic genetic effects. For each `ALADYNOULLI` disease signature, we identified lead genetic variants (SNPs) from GWAS of the area-under-the-curve (AUC) of normalized signature loadings (*θ*) across individuals. We then tested each lead SNP for association with a broad set of component disease phenotypes using logistic regression, adjusting for sex and ancestry principal components. The heatmap displays association strength (log_10_ *P*) for lead SNPs from each signature across constituent disease phenotypes, with significance (*P <* 5 10^−8^) indicated. The full matrix enables identification of signature-specific loci—variants associated with the signature but not with any single constituent disease. These results reveal pleiotropic genetic effects that are captured by the multi-disease signature but are not apparent in single-disease GWAS.

**Figure S8:**
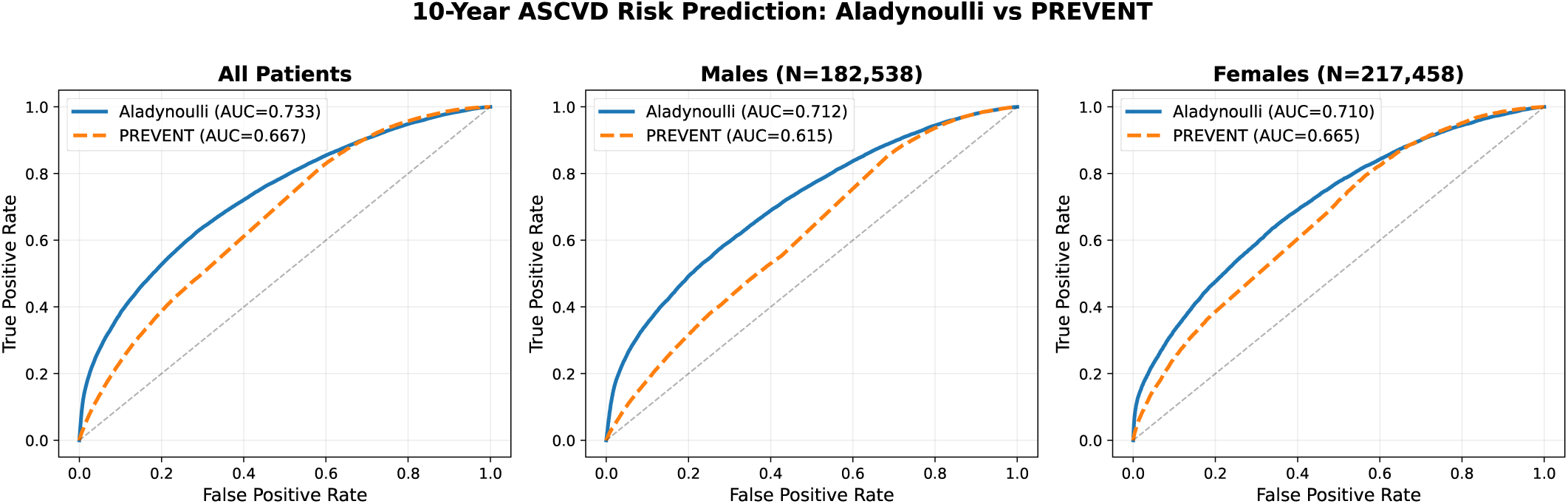
ROC curves comparing ALADYNOULLI to PREVENT cardiovascular risk score. Performance comparison for 10-year ASCVD risk prediction using predictions at enrollment to predict 10-year outcomes in (**A**) the full population (Aladynoulli AUC=0.733, PREVENT AUC=0.667), (**B**) males only (N=182,538; Aladynoulli AUC=0.712, PREVENT AUC=0.615), and (**C**) females only (N=217,458; Aladynoulli AUC=0.710, PREVENT AUC=0.665). `ALADYNOULLI` demonstrates superior discrimination across all groups compared to PREVENT. This static prediction approach (using enrollment-time predictions for a fixed 10-year horizon) is not the optimal way to use `ALADYNOULLI`, as we recommend dynamic updates that incorporate new information at each time point; however, we show this comparison for direct comparability with PREVENT, which predicts for a fixed 10-year horizon. The similar performance of `ALADYNOULLI` in males and females (AUC 0.712 vs 0.710) contrasts with the sex-specific differences observed in PREVENT, where the gap between Aladynoulli and PREVENT is larger for males (difference +0.097) than for females (difference +0.045). (*63*)

**Figure S9:**
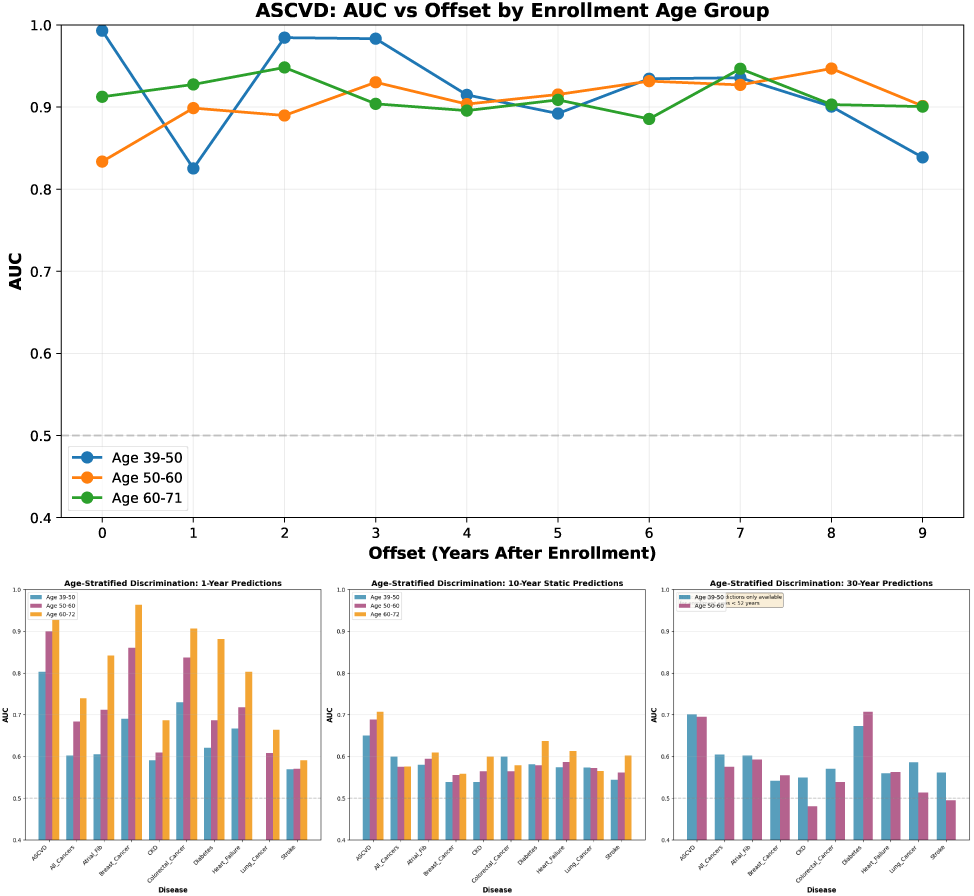
Age-stratified discrimination performance for ASCVD predictions. (**Top panel**) AUC performance across 0-9 years after enrollment, stratified by enrollment age groups (39-50, 50-60, 60-71 years). This analysis evaluates how discrimination changes when using models trained at different time offsets (enrollment + 0, 1, 2, . . ., 9 years), ensuring that each age group’s performance reflects predictions made at comparable stages of the life course. (**Bottom panel**) AUC performance for 1-year dynamic and 10-year static predictions, stratified by enrollment age groups. This analysis evaluates age-specific discrimination by grouping patients by their enrollment age (age at prediction) and assessing model performance separately for each age group. Results demonstrate robust discrimination across all age groups and time horizons, with performance improving as expected in older age groups where disease incidence is higher.

**Figure S10:**
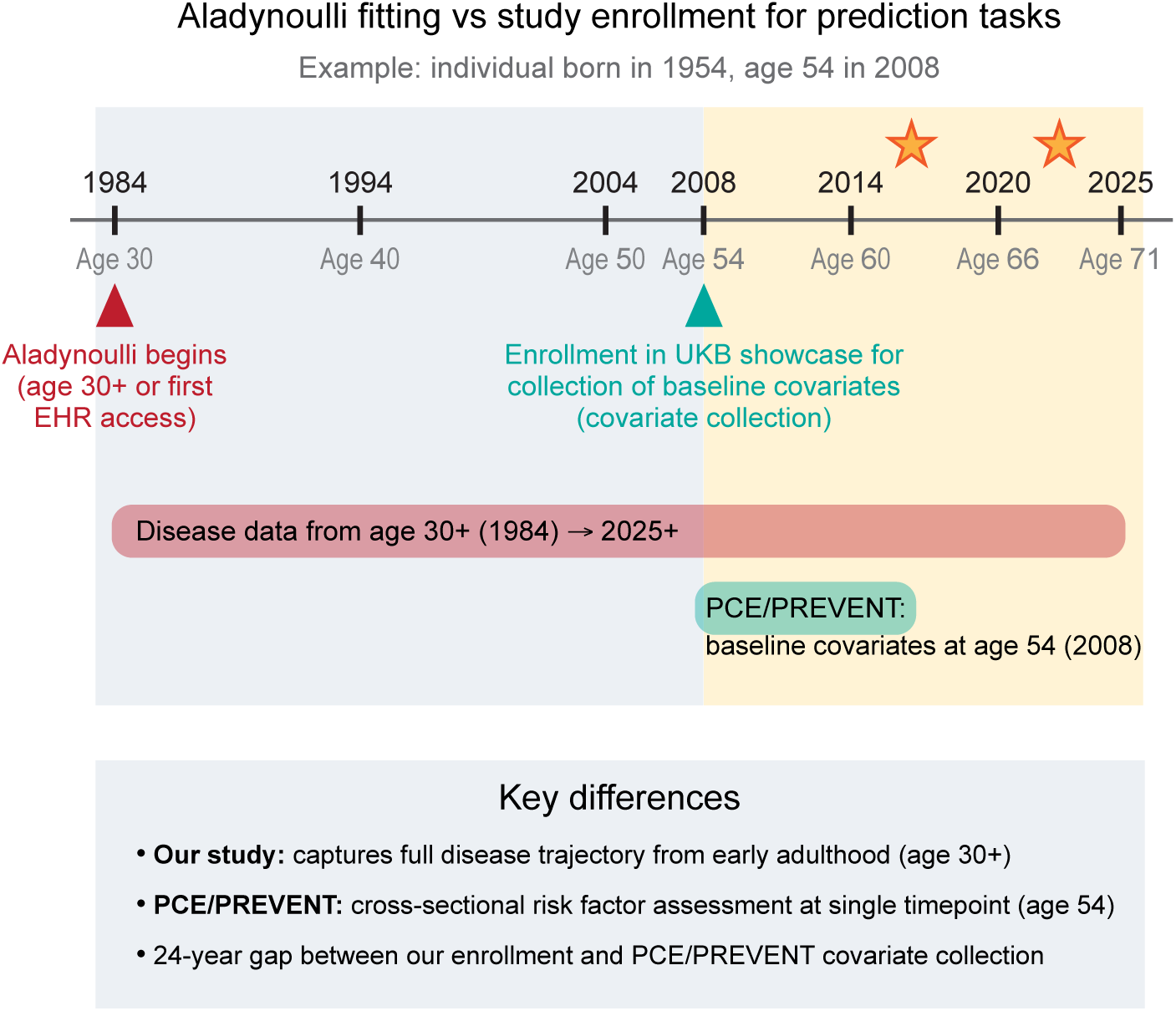
Enrollment Timeline. Schematic illustrating the distinction between the Aladynoulli model’s use of longitudinal disease history (red) and the cross-sectional covariate collection at study recruitment (green) for risk prediction tasks. In this example, an individual’s disease trajectory is captured from age 30 (or first EHR access) onward, enabling the Aladynoulli model to leverage decades of prior health data. In contrast, PCE and PREVENT models (Figure S8) use only baseline covariates collected at recruitment (age 54 in 2008). Outcome assessment is performed prospectively after recruitment (stars). The timeline highlights the 24-year gap between the start of disease data collection and the baseline covariate assessment, underscoring the unique ability of our approach to incorporate the full disease trajectory for prediction.

**Figure S11:**
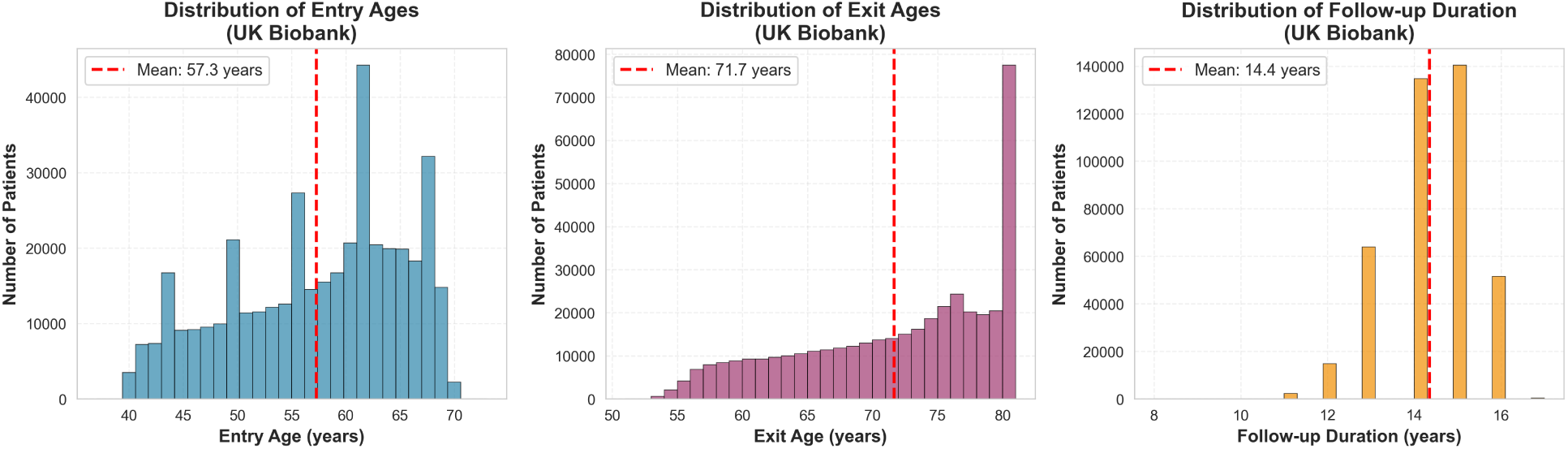
UK Biobank enrollment and follow-up characteristics. Summary statistics and distributions for entry age (enrollment age), exit age (last follow-up), and follow-up duration. Entry age shows a median of 59 years (range: 37-73 years), with individuals enrolling between 2006-2010. Exit age shows a median of 73 years (range: 51-81 years), reflecting follow-up through 2023/2024. Follow-up duration shows a median of 14.4 years (range: 8-17 years), demonstrating substantial longitudinal follow-up. The distributions illustrate the age range and follow-up periods available for prediction analysis, where we evaluate the first 10 years from enrollment to ensure consistent evaluation across all participants.

**Figure S12:**
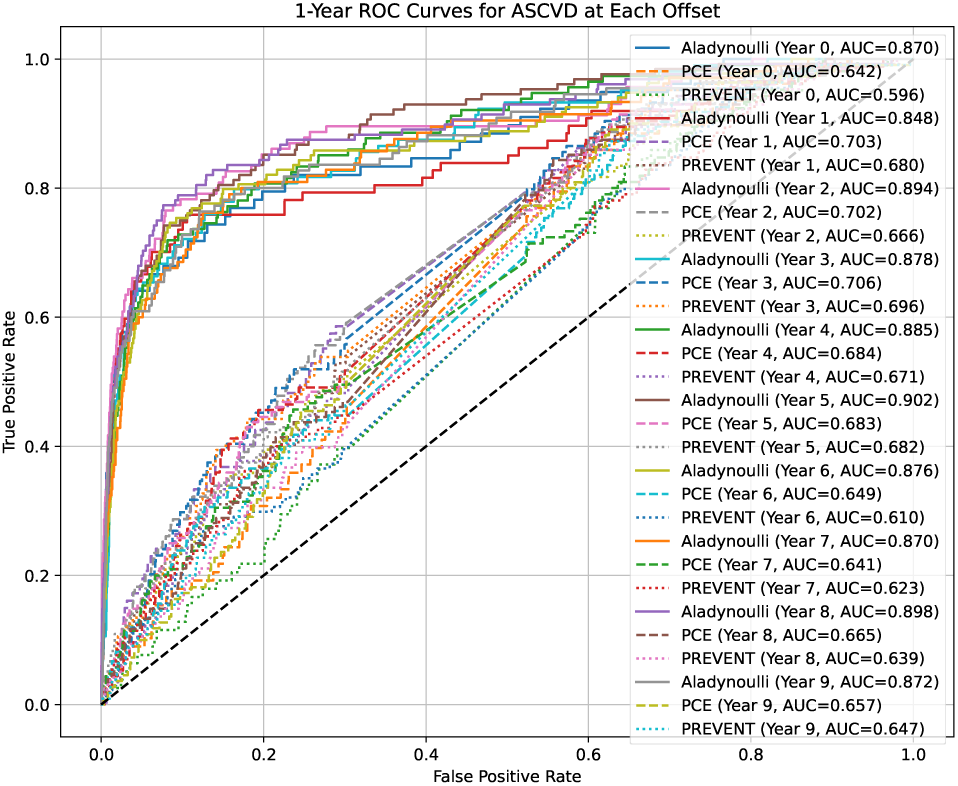
Distribution of 1-year AUCs across offsets for ASCVD versus commonly used clinical risk scores. Receiver operating characteristic (ROC) curves for 1-year ASCVD risk prediction at each followup year, comparing the Aladynoulli model (solid lines), Pooled Cohort Equations (PCE, dashed lines), and PREVENT (dotted lines). For each offset (year since recruitment), the model’s predicted 1-year risk is evaluated against observed 1-year outcomes, excluding individuals with prevalent ASCVD at the start of each interval. The area under the curve (AUC) for each method and year is shown in the legend. This visualization demonstrates the discrimination performance of each approach over time, highlighting the dynamic updating capability of Aladynoulli compared to static clinical risk scores.

**Figure S13:**
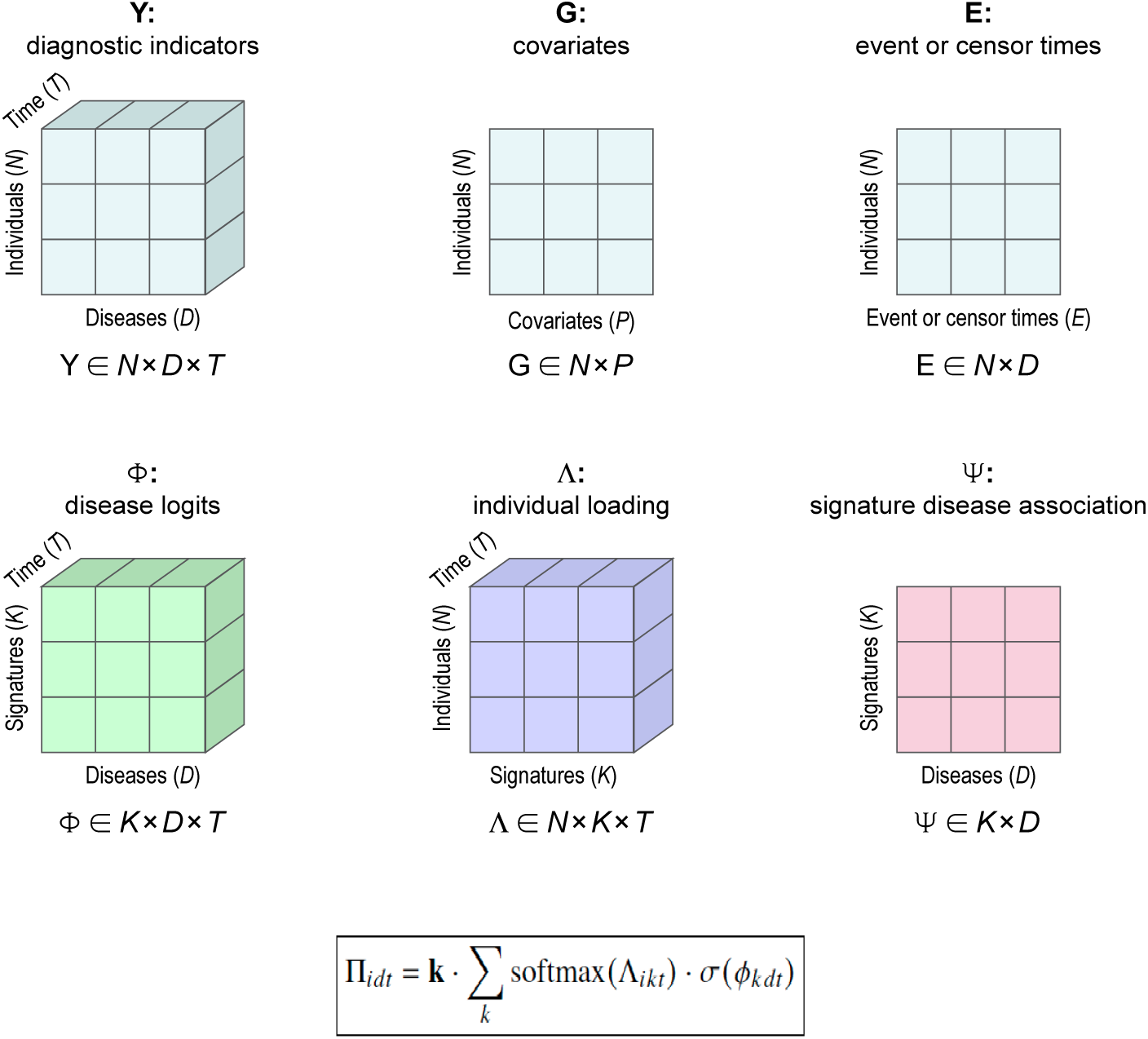
ALADYNOULLI data structure and model components. The figure illustrates the key data matrices and their relationships in the `ALADYNOULLI` framework. Top row: Input data includes Y (diagnostic indicators, a 3D tensor of binary disease outcomes across individuals, diseases, and time), G (covariates matrix for individuals), and E (event or censoring times). Bottom row: Model parameters include Φ (disease logits by signature and time), Λ (individual loadings representing time-varying signature associations), and Ψ (static signature-disease association strengths). The mathematical formula shows how these components combine to generate disease probabilities Π*_idt_* through a mixture of softmax-transformed individual loadings and sigmoid-transformed disease logits, scaled by a global calibration parameter.

**Figure S14:**
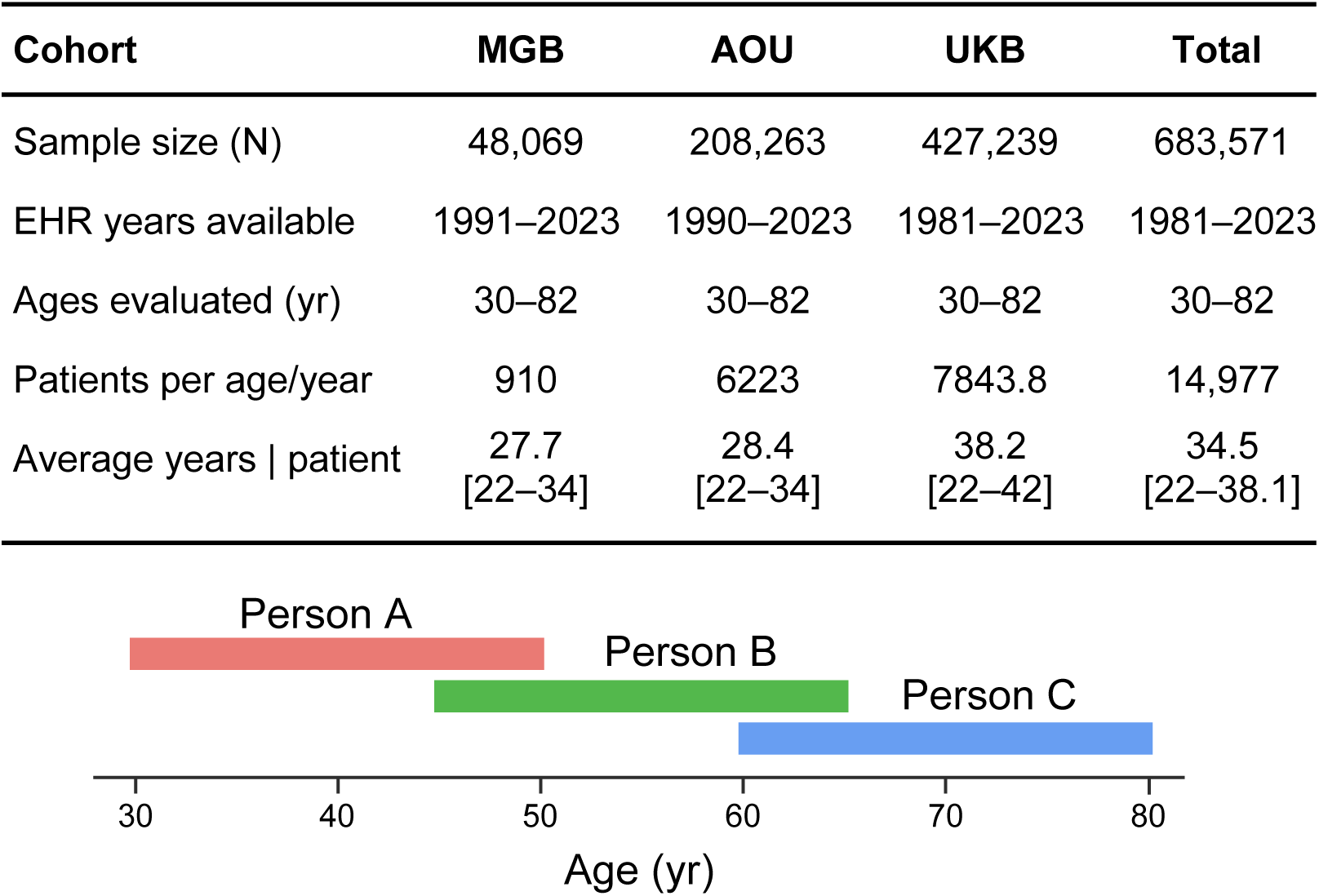
Cohort characteristics and study design. Integration of partial life trajectories from three age groups (A: 30-50, B: 45-65, C: 60-82 years). The overlapping periods within each cohort enable robust estimation of disease trajectories across the full age spectrum.

**Figure S15:**
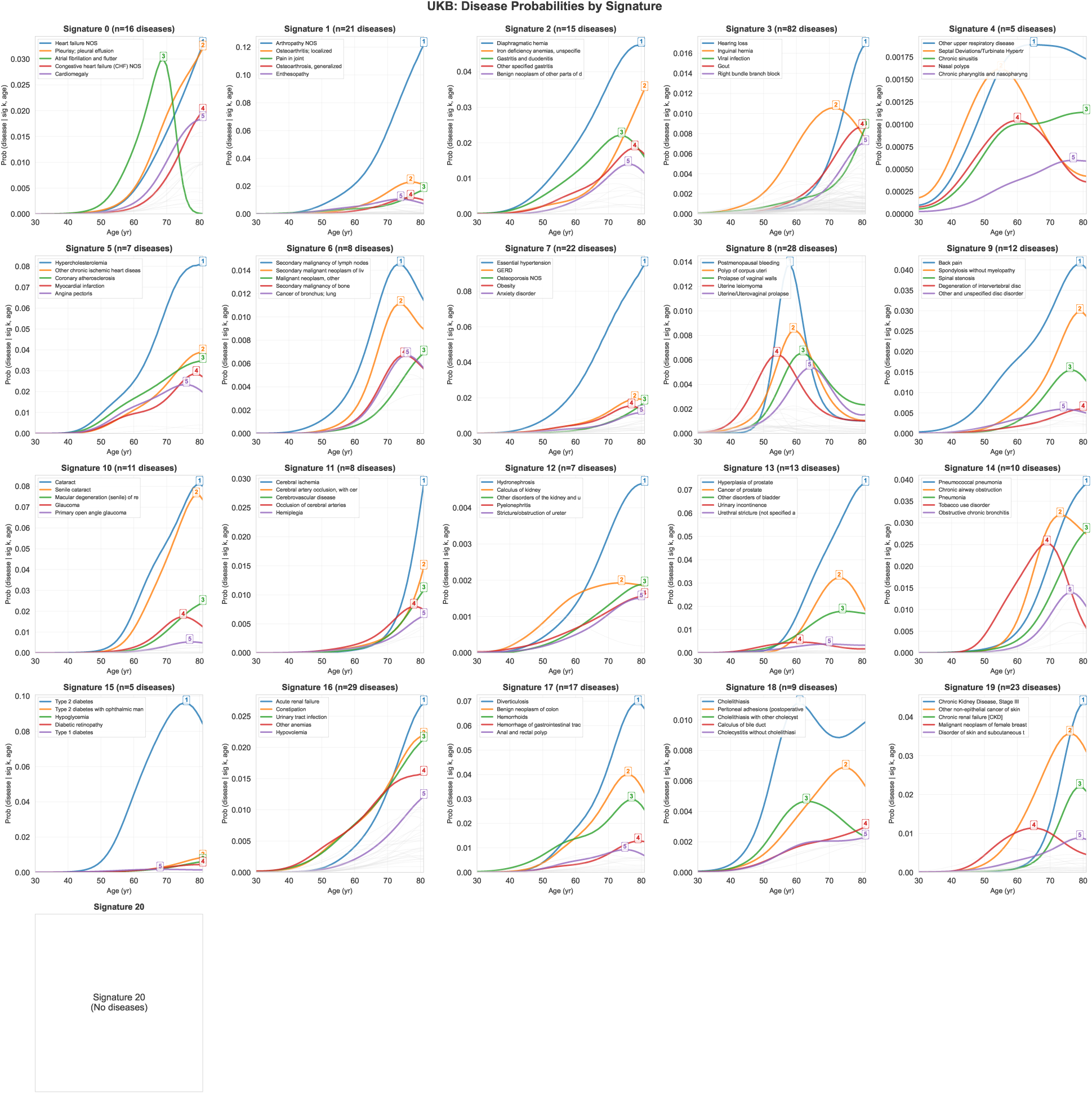
Temporal patterns of disease signatures across age in UK Biobank. Each panel displays age-dependent disease probabilities for one of 21 disease signatures (Signatures 0-20), showing how disease risk evolves from ages 30-81 years. For each signature, colored lines represent diseases assigned to that signature (based on maximum posterior *ψ_kd_* association strength), while light grey lines show background diseases assigned to other signatures. The y-axis displays the probability of each disease given membership in the signature and age, computed by applying the sigmoid function to the time-varying disease-signature association parameters (*ϕ_kdt_*) from the model. These probability trajectories reveal how diseases within each signature exhibit characteristic age-dependent risk patterns, with some diseases showing early-onset patterns (peaking in middle age) and others showing later-onset patterns (increasing with age), demonstrating the model’s ability to capture clinically meaningful temporal disease progression within biological pathways.

**Figure S16:**
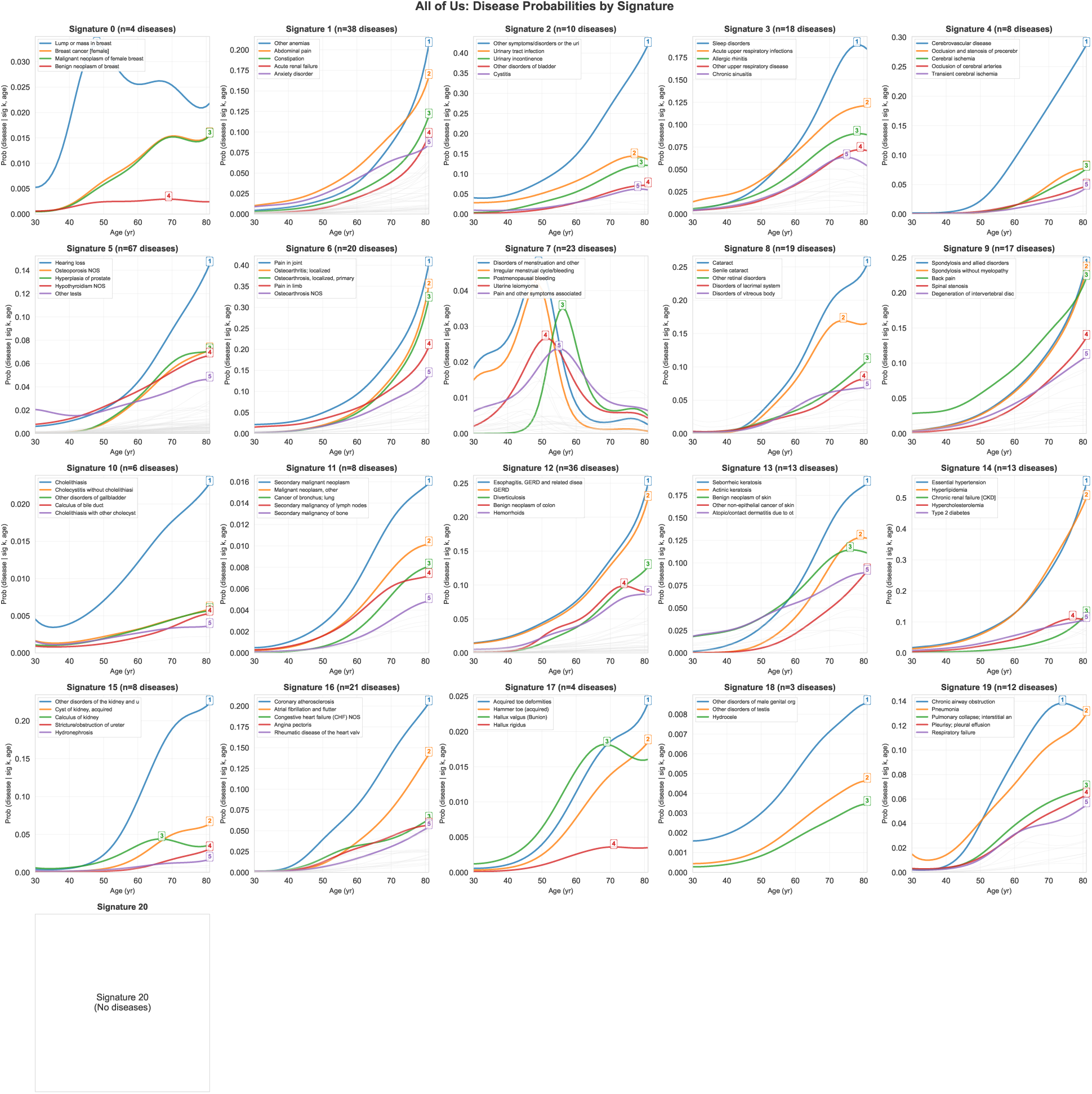
Temporal patterns of disease signatures across age in All of Us. Each panel displays age-dependent disease probabilities for one of 21 disease signatures (Signatures 0-20), showing how disease risk evolves from ages 30-81 years. For each signature, colored lines represent diseases assigned to that signature (based on maximum posterior *ψ_kd_* association strength), while light grey lines show background diseases assigned to other signatures. The y-axis displays the probability of each disease given membership in the signature and age, computed by applying the sigmoid function to the time-varying disease-signature association parameters (*ϕ_kdt_*) from the model. These probability trajectories reveal how diseases within each signature exhibit characteristic age-dependent risk patterns, with some diseases showing early-onset patterns (peaking in middle age) and others showing later-onset patterns (increasing with age), demonstrating the model’s ability to capture clinically meaningful temporal disease progression within biological pathways.

**Figure S17:**
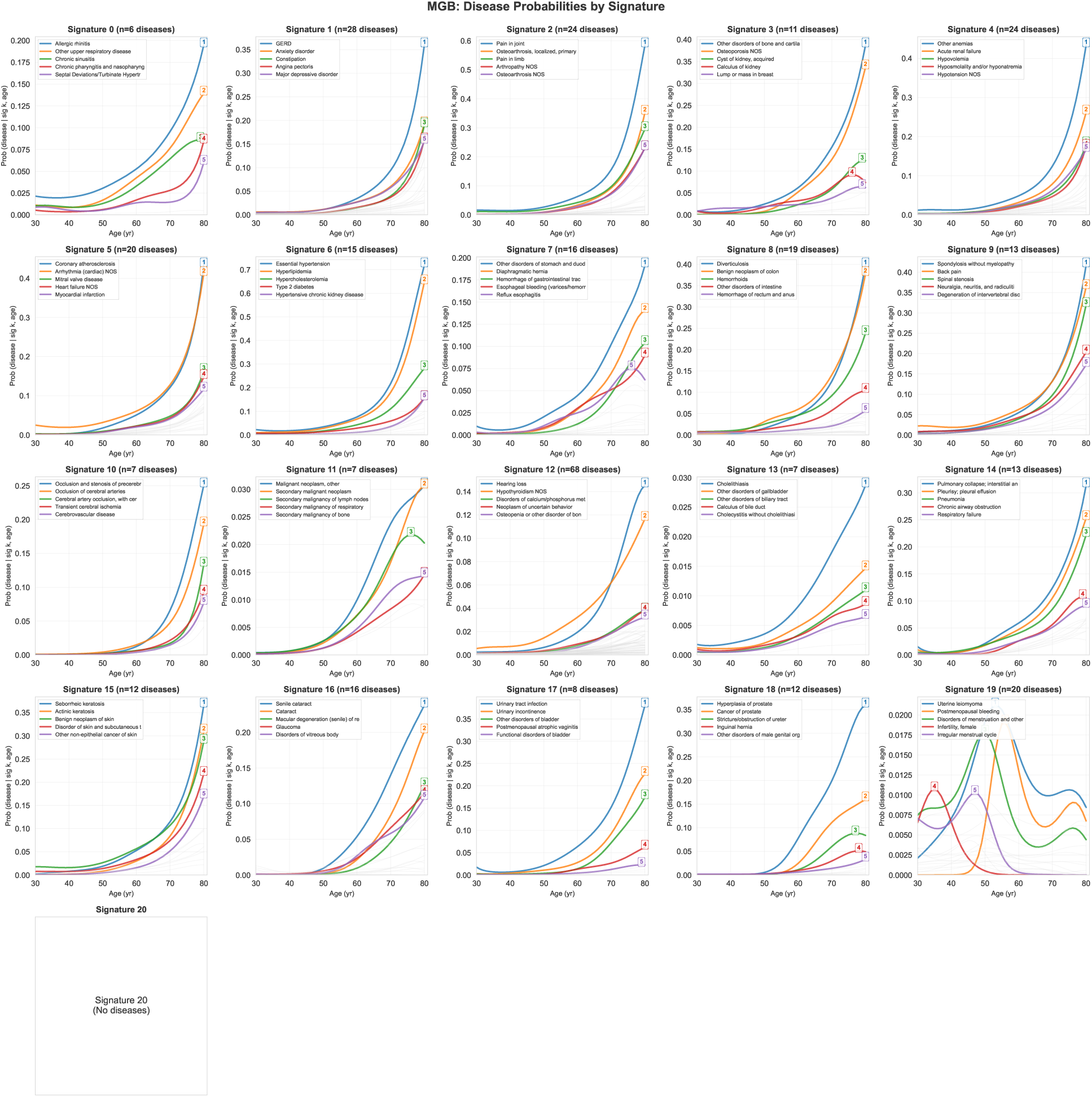
Temporal patterns of disease signatures across age in MGB Biobank. Each panel displays age-dependent disease probabilities for one of 21 disease signatures (Signatures 0-20), showing how disease risk evolves from ages 30-81 years. For each signature, colored lines represent diseases assigned to that signature (based on maximum posterior *ψ_kd_* association strength), while light grey lines show background diseases assigned to other signatures. The y-axis displays the probability of each disease given membership in the signature and age, computed by applying the sigmoid function to the time-varying disease-signature association parameters (*ϕ_kdt_*) from the model. These probability trajectories reveal how diseases within each signature exhibit characteristic age-dependent risk patterns, with some diseases showing early-onset patterns (peaking in middle age) and others showing later-onset patterns (increasing with age), demonstrating the model’s ability to capture clinically meaningful temporal disease progression within biological pathways.

**Figure S18:**
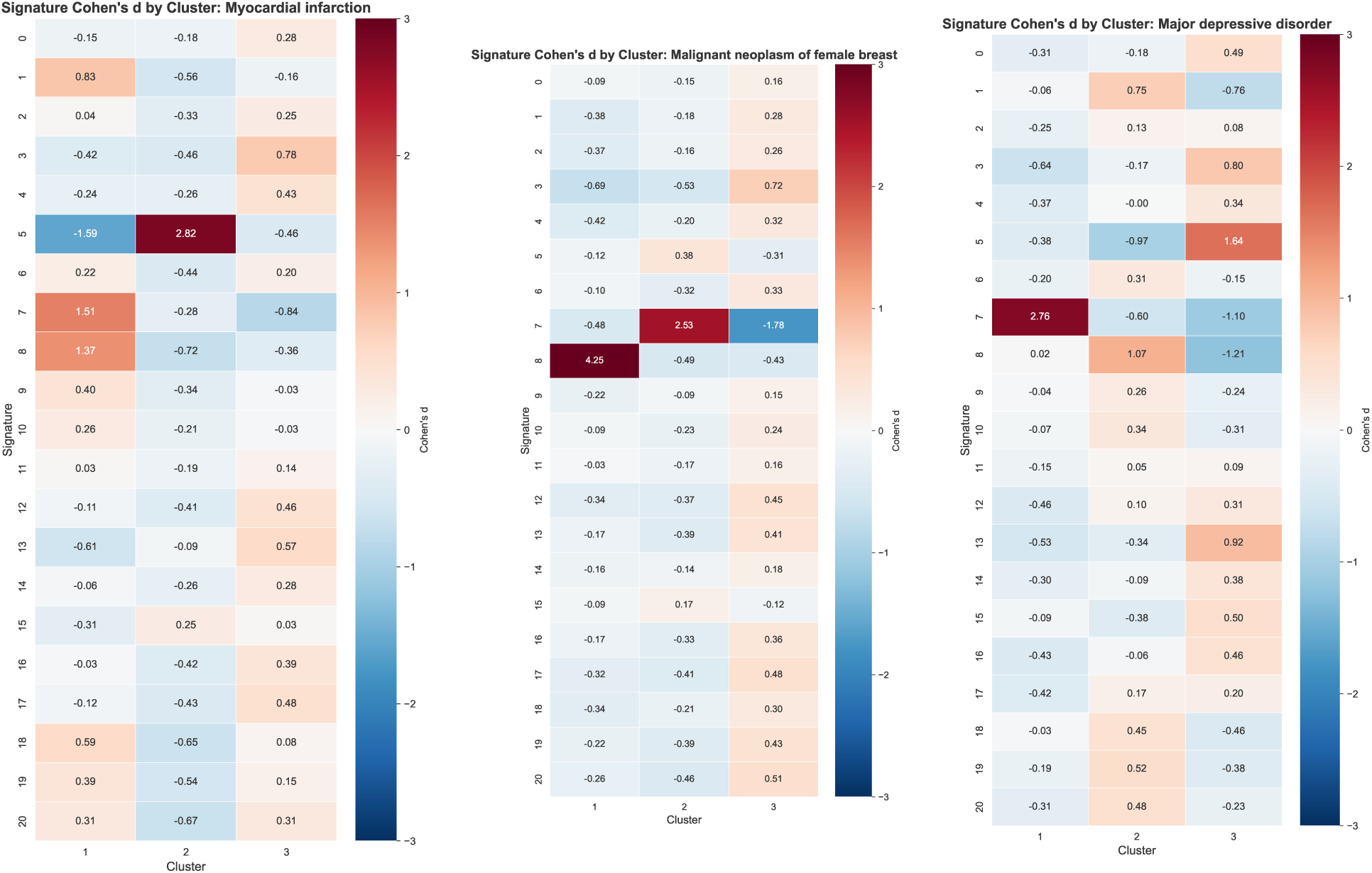
Signature-based patient stratification reveals biological heterogeneity within clinical diagnoses. Cohen’s d effect sizes quantifying the separation of time-averaged signature loadings between patient clusters for three representative diseases: (**Left**) Myocardial infarction, (**Center**) Breast cancer, and (**Right**) Major depressive disorder. For each disease, patients were first identified and then clustered into three subgroups using k-means clustering applied to their time-averaged normalized signature loadings (*θ̄_ik_* = <u>^1^</u> *_t_ θ_ikt_*), where *θ_ikt_* represents patient *i*’s normalized association with signature *k* at time *t* (see **Figure 3C** for visualization of these illustrative clusters). For each cluster *c* and signature *k*, Cohen’s *d* was calculated as the standardized difference: *d_ck_* = *θ̄_in_ θ̄_out_ s_Pooled_*, where *θ̄_in_* and *θ̄_out_* are the mean timeaveraged signature loadings for patients within and outside cluster *c*, respectively, and *s_Pooled_* is the pooled standard deviation. Each heatmap displays signatures (rows) and clusters (columns), with color intensity representing Cohen’s *d* values. Large positive values *d* 0.8 indicate strong enrichment of a signature within that patient subgroup (red), while negative values indicate depletion (blue). Values are centered at zero, with the color scale ranging from -3 to +3. These clusters are illustrative examples of heterogeneity rather than definitive diagnostic categories; they demonstrate that patients sharing the same clinical diagnosis can exhibit distinct underlying biological patterns reflected in their signature profiles. This analysis reveals substantial biological heterogeneity within traditional diagnostic categories, with different disease signatures showing varying degrees of patient stratification across clusters.

**Figure S19:**
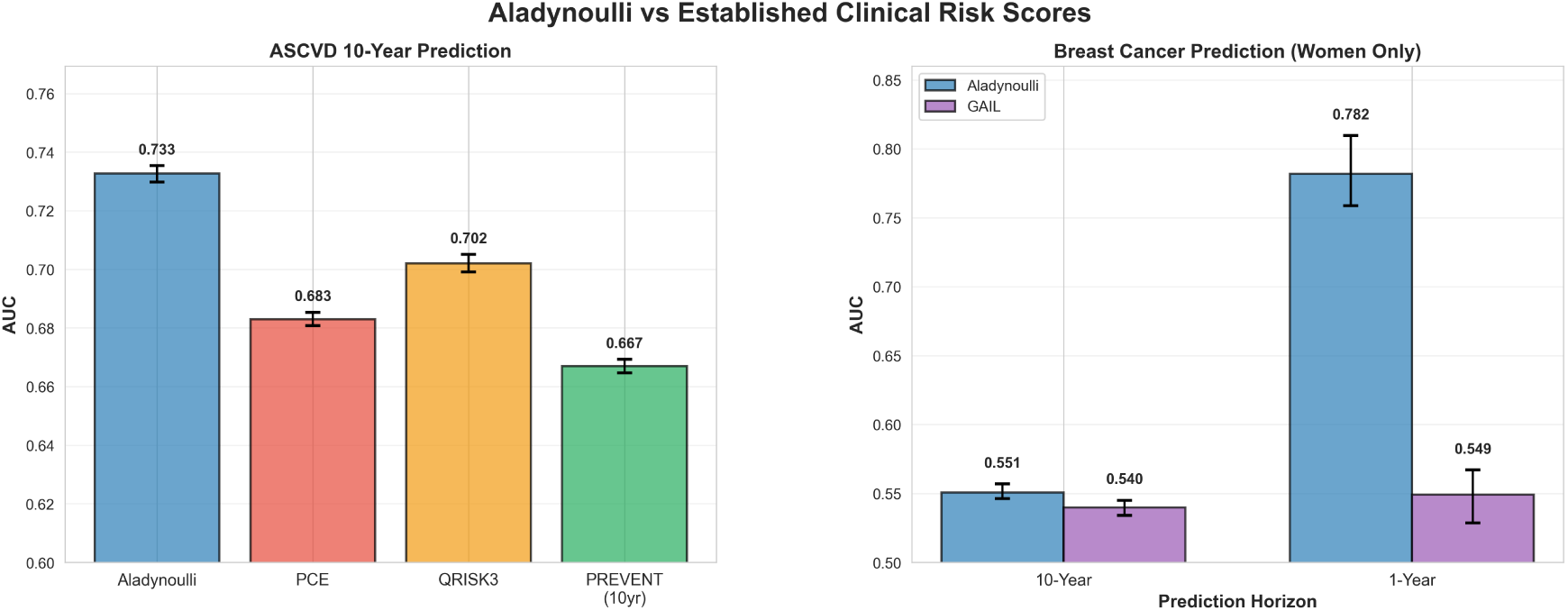
Comparison of ALADYNOULLI with external clinical risk scores. (**A**) 10-year ASCVD risk prediction comparing `ALADYNOULLI` (AUC: 0.733) with PCE (AUC: 0.683), QRISK3 (AUC: 0.702), and PREVENT (AUC: 0.667). (**B**) 10-year breast cancer risk prediction for women only, comparing `ALADYNOULLI` (AUC: 0.551) with GAIL (AUC: 0.540), demonstrating a difference of +0.011. (**C**) 1-year breast cancer risk prediction for women only, comparing `ALADYNOULLI` (AUC: 0.782) with GAIL (AUC: 0.549), demonstrating a difference of +0.233. Error bars indicate 95% confidence intervals.

**Figure S20:**
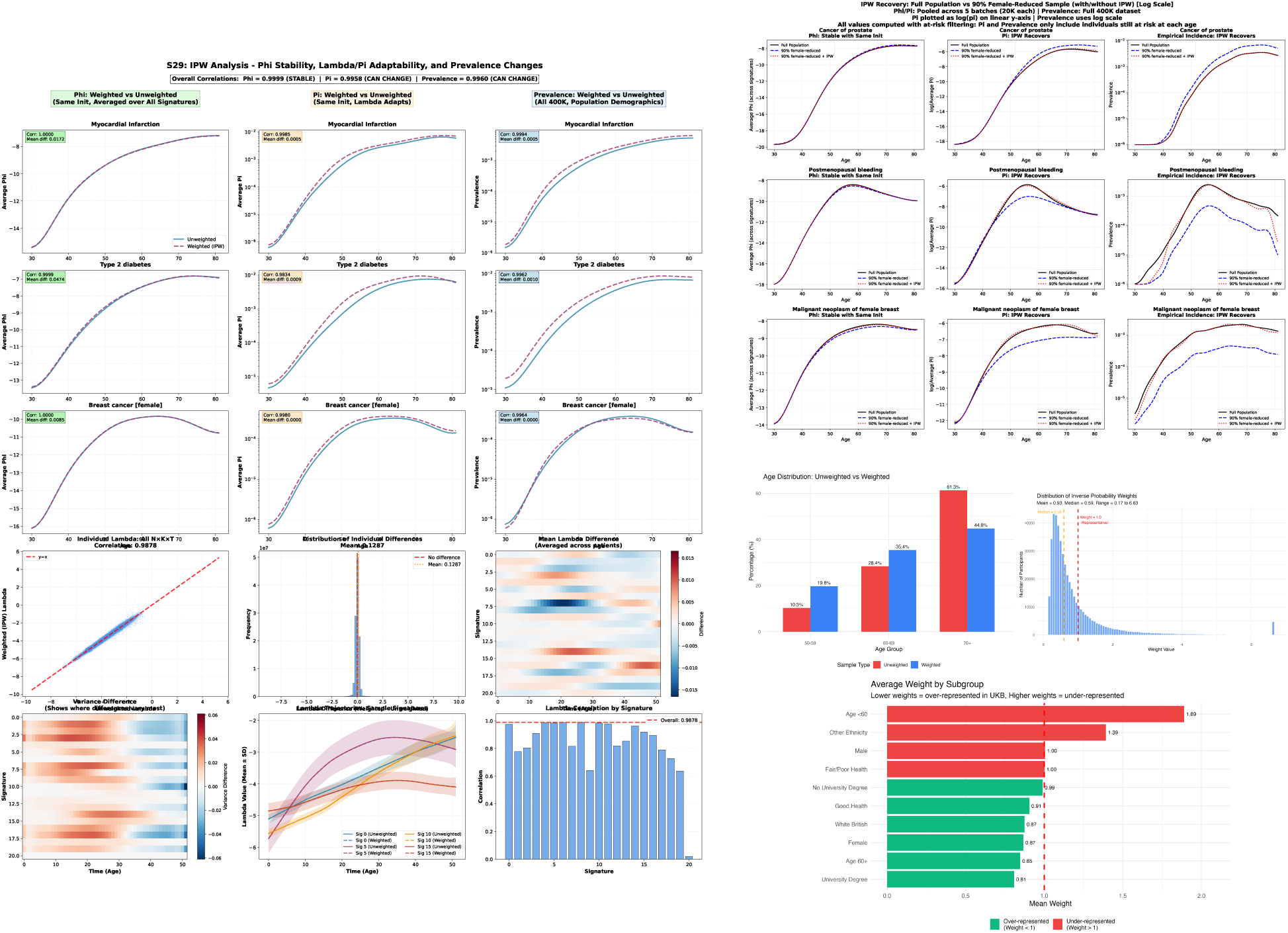
Inverse probability weighting (IPW) analysis for selection bias correction. IPW corrects for UK Biobank selection bias in two complementary directions: (**1**) Forward correction: When applying IPW weights derived from (*39*) to the full UKB sample (which is unbalanced relative to the general population), *ϕ*(signature-disease associations) remains stable (correlation ≥ 0.99), preserving biological disease-signature relationships, while lambda and pi adapt to capture the reweighted population characteristics. (**2**) Reverse correction: When taking a restricted subsample (e.g., dropping a randomly sampled 90% of women) and applying IPW reweighting, the model successfully recovers the full population prevalence patterns and predicted hazards, demonstrating that IPW can restore unbiased population estimates from biased samples. (**Left panel**) IPW analysis overview showing weights distribution, *ϕ*/*π*/prevalence comparisons, and individual *λ* differences (see Extended Data for full details). (**Right panel, top**) Model parameter comparison for full population vs contrived unbalanced subsample (90% women dropped) with and without IPW, showing three columns: (**Left**) *ϕ* trajectories (stable across conditions, correlation ≥0.99), (**Middle**) *π*trajectories, and (**Right**) *empirical prevalence* trajectories (observed disease rates calculated from the data, not the model parameter *μ_d_*; weighted vs unweighted showing bias and recovery) across three diseases (Prostate cancer, Postmenopausal bleeding, Breast cancer, from top to bottom). Note that *μ_d_* (disease baseline prevalence) was held fixed across all three scenarios to isolate selection bias effects on other parameters. (**Right panel, bottom**) Demographic distributions showing how IPW reweights the sample: (**Left**) age distribution before and after weighting, (**Middle**) IPW weights distribution (mean=0.93, median=0.59, range 0.17-6.63), and (**Right**) average weights by demographic subgroup, demonstrating that IPW corrects for underrepresentation of older, less healthy, and non-White British participants in UK Biobank. IPW preserves biological disease-signature associations (*ϕ*) while enabling accurate estimation of unbiased population characteristics (*λ*, *π*) in both forward and reverse directions.

**Figure S21:**
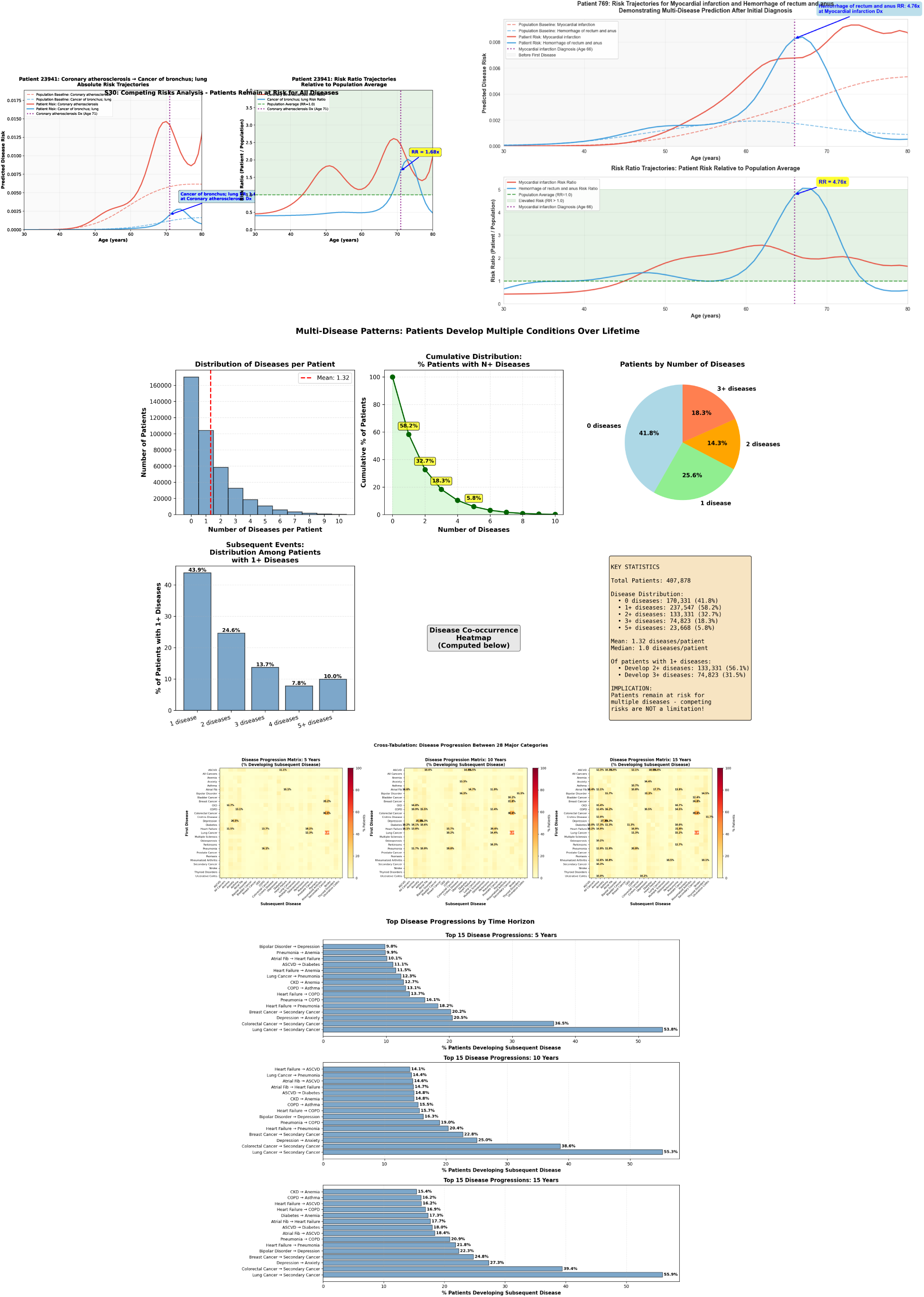
Competing risks and subsequent disease analysis. (**A-B**) Patient trajectory examples showing multiple competing outcomes over time. (**C**) Temporal patterns of subsequent disease development, illustrating how diseases develop sequentially and interact over the life course. (**D-E**) Cross-tabulation analyses showing disease co-occurrence patterns. (**F**) Multi-disease pattern visualization demonstrating how diseases cluster and progress together.

**Figure S22:**
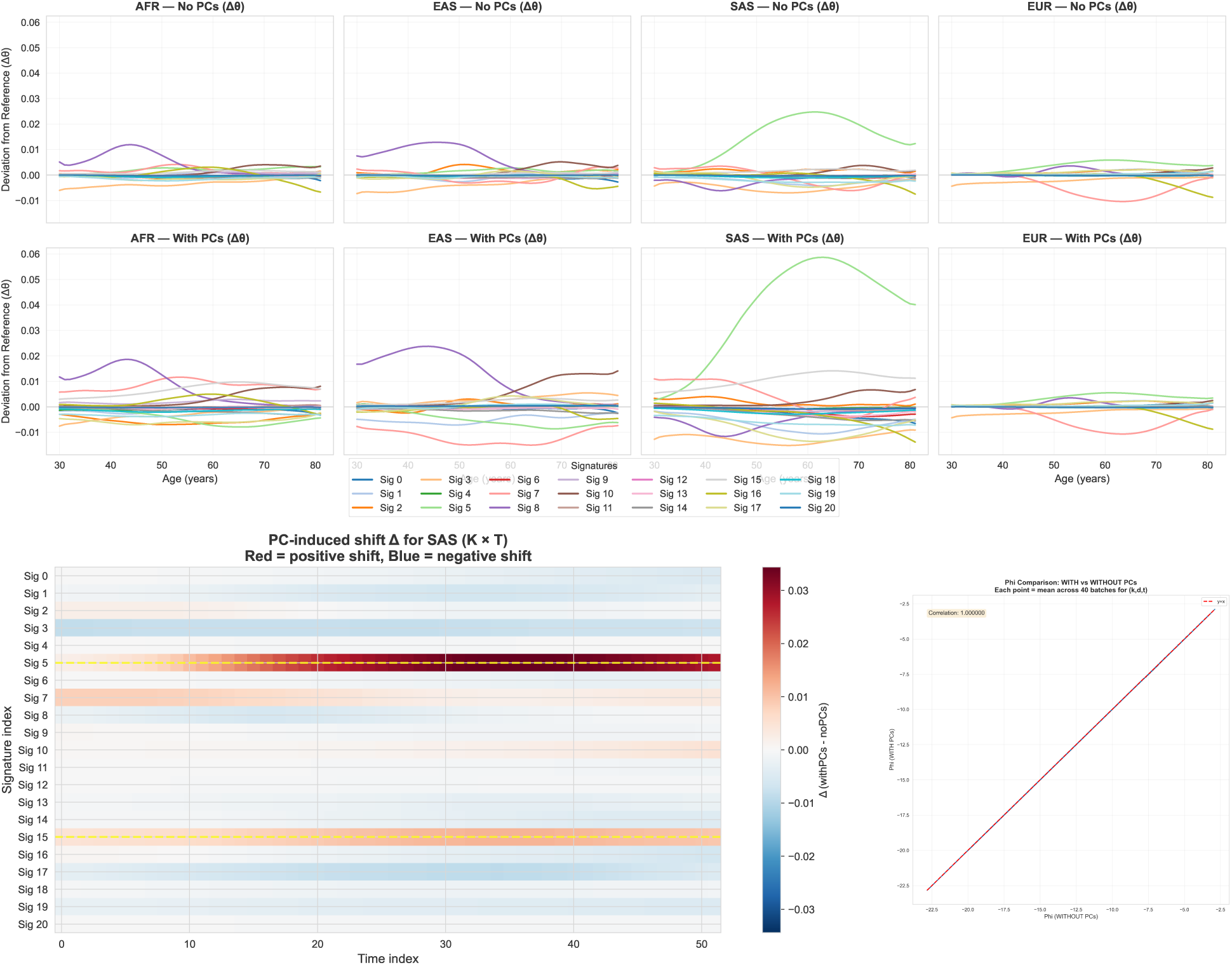
Population stratification and principal component adjustment analysis. (**A**) Signature loading deviations from population mean (Δ*θ*) over age (30-80 years) for four major ancestry groups: African (AFR), East Asian (EAS), South Asian (SAS), and European (EUR). Top row shows analyses without Principal Components (PCs); bottom row shows analyses with PCs included. The temporal dimension reveals that ancestry effects on signatures evolve with age, with peak deviations occurring at different life stages for different ancestries. When PCs are included, ancestry-specific patterns are amplified, demonstrating that genetic ancestry substantially influences signature loadings and that explicitly modeling ancestry reveals stronger ancestry-specific biological patterns. (**B**) Correlation of signature-disease associations (*ϕ*) with and without PC adjustment. Each point represents the mean across 40 batches for each signature-disease-time combination (k, d, t). The high correlation demonstrates minimal structural changes to relationships between signatures and diseases when ancestry is explicitly modeled, indicating that PC adjustment preserves disease-signature associations while accounting for population structure. (**C**) PC-induced shift (Δ) for South Asian (SAS) ancestry displayed as a heatmap across signatures (K) and time (T). Red indicates positive shifts (amplification of signature loadings when PCs are included), while blue indicates negative shifts (reduction). This visualization quantifies how PC adjustment modifies signature loadings for specific ancestry groups, with South Asian ancestry showing particularly strong shifts in cardiovascular Signature 5 and other key signatures. Together, these panels demonstrate that PC adjustment: (1) preserves core biological disease-signature associations (*ϕ*stability), (2) reveals amplified ancestry-specific signature loading patterns that evolve across the life course, and (3) provides stronger evidence for biological validity by demonstrating direct, interpretable relationships between ancestry and biologically meaningful disease signatures.

**Figure S23:**
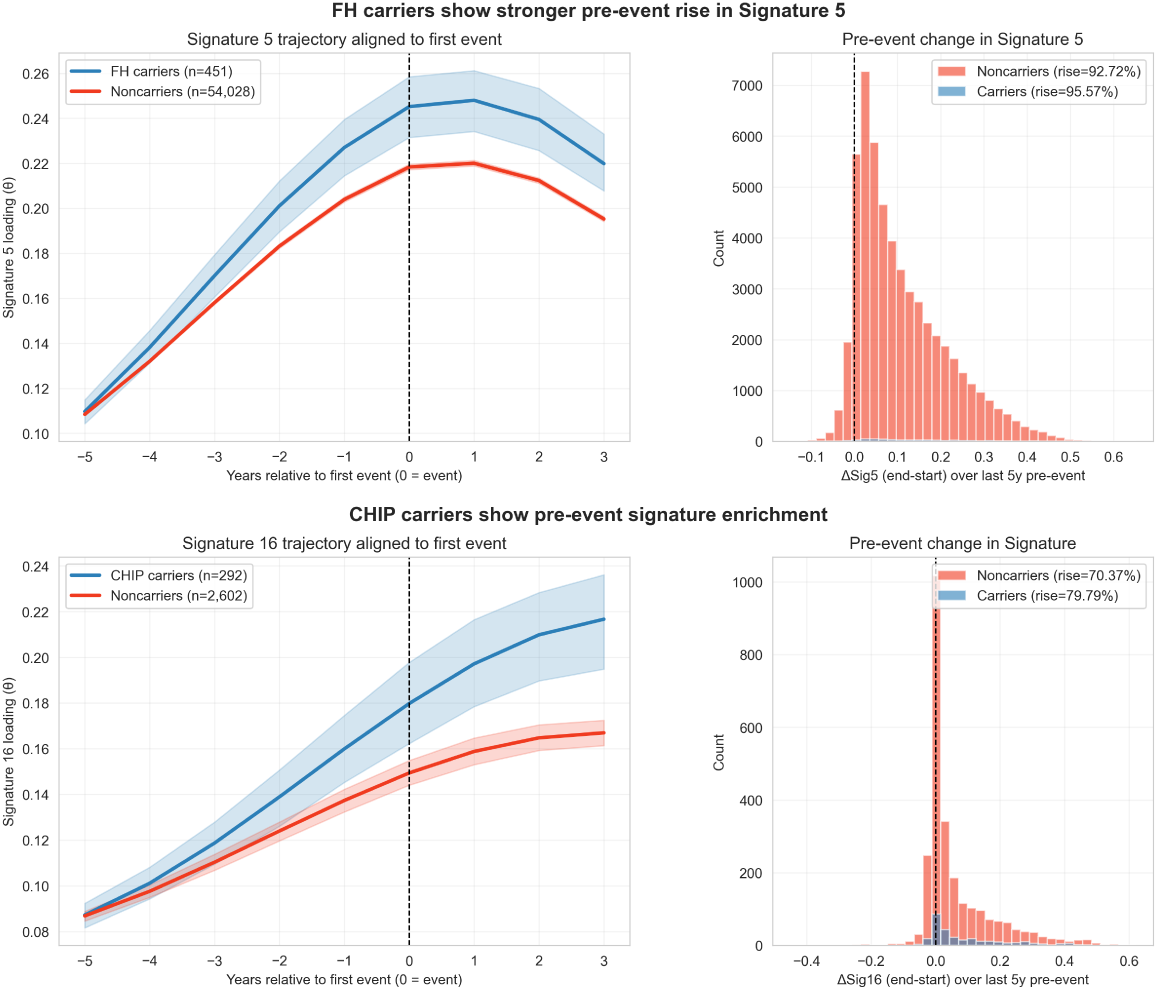
Genetic validation of signature biological meaningfulness. We demonstrate clinical and biological meaningfulness through multiple lines of evidence: (**Top panel**) **Familial Hypercholesterolemia (FH) validation**: (**Left**) Signature 5 (cardiovascular signature) loading trajectories aligned to first ASCVD event for FH carriers (n=451, blue) versus non-carriers (n=54,028, red). Trajectories are aligned such that year 0 represents the time of the first cardiovascular event, showing signature loadings from 5 years before to 3 years after the event. FH carriers show consistently higher Signature 5 loadings across the entire period, with both groups showing a clear rise leading up to the event, peaking around the time of the event. (**Right**) Distribution of pre-event change in Signature 5 loading (ΔSig5) over the last 5 years before the event, calculated as the difference between signature loading at the end of the 5-year window (1 year before event) and at the start (5 years before event). FH carriers show a higher proportion experiencing a rise (96.1%, n=446/464) compared to noncarriers (93.5%, n=52,221/55,844), with an odds ratio of 1.7 (Fisher’s exact test, p=0.011). This demonstrates enrichment of Signature 5 activation before ASCVD events in individuals with known genetic predisposition to hypercholesterolemia, validating the LDL/cholesterol → cardiovascular disease pathway. (**Bottom panel**) **Clonal Hematopoiesis of Indeterminate Potential (CHIP) validation**: (**Left**) Signature 16 (critical care/inflammation signature) loading trajectories aligned to first event for CHIP carriers (n=1,311, blue) versus noncarriers (n=14,717, red). CHIP carriers show consistently higher Signature 16 loadings and a more pronounced increase, particularly in the years leading up to and following the event. (**Right**) Distribution of pre-event change in Signature 16 loading (ΔSig16) over the last 5 years before the event. CHIP carriers show a higher proportion experiencing a rise (66.4%) compared to noncarriers (63.2%), demonstrating enrichment of inflammatory signatures before events. Analysis across multiple CHIP mutations and outcomes reveals consistent patterns: DNMT3A carriers show 1.97-fold enrichment (OR=1.97, p=0.0007) for Signature 16 before Leukemia/MDS events, with 81.1% of carriers showing rising trajectories compared to 68.5% of non-carriers. Similarly, TET2 carriers show 1.61-fold enrichment (OR=1.61, *P* = 8.9 × 10^−5^) for Signature 16 before Heart Failure events

**Figure S24:**
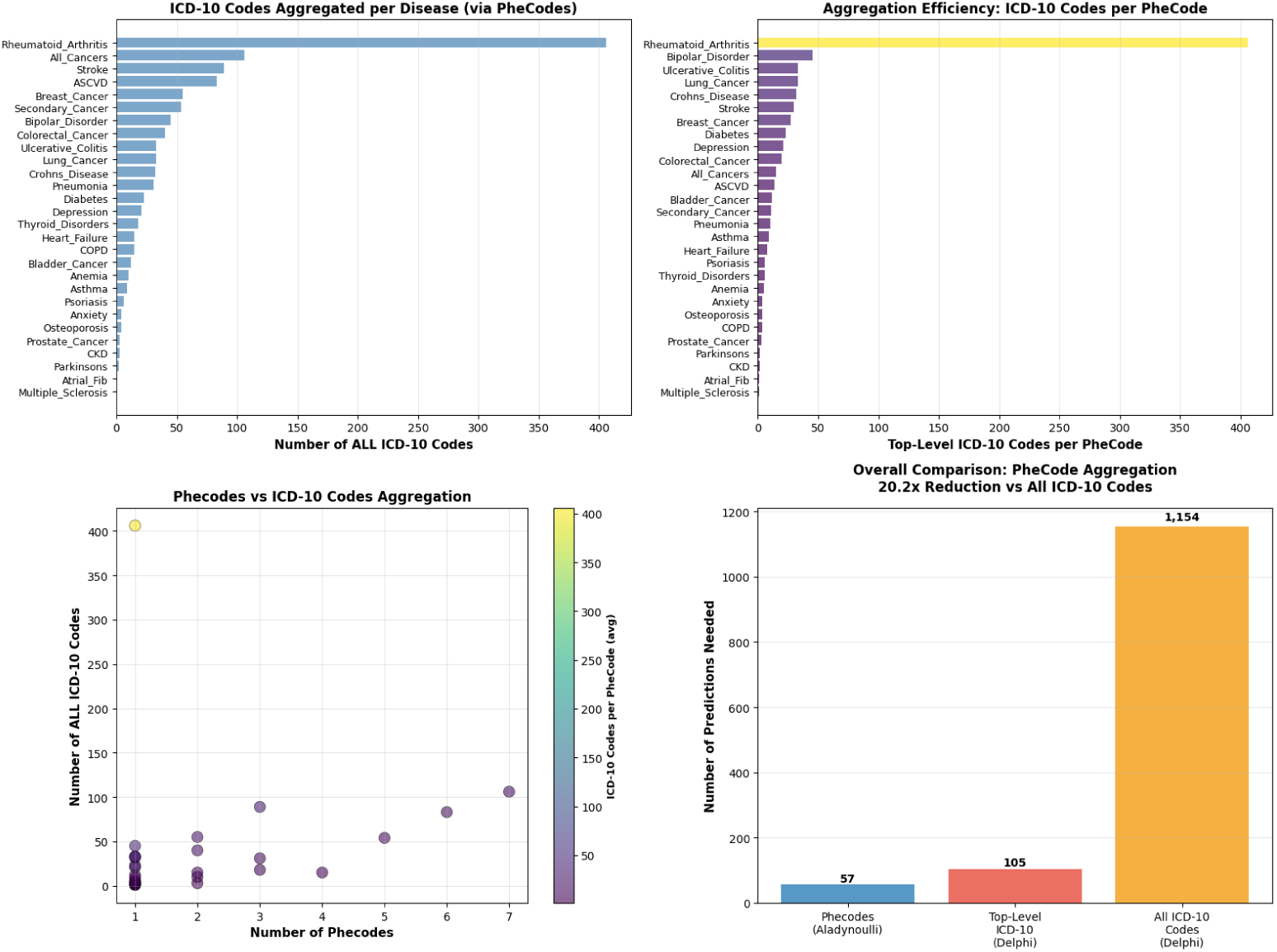
PheCode aggregation efficiency for disease-level predictions. (**Top-left**) Number of unique ICD-10 codes aggregated per disease through PheCode mapping. Diseases like Rheumatoid Arthritis show high aggregation (nearly 400 ICD-10 codes), while others have fewer codes. (**Top-right**) Aggregation efficiency measured as the number of top-level ICD-10 codes per PheCode. Rheumatoid Arthritis shows exceptionally high efficiency (400 codes per PheCode), while most diseases have lower aggregation ratios. (**Bottom-left**) Scatter plot showing the relationship between number of Phecodes and total ICD-10 codes per disease. Color indicates average ICD-10 codes per PheCode, with yellow indicating high efficiency (e.g., Rheumatoid Arthritis with 1 Phecode mapping to 400 ICD-10 codes) and purple indicating lower efficiency. Most diseases cluster in the lower-left region with 1-3 Phecodes and fewer than 100 ICD-10 codes. (**Bottom-right**) Overall comparison of prediction requirements: PheCode aggregation (Aladynoulli) requires 57 disease-level predictions, compared to 105 top-level ICD-10 predictions (Delphi) and 1,154 all ICD-10 code predictions (Delphi), demonstrating a 20.2-fold reduction in the number of predictions needed when using PheCode aggregation. This aggregation enables disease-level predictions that are clinically interpretable while dramatically reducing computational complexity compared to code-level approaches.

**Figure S25:**
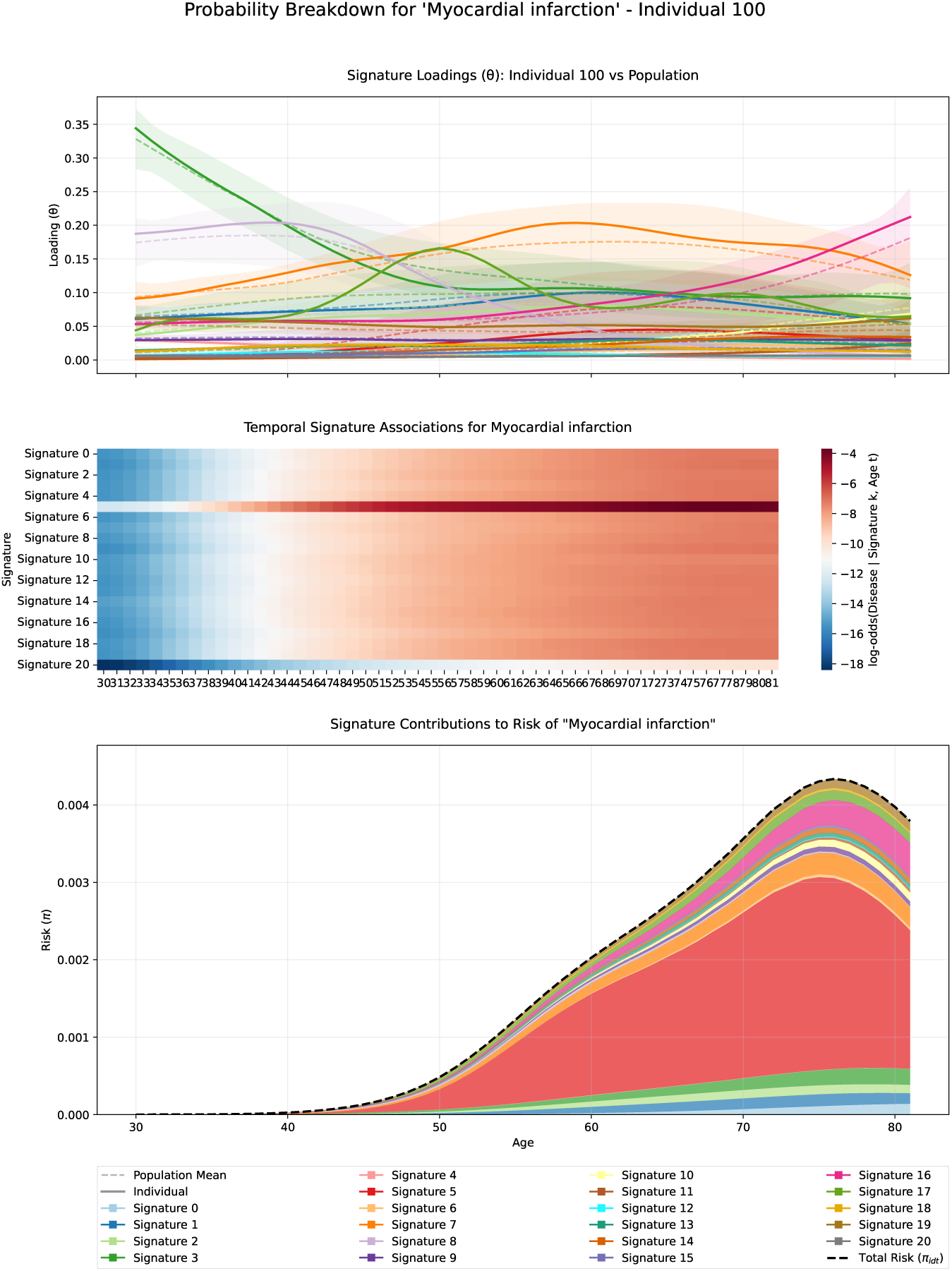
Individual and population contributions to disease probability decomposition. This figure demonstrates how population-level disease-signature associations (*ϕ*) combine with individual-specific signature loadings (*λ* transformed to *θ* via softmax) to generate personalized disease probabilities (*π*) for a representative patient (individual 100) with myocardial infarction. (**Top panel**) **Signature loadings (***θ*): Normalized individual signature loadings (solid lines) for the selected patient versus population mean loadings (dashed lines) across all 21 signatures over ages 30-81 years. Shaded regions represent one standard deviation around the population mean. Individual loadings are computed as 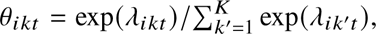 where *λ_ikt_* represents patient *i*’s latent association with signature *k* at age *t*. (**Middle panel**) **Temporal signature-disease associations (***ϕ*): Heatmap displaying the log-odds of myocardial infarction given each signature and age, computed as *ϕ_kdt_* for signature *k*, disease *d* (myocardial infarction), and timepoint *t*. Color intensity represents the strength of association (red: positive association, blue: negative association). These population-level parameters capture how disease risk varies with age within each biological signature. (**Bottom panel**) **Signature contributions to disease probability:** Stacked area plot showing how each signature contributes to the total disease probability (*π_idt_*) for this individual over time. Each colored segment represents the contribution from one signature, computed as *θ_ikt_* sigmoid *ϕ_kdt_*, scaled by the global calibration parameter *κ* so that the stacked contributions sum to the total predicted risk (black dashed line). The total probability follows Equation 1: 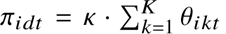 · sigmoid(*ϕ_kdt_*). This decomposition illustrates how individual signature loadings modulate population-level disease-signature associations to produce personalized risk estimates, with different signatures contributing varying amounts at different ages based on both the individual’s signature profile and age-dependent disease-signature relationships.

**Figure S26:**
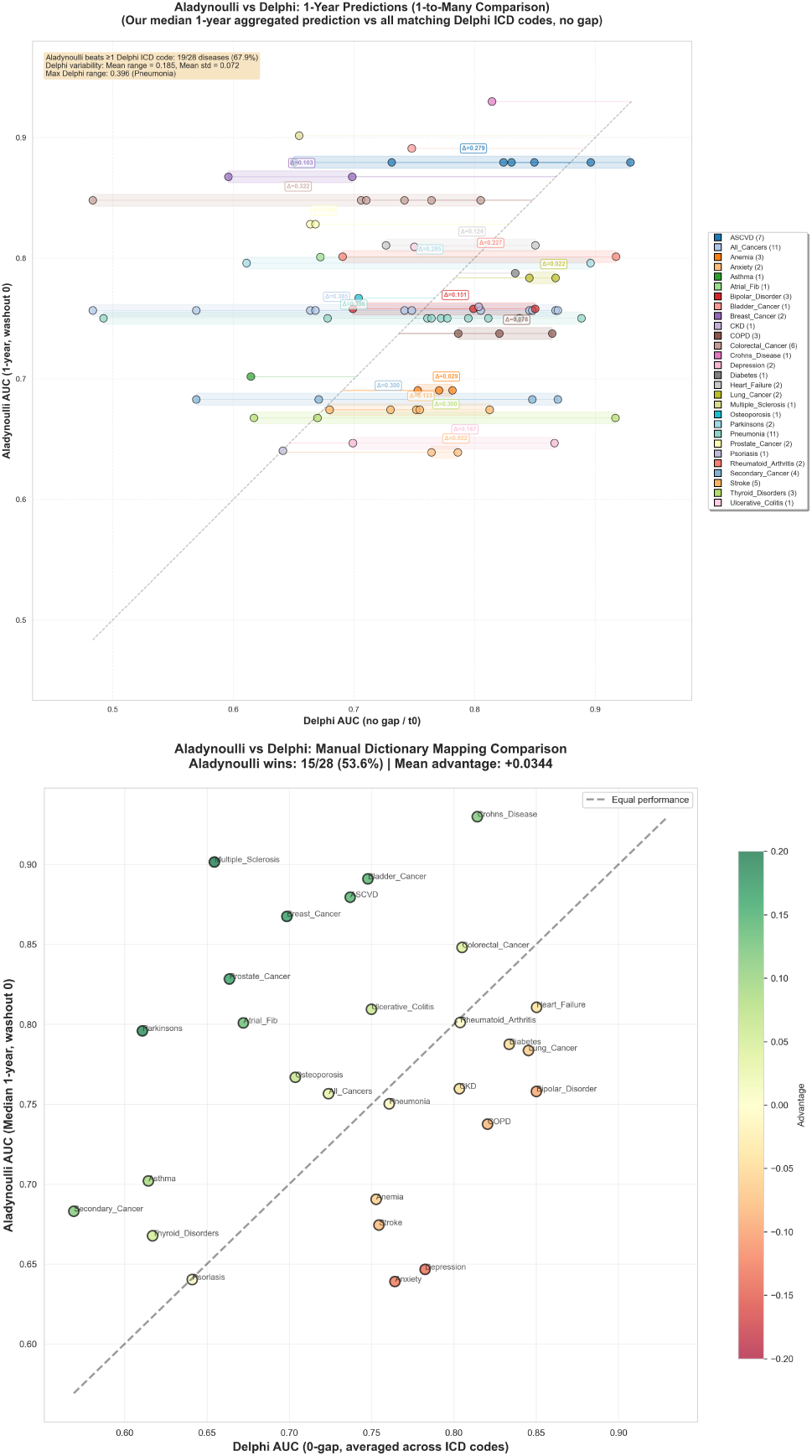
Comparison of Aladynoulli disease-level predictions versus Delphi ICD code-level predictions. **(Left panel) PheCode-based comparison:** Aladynoulli 1-year predictions (median of 1 year predictions) versus Delphi predictions aggregated across all ICD codes that map to each PheCode-defined disease. Each disease is represented by a horizontal bar showing the range of Delphi AUC values across multiple ICD codes, with a circle indicating the Aladynoulli AUC. The comparison uses the actual Phe-Code→ICD aggregation logic that our model employs, ensuring fair comparison by matching on the same disease definitions. Aladynoulli outperforms at least one Delphi ICD code for 19 out of 28 diseases (67.9), demonstrating that disease-level predictions provide superior or competitive performance compared to code-level predictions. Delphi shows substantial variability across ICD codes for the same disease (mean range = 0.185, mean std = 0.072), with Pneumonia showing the largest range (0.396), highlighting the challenge of interpreting code-level predictions. **(Right panel) Manual dictionary mapping comparison (simple mapping):** Aladynoulli versus Delphi using a manual dictionary mapping approach that matches ICD codes to diseases via simple string matching rather than PheCode aggregation. This alternative comparison method provides a baseline comparison but does not use the same PheCode→ICD aggregation logic employed by our model. Aladynoulli achieves higher AUC than Delphi for 15 out of 28 diseases (53.6), with a mean advantage of +0.0344. Color indicates the advantage (Aladynoulli AUC - Delphi AUC), with green indicating Aladynoulli advantage and red/orange indicating Delphi advantage.

**Figure S27:**
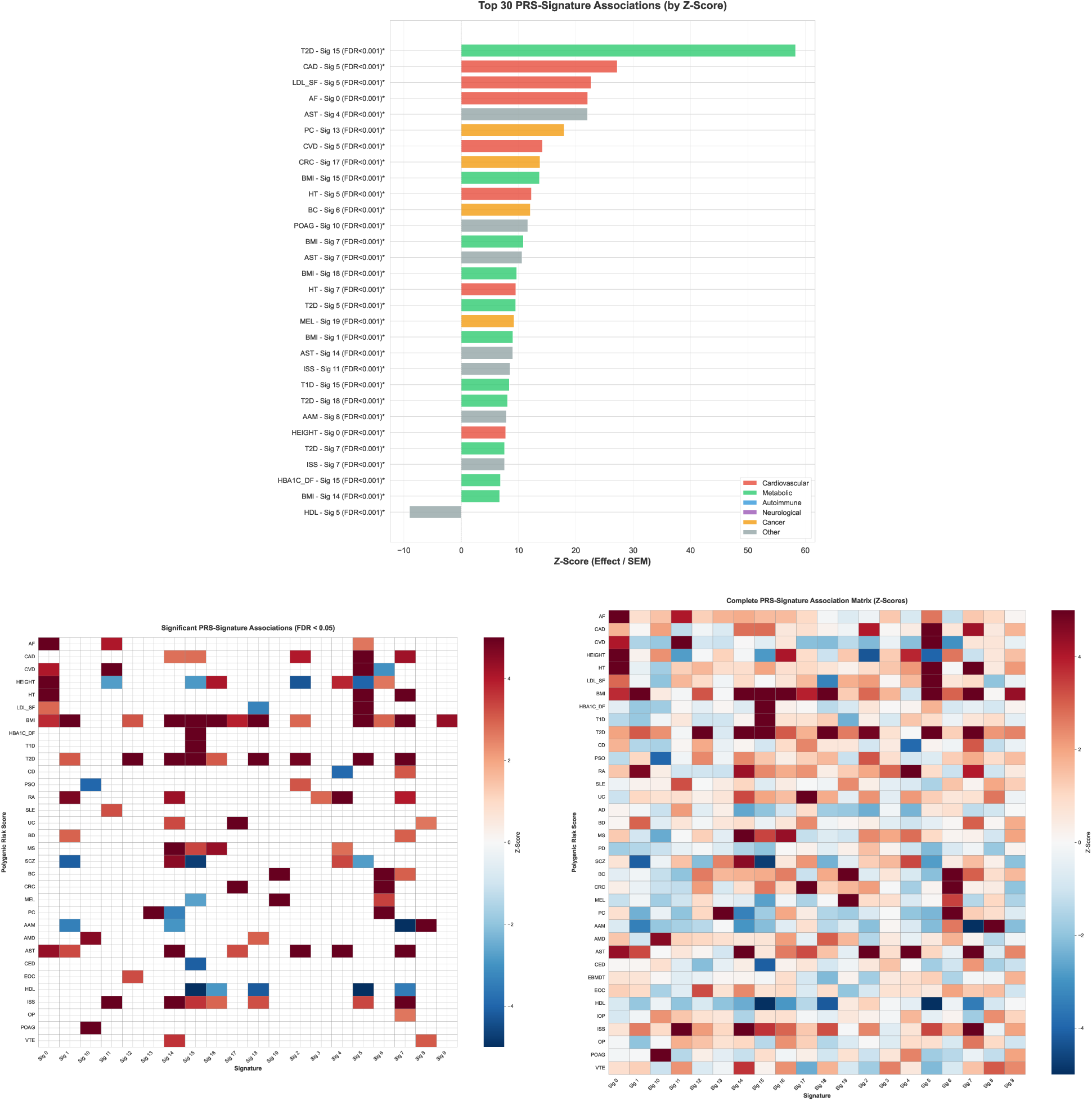
Polygenic risk score (PRS) associations with disease signatures. (**A**) Top PRS-signature associations ranked by Z-score, showing the strongest genetic effects for signatures with known heritable components. (**B**) Heatmap of significant PRS-signature associations (FDR ¡ 0.05), highlighting associations that survive multiple testing correction. (**C**) Complete PRS-signature association matrix showing all Z-scores across 36 PRS and 21 signatures. Associations between 36 external polygenic risk scores and signature loadings through the Γ*_k_* parameters model genetic effects in the framework. Z-statistics are calculated from batch-aggregated estimates across model replicates (effect size / standard error). Using Benjamini-Hochberg FDR correction, we identified 116 significant PRS-signature associations out of 756. The strongest genetic effects align with known biology: coronary artery disease PRS on the cardiovascular signature (Signature 5, *γ* = 0.153, Z = 27.2), LDL cholesterol PRS on Signature 5 (*γ* = 0.071, Z = 22.7), and type 2 diabetes PRS on the metabolic signature (Signature 15, *γ* = 0.154, Z = 58.3). PRS categories are color-coded: cardiovascular (red), metabolic (blue), neurological (green), cancer (purple), autoimmune (orange), and other (gray).

**Figure S28:**
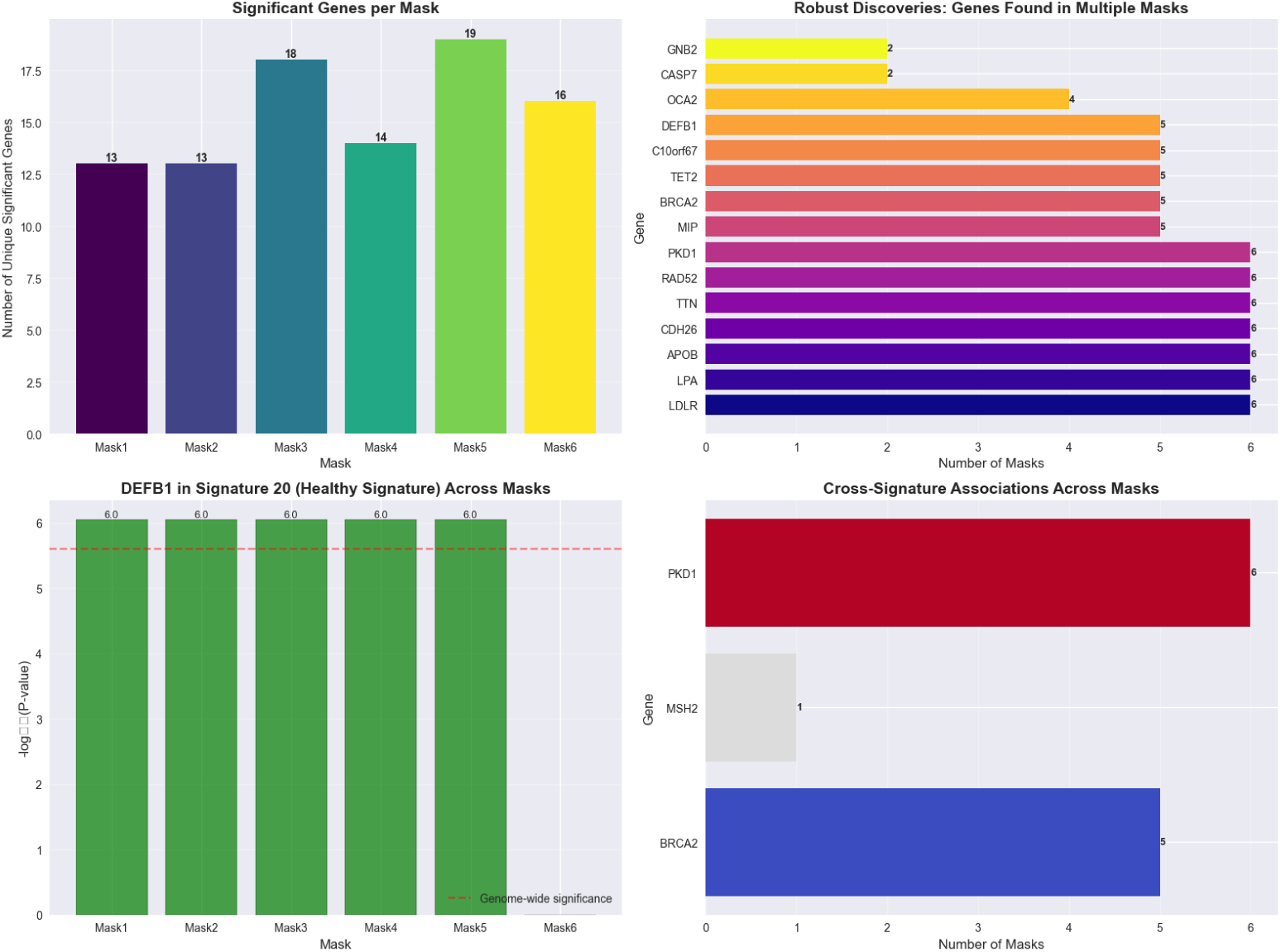
Rare variant association study (RVAS) robustness across variant filtering masks. Gene-based rare variant association studies were performed using aggregated loss-of-function (LoF) variants within genes, testing whether gene-level rare variant burden is associated with signature exposure across six different variant filtering masks (Mask1-Mask6), representing different minor allele frequency (MAF) thresholds and variant quality filters. (**Top-left**) Number of unique significant genes identified per mask, showing consistent discovery across different filtering approaches (13-19 genes per mask). (**Top-right**) Robust discoveries: genes found in multiple masks, demonstrating reproducibility across variant filtering strategies. Seven genes (LDLR, LPA, APOB, CDH26, TTN, RAD52, PKD1) are robustly discovered across all 6 masks, while additional genes (MIP, BRCA2, TET2, C10orf67, DEFB1) are found in 5 masks. This cross-mask consistency validates that these associations are not artifacts of specific variant filtering choices. (**Bottom-right**) Cross-signature associations across masks, showing genes that associate with multiple signatures. PKD1 shows cross-signature associations in all 6 masks, BRCA2 in 5 masks, demonstrating that some genes have pleiotropic effects across multiple disease signatures. Together, these panels demonstrate that rare variant associations with signatures are robust across different variant filtering approaches, validating the biological meaningfulness of the discovered gene-signature relationships.

**Figure S29:**
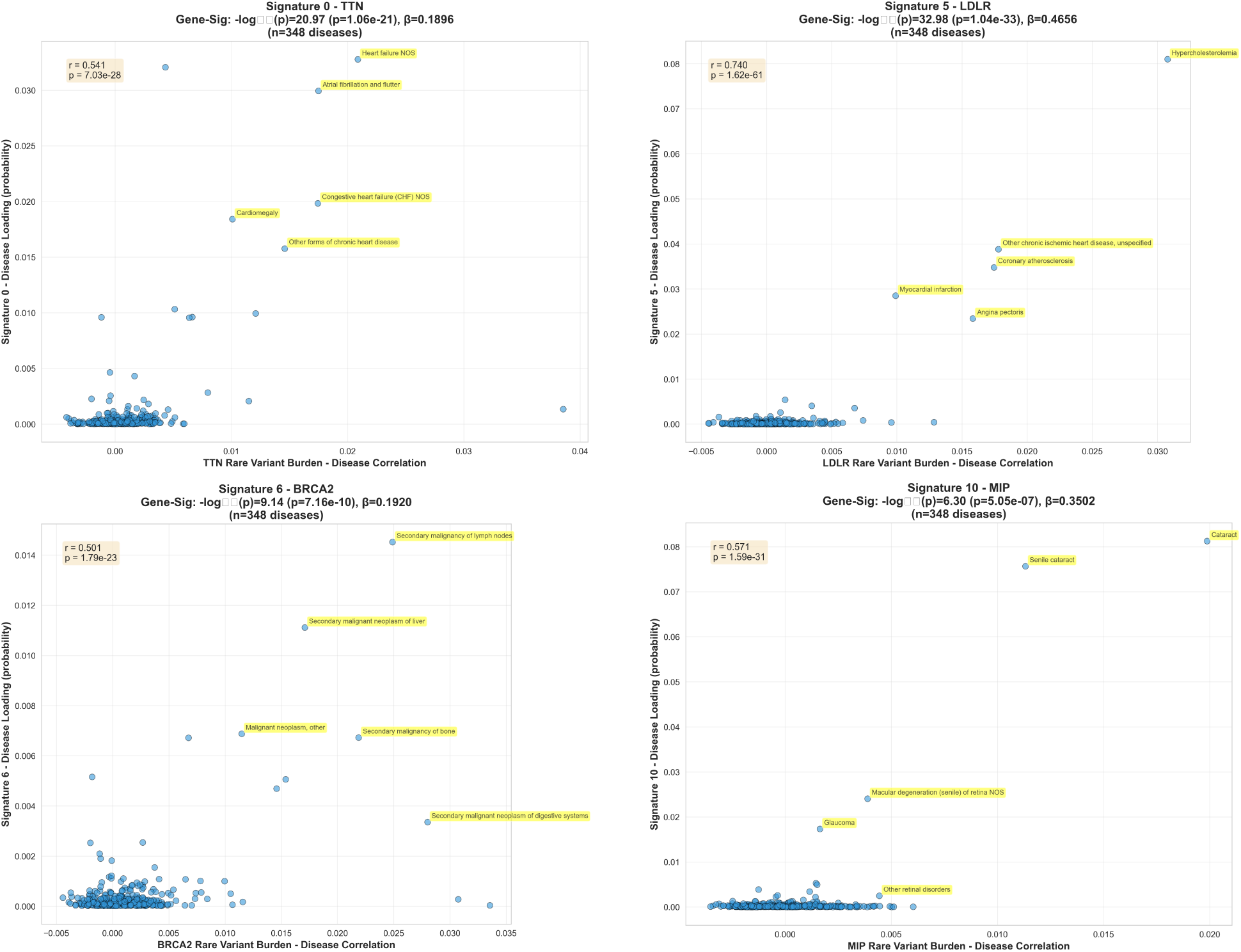
Three-way validation of rare variant associations: signature-gene, gene-disease, and disease-signature consistency. To validate rare variant associations through an independent pathway, we examined the three-way consistency between signature-gene associations (from RVAS), gene-disease correlations (from rare variant burden association with diseases), and disease-signature loadings (from *ϕ* parameters). For each signature-gene pair identified through RVAS, we tested whether diseases with high rare variant burden in that gene also showed high signature loading. Each scatter plot shows gene-disease rare variant burden correlations (x-axis) versus maximum signature-disease association over time transformed to probability scale via sigmoid (sigmoid max*_t_ ϕ_kdt_*, y-axis) for diseases with statistically significant gene-disease correlations (p *<* 0.05). (**Top-left**) Signature 0 - TTN: r = 0.535, *P* = 6.710^−17^. TTN encodes titin, a key cardiac structural protein, and shows strong associations with cardiomyopathies and arrythmias. (**Top-right**) Signature 5 - LDLR: r = 0.743, *P*= 7.1*x*10^−34^. LDLR shows the strongest three-way consistency, with diseases like hypercholesterolemia, coronary atherosclerosis, and myocardial infarction showing both high LDLR rare variant burden and high Signature 5 loading. (**Bottom-left**) Signature 6 - BRCA2: r = 0.496, p = 7.1*x*10^−16^. BRCA2 shows strong associations with secondary malignancies, particularly metastatic cancers of lymph nodes, liver, and bone, validating that Signature 6 captures cancer progression pathways. (**Bottom-right**) Signature 10 (ophthalmologic) - MIP: r = 0.577, p = 1.1*x*10^−19^. MIP shows strong associations with eye diseases including cataracts, macular degeneration, and glaucoma. The high correlations across these three measures demonstrate consistency in co-association patterns, suggesting that the rare genetic basis of signatures is reflected in both gene-disease and signature-disease relationships, without implying causal directionality.

**Figure S30:**
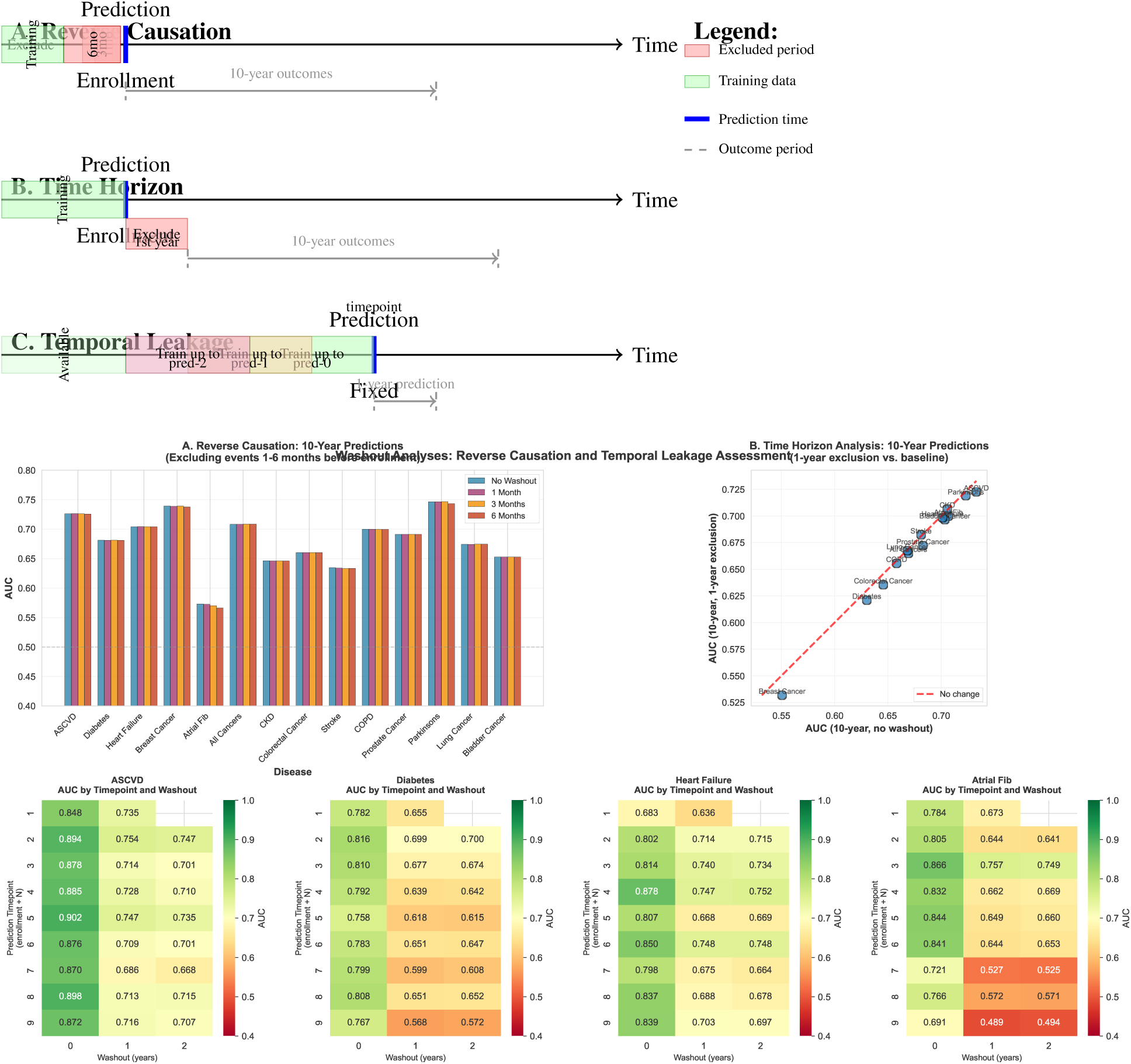
Temporal leakage and reverse causation assessment. (Top schematic) Three complementary washout approaches: (**A**) *Reverse causation*: Exclude events occurring 1, 3, or 6months before enrollment, then make 10-year predictions. This tests whether predictions rely on diagnostic cascades immediately preceding enrollment. (**B**) *Time horizon*: Exclude events in the first year after enrollment, then make 10-year predictions. This evaluates robustness to excluding the first year of outcomes. (**C**) *Temporal leakage*: Fix prediction timepoint and train models using data up to different timepoints (prediction-0, prediction-1, or prediction-2years), then make 1-year predictions. This tests for temporal information leakage by varying the amount of training data available. (Bottom panels) Results from each analysis: (Left) Reverse causation results showing minimal AUC degradation with 1-6 month exclusions. (Right) Temporal leakage heatmaps for four key diseases showing AUC performance across different prediction timepoints and washout periods.

**Figure S31:**
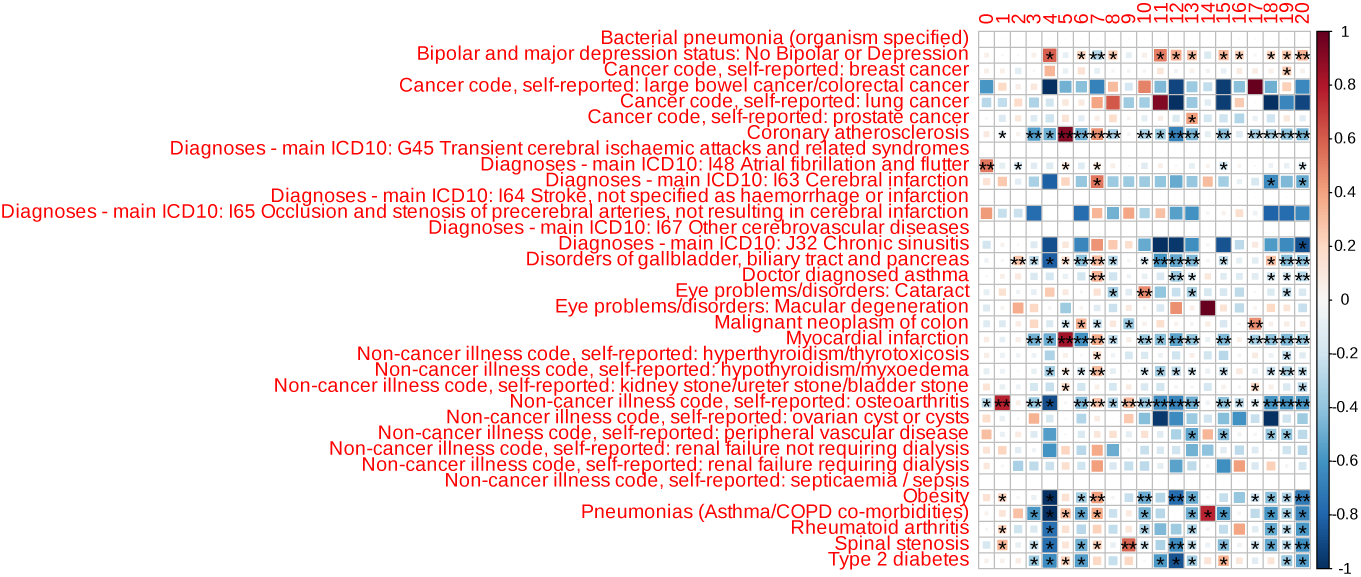
Genetic correlations between disease signatures and external complex trait GWAS. Heatmap displaying genetic correlations (*r_g_*) computed via linkage disequilibrium score regression (LDSC) between signature GWAS summary statistics and external published GWAS for a broad set of complex traits and diseases. For each signature, GWAS was performed on the area-under-the-curve (AEX) phenotype, defined as the lifetime signature exposure for each individual (integrated signature loadings over time: AEX*_ik_* = *θ_ik_ t dt*). LDSC was then applied to compute genetic correlations between each signature’s GWAS summary statistics and external trait GWAS summary statistics from published studies. The heatmap shows signatures (rows) versus external traits (columns), with color intensity representing the genetic correlation *r_g_* (red: positive correlation, blue: negative correlation, white: no correlation). Strong positive correlations (dark red) indicate shared genetic architecture between a signature and an external trait, demonstrating that signatures capture biologically meaningful genetic risk factors with pleiotropic effects across related conditions. For example, the cardiovascular signature (Signature 5) shows strong positive correlations with external cardiovascular trait GWAS (myocardial infarction, coronary artery disease, hypercholesterolemia), validating that this signature captures genuine cardiovascular genetic risk. Similarly, the metabolic/diabetes signature shows strong correlations with type 2 diabetes GWAS, while signatures show expected depletion (blue/white) for unrelated traits, confirming the biological specificity of each signature. These genetic correlations provide independent validation that disease signatures represent distinct biological pathways with shared genetic architectures, rather than statistical artifacts, and demonstrate the utility of signature-based phenotypes for understanding pleiotropic genetic effects across related conditions.

**Figure S32:**
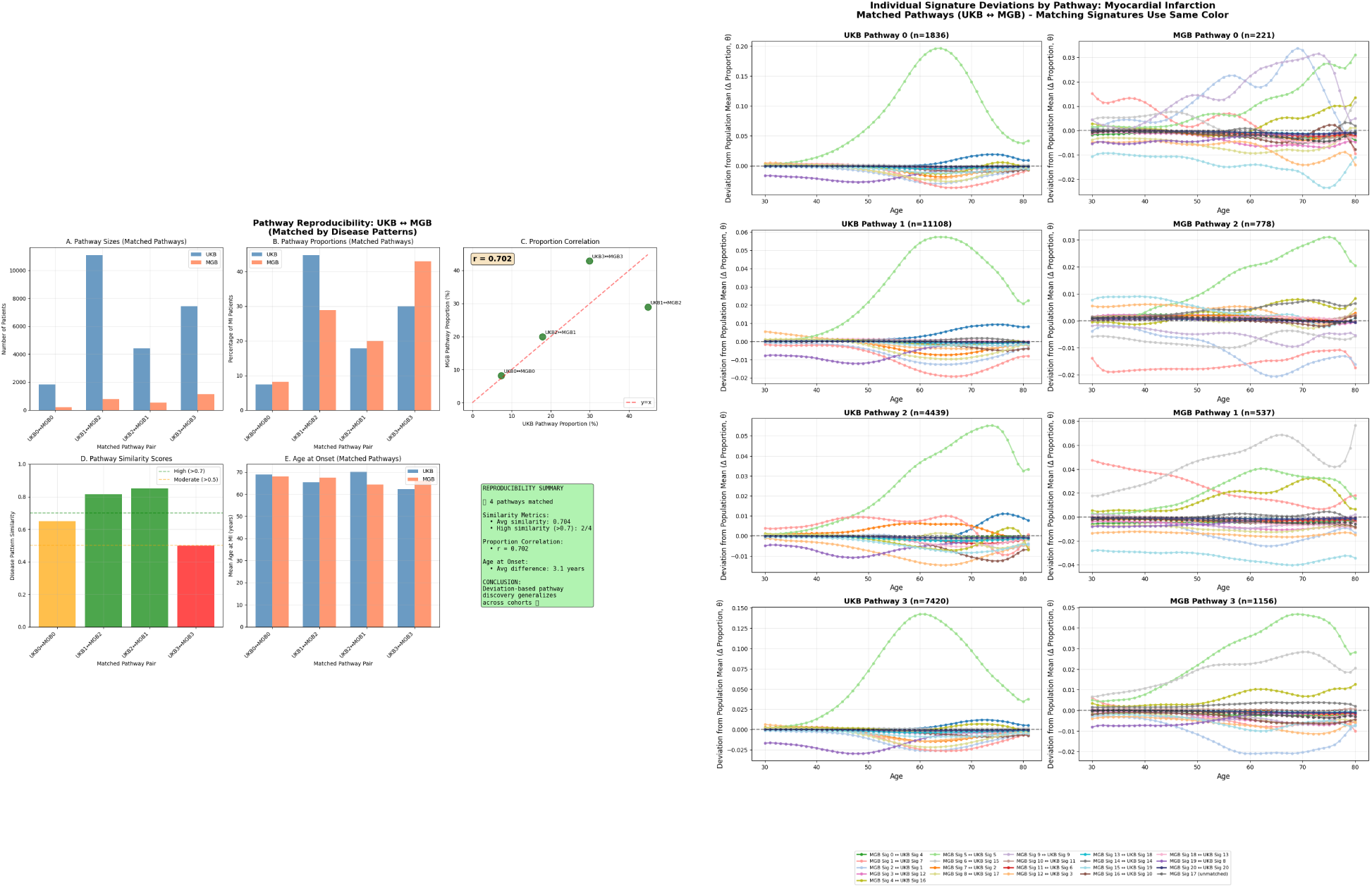
Cross-cohort validation of deviation-based pathway discovery for myocardial infarction. To further demonstrate biological heterogeneity within disease categories, we performed a supplementary pathway analysis using deviation-from-reference clustering, which complements our main analysis based on average signature loadings (**Figure 3C**). For each patient with myocardial infarction, we computed signature deviations from the population reference over the 10-year window preceding their MI event: *δ_ik_ t* = *θ_ik_ t θ̄_k_ t*, where *θ_ik_ t* is patient *i*’s signature *k* loading at age *t*and *θ̄_k_ t* is the population mean signature *k* loading at age *t*. We then applied k-means clustering (k=4) to these deviation vectors (210 features: 21 signatures × 10 timepoints), identifying four illustrative pathways that represent distinct biological routes to MI: Pathway 0 (7.4% of patients, n=1,836 UKB), Pathway 1 (44.8%, n=11,108), Pathway 2 (17.9%, n=4,439), and Pathway 3 (29.9%, n=7,420). These pathways are illustrative examples of heterogeneity rather than definitive categories, demonstrating how patients with the same diagnosis can follow distinct biological trajectories. (**Left panel**) **Pathway reproducibility metrics:** Five sub-panels comparing matched pathways between UK Biobank (UKB, blue bars) and Mass General Brigham (MGB, orange bars): (**A**) Pathway sizes (number of patients), (**B**) Pathway proportions (percentage of all MI patients), (**C**) Proportion correlation scatter plot (r = 0.702), (**D**) Disease pattern similarity scores (average 0.704, with 3/4 pathways showing high similarity ≥0.7), and (**E**) Mean age at MI onset. Pathway matching was performed based on disease pattern similarity and signature deviation profiles. (**Right panel**) **Individual signature deviations by pathway:** Line plots showing mean signature deviations (Δ Proportion, *δ*) from population reference over age (30-80 years) for matched pathways in UKB (left column) and MGB (right column). Each colored line represents one of 21 signatures, with matching signatures using the same color across cohorts to enable direct comparison. Signature 5 (cardiovascular, bright green) consistently shows strong positive deviations, particularly in Pathways 0 and 3, with peak deviations around age 60-70 years. The consistent patterns across independent cohorts validate that these deviation-based pathways capture reproducible biological heterogeneity rather than cohort-specific artifacts.

**Figure S33:**
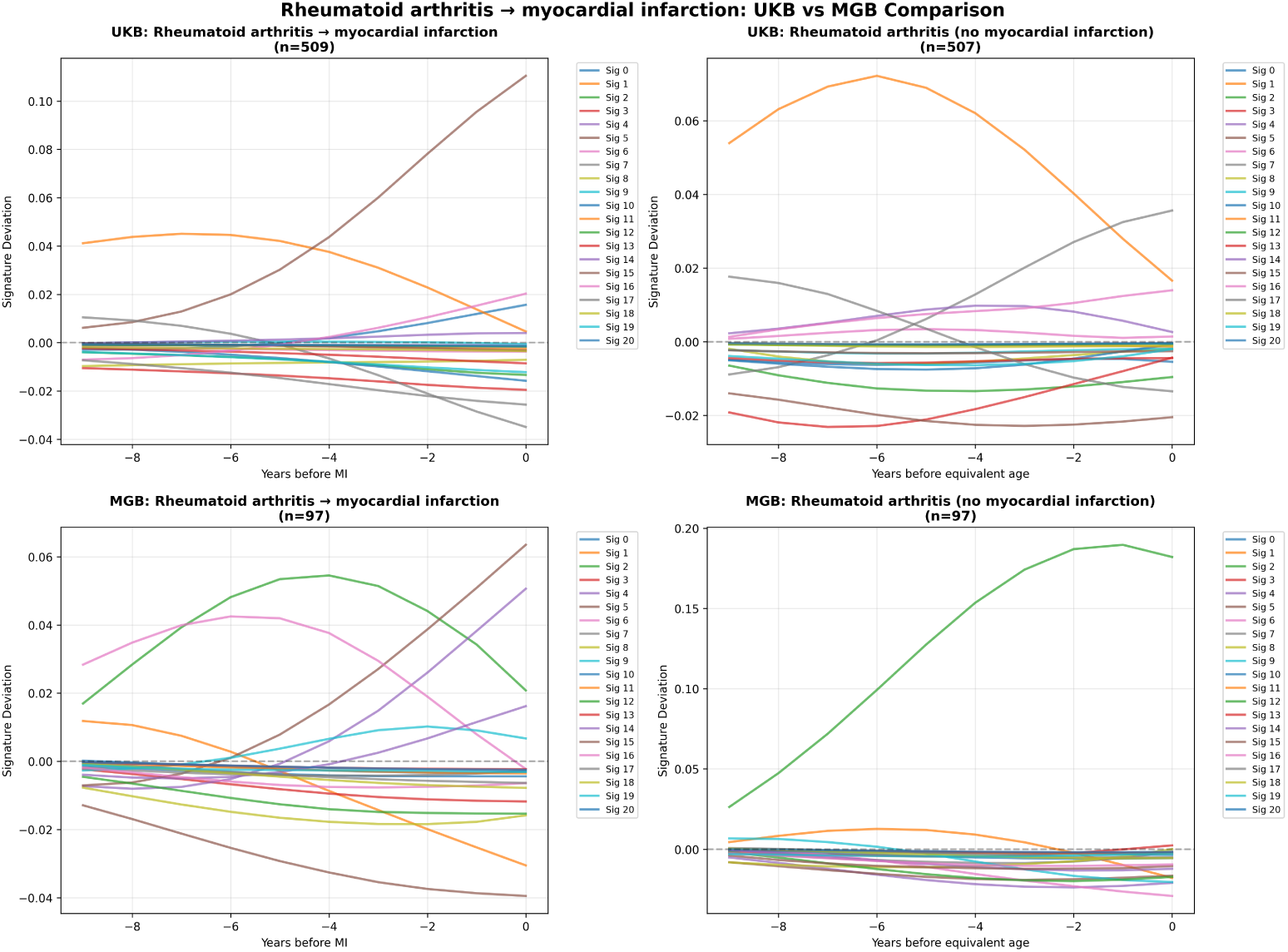
Temporal signature deviation trajectories for rheumatoid arthritis patients who develop myocardial infarction. This analysis demonstrates how signature deviations from population reference differ between RA patients who develop MI (cases) versus those who do not (controls), comparing UK Biobank (UKB) and Mass General Brigham (MGB) cohorts. For each patient with rheumatoid arthritis, signature deviations were computed as *δ_ik_ t* = *θ_ik_ t θ̄_k_ t* over the 10-year window preceding their MI event (or matched timepoint for controls), where *θ_ik_ t* represents patient *i*’s signature *k* loading at age *t*and *θ̄_k_ t* is the population mean. Mean deviations were then calculated separately for RA patients who developed MI versus RA patients who did not, for each signature and timepoint within the 10-year pre-event window. The plots show how signature deviations evolve over this pre-event period, with Signature 5 (cardiovascular, shown as a prominent colored line) exhibiting a pronounced positive deviation increase leading up to MI in both cohorts, while remaining relatively flat in RA patients who do not develop MI. This cross-cohort validation demonstrates that signature deviation patterns are reproducible across independent healthcare systems and coding practices, confirming that signature-based pathway identification captures fundamental biological variation in disease progression trajectories. The analysis serves as an illustrative example of how signature deviations can reveal distinct biological pathways leading to the same endpoint (MI) within a high-risk population (RA patients), demonstrating the model’s ability to identify heterogeneous progression patterns even among patients sharing both an initial condition (RA) and a subsequent outcome (MI).

### Extended Tables

**Table S1:** Notation and Dimensions. Mathematical notation and parameter definitions used throughout the `ALADYNOULLI` model. The table organizes symbols by category (dimensions, data structures, model parameters, and hyperparameters) to provide a comprehensive reference for the model’s mathematical framework. Each symbol is defined with its corresponding dimension, data type, and role in the model architecture.

| Symbol | Description | Dimension | Type |
| --- | --- | --- | --- |
| *Dimensions* |  |  |  |
| $N$ | Number of individuals | | |
| $D$ | Number of diseases | | |
| $T$ | Number of time points | | |
| $K$ | Number of signatures | | |
| $P$ | Number of covariates (47: 36 PRS + sex + 10 PCs) | | |
| *Data* |  |  |  |
| $\mathbf{Y}$ | Disease indicator tensor | $N \times D \times T$ | Binary |
| $\mathbf{g}_i$ | Covariate (genetic/demographic) vector for individual $i$ | $P$ | Real |
| $\mathbf{E}$ | Event/censoring time matrix | $N \times D$ | Integer |
| *Model Parameters* |  |  |  |
| $\mathbf{\Pi}$ | Disease probability tensor | $N \times D \times T$ | $[0, 1]$ |
| $\mathbf{\Theta}$ | Normalized loadings (softmax of $\mathbf{\Lambda}$) | $N \times K \times T$ | $[0, 1]$ |
| $\mathbf{\Lambda}$ | Latent signature loadings | $N \times K \times T$ | Real |
| $\mathbf{\Phi}$ | Disease-signature association | $K \times D \times T$ | Real |
| $\mathbf{\Psi}$ | Static signature-disease strength | $K \times D$ | Real |
| $\mu_d$ | Disease baseline trajectory for disease $d$ | $T$ | Real |
| $\gamma_k$ | Covariate effects for signature $k$ | $P$ | Real |
| $\kappa$ | Global calibration parameter | Scalar | $\mathbb{R}^+$ |
| *Hyperparameters* |  |  |  |
| $\alpha_\lambda$ | Amplitude for Gaussian Process on $\lambda$’s | Scalar | $\mathbb{R}^+$ |
| $l_\lambda$ | Length scale for Gaussian Process on $\lambda$’s | Scalar | $\mathbb{R}^+$ |
| $\alpha_\phi$ | Amplitude for Gaussian Process on $\phi$’s | Scalar | $\mathbb{R}^+$ |
| $l_\phi$ | Length scale for Gaussian Process on $\phi$’s | Scalar | $\mathbb{R}^+$ |
| $\sigma_\gamma^2$ | Variance of covariate effects | Scalar | $\mathbb{R}^+$ |

**Table S2:** Baseline characteristics vary across the three study cohorts. The table presents demographic and clinical characteristics for each cohort (MGB, AoU, UKB), including recruitment age, sex distribution, genetic ancestry composition, and healthcare utilization patterns. Each column represents a different cohort, with values shown as mean (SD) for continuous variables and n (%) for categorical variables. The genetic ancestry categories (EUR, AFR, AMR, EAS, SAS) reflect the population diversity across cohorts, while healthcare utilization metrics (number of diagnoses, median age at diagnosis) indicate differences in data availability and clinical patterns.

| Variable | MGB (N=48,069) | AoU (N=208,263) | UKB (N=427,239) |
| --- | --- | --- | --- |
| recruitment Age (years) | 54.3 (17.1) | 59.4 (14.3) | 57.2 (8.0) |
| Female, n (%) | 26,295 (55.4%) | 128,082 (61.5%) | 233,205 (54.6%) |
| *Genetic Ancestry* |  |  |  |
| EUR | 32,811 (69.2%) | 84,164 (40.4%) | 387,119 (90.6%) |
| AFR | 2,113 (4.5%) | 28,651 (13.7%) | 7,158 (1.7%) |
| AMR | 2,785 (5.9%) | 21,401 (10.3%) | 607 (0.1%) |
| EAS | 770 (1.6%) | 2,313 (1.1%) | 1,715 (0.4%) |
| SAS | 425 (0.9%) | 1,147 (0.6%) | 7,915 (1.9%) |
| NA (Missing) | 8,524 (18.0%) | 58,410 (28.0%) | 22,727 (5.3%) |
| Number of Diagnoses, median [IQR] | 28.0 [38.0] | 18.0 [28.0] | 6.0 [9.0] |
| Median Age at Diagnosis, median [IQR] | 58.0 [26.0] | 54.0 [22.0] | 63.3 [15.2] |
| Ages Considered | 30–81 | 30–81 | 30–81 |
| EHR Years Available | 1991–2023 | 1990–2023 | 1981–2023 |
| Average number of patients per age-year bin | 910 | 6,223 | 7,843 |

**Table S3:** Disease signatures identified by ALADYNOULLI show distinct clinical domains. The table lists all 20 disease signatures (excluding the health signature, signature 20) discovered by the model, showing representative associated diseases ranked by average signature-disease association strength (Avg PSI), the total number of diseases in each signature, and the signature name based on the primary clinical domain. Diseases are ranked by their average PSI values across model replicates, with the top 3 diseases shown for each signature.

| Sig | Representative Diseases (Top by Avg PSI) [Total: N diseases] | Signature Name |
| --- | --- | --- |
| 0 | Pleurisy; pleural effusion, Other forms of chronic heart disease, Cardiomegaly [16] | Cardiac Arrhythmias |
| 1 | Osteoarthritis (localized, primary), Arthropathy NOS, Osteoarthritis (localized) [21] | Musculoskeletal |
| 2 | Benign neoplasm of other parts of digestive system, Iron deficiency anemias, Barrett's esophagus [15] | Upper GI/Esophageal |
| 3 | Viral infection, Arrhythmia (cardiac) NOS, Facial nerve disorders [82] | Mixed/General Medical |
| 4 | Other upper respiratory disease, Chronic sinusitis, Nasal polyps [5] | Upper Respiratory |
| 5 | Hypercholesterolemia, Coronary atherosclerosis, Unstable angina [7] | Ischemic Cardiovascular |
| 6 | Malignant neoplasm (other), Secondary malignant neoplasm of liver, Secondary malignant neoplasm of digestive systems [8] | Metastatic Cancer |
| 7 | Other disorders of soft tissues, Spondylosis and allied disorders, Hyperlipidemia [22] | Pain/Inflammation |
| 8 | Prolapse of vaginal walls, Uterine leiomyoma, Benign neoplasm of ovary [28] | Gynecologic |
| 9 | Displacement of intervertebral disc, Back pain, Thoracic or lumbosacral neuritis or radiculitis [12] | Spinal Disorders |
| 10 | Macular degeneration (senile) of retina NOS, Myopia, Retinal vascular changes and abnormalities [11] | Ophthalmologic |
| 11 | Cerebral artery occlusion (with cerebral infarction), Cerebral ischemia, Cerebrovascular disease [8] | Cerebrovascular |
| 12 | Renal colic, Calculus of ureter, Hydronephrosis [7] | Renal/Urologic |
| 13 | Urinary incontinence, Cancer of prostate, Cystitis [13] | Male Urogenital |
| 14 | Pneumonia, Chronic airway obstruction, Pneumococcal pneumonia [10] | Pulmonary/Smoking |
| 15 | Hypoglycemia, Diabetic retinopathy, Type 2 diabetes with ophthalmic manifestations [5] | Metabolic/Diabetes |
| 16 | Other diseases of respiratory system (NEC), Disorders of calcium/phosphorus metabolism, Hyperkalemia [29] | Infectious/Critical Care |
| 17 | Diverticulosis, Noninfectious gastroenteritis, Hemorrhage of rectum and anus [17] | Lower GI/Colon |
| 18 | Other disorders of biliary tract, Calculus of bile duct, Cholelithiasis [9] | Hepatobiliary |
| 19 | Other non-epithelial cancer of skin, Malignant neoplasm of female breast, Benign neoplasm of skin [23] | Dermatologic/Oncologic |

**Table S4:** Signature 5 loci not detected in single-trait GWAS (exact matching). The 23 genome-wide significant loci in Signature 5 that were not found as lead loci in any constituent trait GWAS (Angina, Coronary Atherosclerosis, Hypercholesterolemia, Myocardial Infarction, Acute IHD, Chronic IHD) using exact SNP matching. These loci were detected in the signature GWAS but not in our corresponding single-trait GWAS analyses.

| Rank | Gene | rsID/Position | Chr | Position |
| --- | --- | --- | --- | --- |
| 1 | PDGFD | rs1384705 | 11 | 103696851 |
| 2 | ZNF259 | rs964184 | 11 | 116648917 |
| 3 | RP11-203J5.3 | — | — | — |
| 4 | CFDP1 | rs7192155 | 16 | 75382194 |
| 5 | SCARB1 | rs11057839 | 12 | 125316031 |
| 6 | FNDC3B | rs1499813 | 3 | 171760427 |
| 7 | C1S | rs112796495 | 12 | 7172665 |
| 8 | RAB23 | rs4715650 | 6 | 57098484 |
| 9 | SMAD3 | rs56375023 | 15 | 67448363 |
| 10 | ARMS2 | rs72631113 | 10 | 124213449 |
| 11 | HYOU1 | rs2509121 | 11 | 118928253 |
| 12 | EHBP1 | rs360807 | 2 | 62926859 |
| 13 | C7orf55 | rs4732365 | 7 | 139010660 |
| 14 | HLA-DOB | rs2199874 | 6 | 32769926 |
| 15 | WWP2 | rs62053262 | 16 | 69969299 |
| 16 | OPRL1 | rs8121509 | 20 | 62712053 |
| 17 | RP11-306G20.1 | rs11556924 | 7 | 129663496 |
| 18 | IL6R | rs6687726 | 1 | 154400320 |
| 19 | PLCG2 | rs35039495 | 16 | 81904375 |
| 20 | NR2F2-AS1 | rs7168222 | 15 | 96027903 |
| 21 | R3HDM2 | rs4760278 | 12 | 57771153 |
| 22 | UNC5C | 4:96088139 | 4 | 96088139 |
| 23 | LIPC | rs1532085 | 15 | 58683366 |

**Table S5:** Prediction methodologies differ in their temporal approach and data usage. The table defines five distinct prediction approaches used to evaluate `ALADYNOULLI` performance, and additionally includes the rolling interpolation. Each method is described in terms of its temporal updating strategy, data availability constraints, and evaluation framework. The methods progress from most sophisticated (dynamic updating with temporal censoring) to baseline comparisons (traditional Cox models without `ALADYNOULLI` features), providing a comprehensive evaluation of the model’s predictive capabilities across different clinical scenarios.

| Method | Definition |
| --- | --- |
| Median Alady-noulli 1-year | Dynamic 1-year risk predictions using models trained with data available up to each prediction time point, evaluated over 1-year windows. Median AUC across rolling one-year predictions is reported. |
| Aladynoulli 1-year | 1-year risk prediction made at cohort recruitment time using model trained on data available up to recruitment . Single prediction evaluated against 1-year outcomes. |
| Rolling Alady-noulli 10-year | Cumulative 10-year risk calculated as the product of yearly survival probabilities: $1 - \prod_{t=1}^{10} (1 - \pi_t)$ where each $\pi_t$ comes from the rolling 1-year predictions above. |
| Aladynoulli 10-year | 10-year risk prediction using single model trained only on data available at recruitment, without any temporal updating during follow-up, evaluated on 10 year outcomes. |
| Cox without Alady-noulli | Cox proportional hazards model using only traditional predictors (age, sex, family history when available). Baseline comparison model. |

**Table S6:** Aladynoulli model performance across different prediction horizons. AUC values are shown with 95% confidence intervals in parentheses.

| Disease | Aladynoulli<br>1yr (Baseline) | Aladynoulli<br>1yr (Median) | Aladynoulli<br>10yr |
| --- | --- | --- | --- |
| ASCVD | 0.881 (0.873–0.889) | 0.879 | 0.733 (0.730–0.736) |
| All Cancers | 0.753 (0.737–0.769) | 0.757 | 0.674 (0.670–0.677) |
| Anemia | 0.648 (0.635–0.664) | 0.690 | 0.588 (0.584–0.592) |
| Anxiety | 0.604 (0.558–0.648) | 0.639 | 0.514 (0.509–0.520) |
| Asthma | 0.690 (0.677–0.706) | 0.702 | 0.529 (0.525–0.533) |
| Atrial Fibrillation | 0.797 (0.777–0.811) | 0.801 | 0.707 (0.703–0.711) |
| Bipolar Disorder | 0.758 (0.692–0.840) | 0.758 | 0.492 (0.472–0.513) |
| Bladder Cancer | 0.825 (0.783–0.861) | 0.891 | 0.708 (0.698–0.716) |
| Breast Cancer | 0.782 (0.759–0.810) | 0.867 | 0.554 (0.548–0.560) |
| CKD | 0.651 (0.610–0.703) | 0.760 | 0.708 (0.703–0.712) |
| COPD | 0.736 (0.720–0.754) | 0.738 | 0.658 (0.655–0.662) |
| Colorectal Cancer | 0.825 (0.791–0.857) | 0.848 | 0.648 (0.641–0.655) |
| Crohn's Disease | 0.896 (0.862–0.919) | 0.930 | 0.580 (0.563–0.598) |
| Depression | 0.616 (0.591–0.643) | 0.647 | 0.484 (0.478–0.488) |
| Diabetes | 0.741 (0.728–0.757) | 0.787 | 0.651 (0.648–0.654) |
| Heart Failure | 0.769 (0.746–0.800) | 0.811 | 0.701 (0.696–0.707) |
| Lung Cancer | 0.699 (0.650–0.751) | 0.784 | 0.669 (0.663–0.677) |
| Multiple Sclerosis | 0.840 (0.783–0.902) | 0.902 | 0.591 (0.571–0.609) |
| Osteoporosis | 0.756 (0.732–0.782) | 0.767 | 0.681 (0.676–0.686) |
| Parkinson's | 0.809 (0.783–0.846) | 0.796 | 0.724 (0.715–0.735) |
| Pneumonia | 0.634 (0.612–0.659) | 0.750 | 0.644 (0.639–0.649) |
| Prostate Cancer | 0.831 (0.805–0.849) | 0.828 | 0.687 (0.682–0.693) |
| Psoriasis | 0.607 (0.548–0.667) | 0.640 | 0.546 (0.532–0.558) |
| Rheumatoid Arthritis | 0.749 (0.722–0.778) | 0.801 | 0.608 (0.601–0.614) |
| Secondary Cancer | 0.600 (0.574–0.621) | 0.683 | 0.610 (0.605–0.616) |
| Stroke | 0.653 (0.630–0.673) | 0.674 | 0.681 (0.675–0.688) |
| Thyroid Disorders | 0.678 (0.660–0.696) | 0.668 | 0.594 (0.590–0.598) |
| Ulcerative Colitis | 0.816 (0.774–0.860) | 0.809 | 0.583 (0.570–0.599) |

**Table S7:** Cox model and clinical risk score performance. AUC values are shown with 95% confidence intervals where available.

| **Disease** | **Cox**<br>**10yr** | **GAIL**<br>**1yr** | **GAIL**<br>**10yr** | **PCE**<br>**10yr** | **PREVENT**<br>**10yr** | **QRISK3**<br>**10yr** |
| --- | --- | --- | --- | --- | --- | --- |
| ASCVD | 0.634 | – | – | 0.683<br>(0.681–0.685) | 0.667<br>(0.665–0.669) | 0.702<br>(0.699–0.705) |
| Breast Cancer | 0.492 | 0.549<br>(0.529–0.567) | 0.540<br>(0.534–0.545) | – | – | – |
| Prostate Cancer | 0.519 | – | – | – | – | – |
| Colorectal Cancer | 0.521 | – | – | – | – | – |
| Heart Failure | 0.592 | – | – | – | – | – |
| Diabetes | 0.600 | – | – | – | – | – |
| Atrial Fibrillation | 0.588 | – | – | – | – | – |
| CKD | 0.529 | – | – | – | – | – |
| Stroke | 0.518 | – | – | – | – | – |
| Parkinson's | 0.534 | – | – | – | – | – |
| Rheumatoid Arthritis | 0.560 | – | – | – | – | – |
| Depression | 0.554 | – | – | – | – | – |
| All Cancers | 0.541 | – | – | – | – | – |

**Table S8:** Dynamic 10-year risk prediction using rolling interpolation. Predictions are updated annually using 1-year intervals, censored at first event, with sex adjustment. This approach demonstrates the benefit of incorporating time-varying information as it becomes available. AUC values are shown with 95% confidence intervals in parentheses.

| Disease | AUC (95% CI) | Events (Rate %) |
| --- | --- | --- |
| ASCVD | 0.836 (0.824–0.851) | 831 (8.3%) |
| All Cancers | 0.735 (0.712–0.754) | 480 (4.8%) |
| Anemia | 0.658 (0.632–0.681) | 523 (5.2%) |
| Anxiety | 0.573 (0.542–0.608) | 241 (2.4%) |
| Asthma | 0.612 (0.592–0.630) | 606 (6.3%) |
| Atrial Fibrillation | 0.781 (0.758–0.811) | 376 (3.8%) |
| Bipolar Disorder | 0.624 (0.530–0.725) | 34 (0.3%) |
| Bladder Cancer | 0.850 (0.787–0.901) | 49 (0.5%) |
| Breast Cancer | 0.767 (0.727–0.803) | 214 (4.0%) |
| CKD | 0.737 (0.705–0.774) | 207 (2.1%) |
| COPD | 0.715 (0.688–0.744) | 394 (3.9%) |
| Colorectal Cancer | 0.791 (0.741–0.840) | 105 (1.1%) |
| Crohn's Disease | 0.737 (0.648–0.824) | 31 (0.3%) |
| Depression | 0.546 (0.520–0.570) | 405 (4.1%) |
| Diabetes | 0.725 (0.703–0.750) | 581 (5.8%) |
| Heart Failure | 0.779 (0.740–0.804) | 205 (2.1%) |
| Lung Cancer | 0.741 (0.674–0.799) | 75 (0.8%) |
| Multiple Sclerosis | 0.690 (0.583–0.794) | 21 (0.2%) |
| Osteoporosis | 0.707 (0.668–0.735) | 219 (2.2%) |
| Parkinson's | 0.787 (0.730–0.856) | 46 (0.5%) |
| Pneumonia | 0.740 (0.710–0.766) | 335 (3.4%) |
| Prostate Cancer | 0.786 (0.750–0.827) | 204 (4.5%) |
| Psoriasis | 0.508 (0.425–0.596) | 40 (0.4%) |
| Rheumatoid Arthritis | 0.707 (0.658–0.754) | 123 (1.2%) |
| Secondary Cancer | 0.664 (0.627–0.704) | 276 (2.8%) |
| Stroke | 0.663 (0.605–0.710) | 129 (1.3%) |
| Thyroid Disorders | 0.637 (0.618–0.661) | 479 (4.8%) |
| Ulcerative Colitis | 0.793 (0.715–0.861) | 50 (0.5%) |

**Table S9:** Comparison of Aladynoulli disease-level predictions versus Delphi ICD code-level predictions. Each Aladynoulli disease (defined via PheCode aggregation) is compared against multiple ICD-10 codes that Delphi predicts separately. This demonstrates that disease-level predictions provide superior clinical interpretability while maintaining competitive or better predictive performance compared to ICD code-level predictions.

| Disease | Aladynoulli<br>1yr AUC | Delphi<br>AUC | ICD-10<br>Code | Delphi Name |
| --- | --- | --- | --- | --- |
| ASCVD | 0.879 | 0.650 | I21 | Acute myocardial infarction |
| ASCVD | 0.879 | 0.731 | I20 | Angina pectoris |
| ASCVD | 0.879 | 0.824 | I25 | Chronic ischaemic heart disease |
| ASCVD | 0.879 | 0.830 | I51 | Complications and ill-defined descriptions |
| ASCVD | 0.879 | 0.849 | I24 | Other acute ischaemic heart diseases |
| ASCVD | 0.879 | 0.896 | I23 | Current complications following MI |
| ASCVD | 0.879 | 0.929 | I22 | Subsequent myocardial infarction |
| All Cancers | 0.757 | 0.483 | C26 | Malignant neoplasm of other digestive organs |
| All Cancers | 0.757 | 0.569 | C79 | Secondary malignant neoplasm of other sites |
| All Cancers | 0.757 | 0.664 | C61 | Malignant neoplasm of prostate |
| All Cancers | 0.757 | 0.668 | D07 | Carcinoma in situ of other genital organs |
| All Cancers | 0.757 | 0.742 | D01 | Carcinoma in situ of other digestive organs |
| All Cancers | 0.757 | 0.748 | C67 | Malignant neoplasm of bladder |
| All Cancers | 0.757 | 0.805 | C18 | Malignant neoplasm of colon |
| All Cancers | 0.757 | 0.845 | C34 | Malignant neoplasm of bronchus and lung |
| All Cancers | 0.757 | 0.848 | C78 | Secondary malignant neoplasm of respiratory/digestive |
| All Cancers | 0.757 | 0.867 | D02 | Carcinoma in situ of middle ear and respiratory system |
| All Cancers | 0.757 | 0.869 | J91 | Pleural effusion in conditions classified elsewhere |
| Anemia | 0.690 | 0.753 | D50 | Iron deficiency anaemia |
| Anemia | 0.690 | 0.770 | D64 | Other anaemias |
| Anemia | 0.690 | 0.782 | D61 | Other aplastic anaemias |
| Anxiety | 0.639 | 0.764 | F41 | Other anxiety disorders |
| Anxiety | 0.639 | 0.786 | F06 | Other mental disorders due to brain damage |
| Asthma | 0.702 | 0.614 | J45 | Asthma |
| Atrial Fibrillation | 0.801 | 0.672 | I48 | Atrial fibrillation and flutter |
| Bipolar Disorder | 0.758 | 0.699 | F32 | Depressive episode |
| Bipolar Disorder | 0.758 | 0.799 | F30 | Manic episode |
| Bipolar Disorder | 0.758 | 0.850 | F31 | Bipolar affective disorder |
| Bladder Cancer | 0.891 | 0.748 | C67 | Malignant neoplasm of bladder |
| Breast Cancer | 0.867 | 0.596 | D05 | Carcinoma in situ of breast |
| Breast Cancer | 0.867 | 0.699 | C50 | Malignant neoplasm of breast |
| CKD | 0.760 | 0.803 | N18 | Chronic renal failure |
| COPD | 0.738 | 0.786 | J98 | Other respiratory disorders |
| COPD | 0.738 | 0.820 | J44 | Other chronic obstructive pulmonary disease |
| COPD | 0.738 | 0.864 | J43 | Emphysema |
| Colorectal Cancer | 0.848 | 0.483 | C26 | Malignant neoplasm of other digestive organs |
| Colorectal Cancer | 0.848 | 0.706 | C21 | Malignant neoplasm of anus and anal canal |
| Colorectal Cancer | 0.848 | 0.710 | C20 | Malignant neoplasm of rectum |
| Colorectal Cancer | 0.848 | 0.742 | D01 | Carcinoma in situ of other digestive organs |
| Colorectal Cancer | 0.848 | 0.764 | C19 | Malignant neoplasm of rectosigmoid junction |
| Colorectal Cancer | 0.848 | 0.805 | C18 | Malignant neoplasm of colon |

**Table S9 – continued from previous page**
| **Disease** | **Aladynoulli**<br>**1yr AUC** | **Delphi**<br>**AUC** | **ICD-10**<br>**Code** | **Delphi Name** |
| --- | --- | --- | --- | --- |
| Crohn's Disease | 0.930 | 0.814 | K50 | Crohn's disease [regional enteritis] |
| Depression | 0.647 | 0.699 | F32 | Depressive episode |
| Depression | 0.647 | 0.866 | F33 | Recurrent depressive disorder |
| Diabetes | 0.787 | 0.834 | E11 | Non-insulin-dependent diabetes mellitus |
| Heart Failure | 0.811 | 0.726 | I09 | Other rheumatic heart diseases |
| Heart Failure | 0.811 | 0.850 | I50 | Heart failure |
| Lung Cancer | 0.784 | 0.845 | C34 | Malignant neoplasm of bronchus and lung |
| Lung Cancer | 0.784 | 0.867 | D02 | Carcinoma in situ of middle ear and respiratory system |
| Multiple Sclerosis | 0.902 | 0.655 | G35 | Multiple sclerosis |
| Osteoporosis | 0.767 | 0.704 | M81 | Osteoporosis without pathological fracture |
| Parkinson's | 0.796 | 0.611 | G20 | Parkinson's disease |
| Parkinson's | 0.796 | 0.896 | G21 | Secondary parkinsonism |
| Pneumonia | 0.750 | 0.492 | A20 | Plague |
| Pneumonia | 0.750 | 0.678 | J16 | Pneumonia due to other infectious organisms |
| Pneumonia | 0.750 | 0.761 | J18 | Pneumonia, organism unspecified |
| Pneumonia | 0.750 | 0.764 | J13 | Pneumonia due to streptococcus pneumoniae |
| Pneumonia | 0.750 | 0.772 | B58 | Toxoplasmosis |
| Pneumonia | 0.750 | 0.777 | A02 | Other salmonella infections |
| Pneumonia | 0.750 | 0.795 | A48 | Other bacterial diseases |
| Pneumonia | 0.750 | 0.811 | A42 | Actinomycosis |
| Pneumonia | 0.750 | 0.836 | J15 | Bacterial pneumonia, not elsewhere classified |
| Pneumonia | 0.750 | 0.838 | A31 | Infection due to other mycobacteria |
| Pneumonia | 0.750 | 0.888 | J14 | Pneumonia due to haemophilus influenzae |
| Prostate Cancer | 0.828 | 0.664 | C61 | Malignant neoplasm of prostate |
| Prostate Cancer | 0.828 | 0.668 | D07 | Carcinoma in situ of other genital organs |
| Psoriasis | 0.640 | 0.641 | L40 | Psoriasis |
| Rheumatoid Arthritis | 0.801 | 0.690 | M06 | Other rheumatoid arthritis |
| Rheumatoid Arthritis | 0.801 | 0.917 | M05 | Seropositive rheumatoid arthritis |
| Secondary Cancer | 0.683 | 0.569 | C79 | Secondary malignant neoplasm of other sites |
| Secondary Cancer | 0.683 | 0.671 | C77 | Secondary and unspecified malignant neoplasm of lymph nodes |
| Secondary Cancer | 0.683 | 0.848 | C78 | Secondary malignant neoplasm of respiratory/digestive |
| Secondary Cancer | 0.683 | 0.869 | J91 | Pleural effusion in conditions classified elsewhere |
| Stroke | 0.674 | 0.680 | G45 | Transient cerebral ischaemic attacks |
| Stroke | 0.674 | 0.730 | M47 | Spondylosis |
| Stroke | 0.674 | 0.752 | G46 | Vascular syndromes of brain |
| Stroke | 0.674 | 0.754 | I63 | Cerebral infarction |
| Stroke | 0.674 | 0.812 | I67 | Other cerebrovascular diseases |
| Thyroid Disorders | 0.668 | 0.617 | E03 | Other hypothyroidism |
| Thyroid Disorders | 0.668 | 0.670 | E05 | Thyrotoxicosis [hyperthyroidism] |
| Thyroid Disorders | 0.668 | 0.917 | E89 | Postprocedural endocrine and metabolic disorders |
| Ulcerative Colitis | 0.809 | 0.750 | K51 | Ulcerative colitis |

**Table S10:**
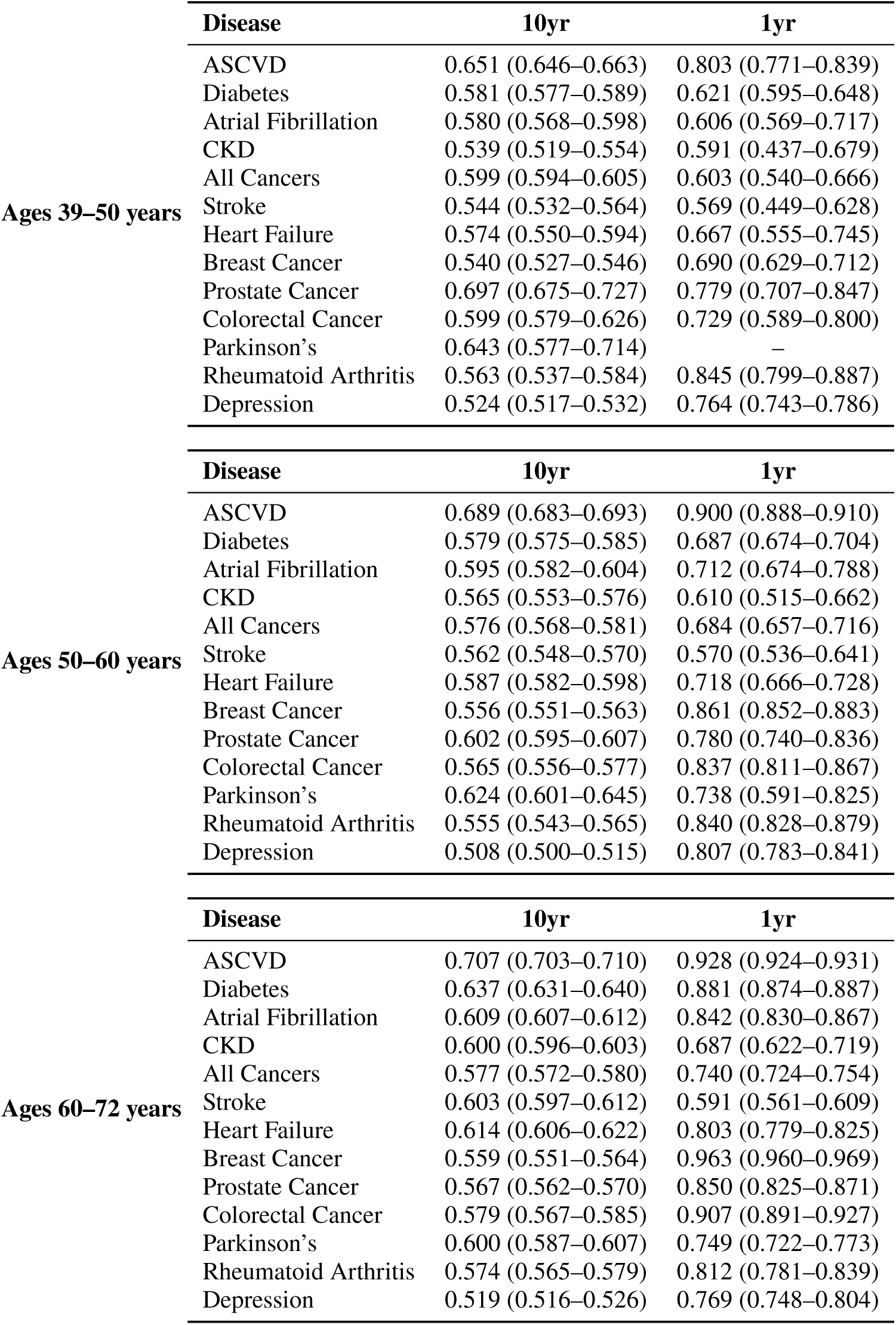
Age-stratified model performance. AUC values are shown with 95% confidence intervals in parentheses for 10-year static predictions and 1-year predictions across three age groups.

|  | **Disease** | **10yr** | **1yr** |
| --- | --- | --- | --- |
| **Ages 39–50 years** | ASCVD | 0.651 (0.646–0.663) | 0.803 (0.771–0.839) |
|  | Diabetes | 0.581 (0.577–0.589) | 0.621 (0.595–0.648) |
|  | Atrial Fibrillation | 0.580 (0.568–0.598) | 0.606 (0.569–0.717) |
|  | CKD | 0.539 (0.519–0.554) | 0.591 (0.437–0.679) |
|  | All Cancers | 0.599 (0.594–0.605) | 0.603 (0.540–0.666) |
|  | Stroke | 0.544 (0.532–0.564) | 0.569 (0.449–0.628) |
|  | Heart Failure | 0.574 (0.550–0.594) | 0.667 (0.555–0.745) |
|  | Breast Cancer | 0.540 (0.527–0.546) | 0.690 (0.629–0.712) |
|  | Prostate Cancer | 0.697 (0.675–0.727) | 0.779 (0.707–0.847) |
|  | Colorectal Cancer | 0.599 (0.579–0.626) | 0.729 (0.589–0.800) |
|  | Parkinson's | 0.643 (0.577–0.714) | – |
|  | Rheumatoid Arthritis | 0.563 (0.537–0.584) | 0.845 (0.799–0.887) |
|  | Depression | 0.524 (0.517–0.532) | 0.764 (0.743–0.786) |
|  | **Disease** | **10yr** | **1yr** |
| **Ages 50–60 years** | ASCVD | 0.689 (0.683–0.693) | 0.900 (0.888–0.910) |
|  | Diabetes | 0.579 (0.575–0.585) | 0.687 (0.674–0.704) |
|  | Atrial Fibrillation | 0.595 (0.582–0.604) | 0.712 (0.674–0.788) |
|  | CKD | 0.565 (0.553–0.576) | 0.610 (0.515–0.662) |
|  | All Cancers | 0.576 (0.568–0.581) | 0.684 (0.657–0.716) |
|  | Stroke | 0.562 (0.548–0.570) | 0.570 (0.536–0.641) |
|  | Heart Failure | 0.587 (0.582–0.598) | 0.718 (0.666–0.728) |
|  | Breast Cancer | 0.556 (0.551–0.563) | 0.861 (0.852–0.883) |
|  | Prostate Cancer | 0.602 (0.595–0.607) | 0.780 (0.740–0.836) |
|  | Colorectal Cancer | 0.565 (0.556–0.577) | 0.837 (0.811–0.867) |
|  | Parkinson's | 0.624 (0.601–0.645) | 0.738 (0.591–0.825) |
|  | Rheumatoid Arthritis | 0.555 (0.543–0.565) | 0.840 (0.828–0.879) |
|  | Depression | 0.508 (0.500–0.515) | 0.807 (0.783–0.841) |
|  | **Disease** | **10yr** | **1yr** |
| **Ages 60–72 years** | ASCVD | 0.707 (0.703–0.710) | 0.928 (0.924–0.931) |
|  | Diabetes | 0.637 (0.631–0.640) | 0.881 (0.874–0.887) |
|  | Atrial Fibrillation | 0.609 (0.607–0.612) | 0.842 (0.830–0.867) |
|  | CKD | 0.600 (0.596–0.603) | 0.687 (0.622–0.719) |
|  | All Cancers | 0.577 (0.572–0.580) | 0.740 (0.724–0.754) |
|  | Stroke | 0.603 (0.597–0.612) | 0.591 (0.561–0.609) |
|  | Heart Failure | 0.614 (0.606–0.622) | 0.803 (0.779–0.825) |
|  | Breast Cancer | 0.559 (0.551–0.564) | 0.963 (0.960–0.969) |
|  | Prostate Cancer | 0.567 (0.562–0.570) | 0.850 (0.825–0.871) |
|  | Colorectal Cancer | 0.579 (0.567–0.585) | 0.907 (0.891–0.927) |
|  | Parkinson's | 0.600 (0.587–0.607) | 0.749 (0.722–0.773) |
|  | Rheumatoid Arthritis | 0.574 (0.565–0.579) | 0.812 (0.781–0.839) |
|  | Depression | 0.519 (0.516–0.526) | 0.769 (0.748–0.804) |

**Table S11:** Disease signatures show significant heritability across multiple domains. The table presents LD score regression results for genome-wide association studies of signature trajectories, showing heritability estimates (*ℎ*^2^), genomic control inflation (*λ_GC_*), and intercept values for each of the 20 disease signatures. Each row represents a different signature, with columns showing the number of SNPs analyzed, heritability estimate with standard error, genomic control metrics, and ratio statistics. Higher heritability values indicate stronger genetic contributions to signature-specific disease risk patterns, with cardiovascular and musculoskeletal signatures showing the strongest genetic signals.

| Signature | nSNP | $h^2$ | $\lambda_{GC}$ | Intercept | Ratio |
| --- | --- | --- | --- | --- | --- |
| 0 | 930,186 | 0.0105 (0.0016) | 1.0757 | 1.0095 (0.007) | 0.109 (0.0806) |
| 1 | 930,186 | 0.0351 (0.0021) | 1.2219 | 0.9972 (0.0088) | <0 |
| 2 | 930,186 | 0.0192 (0.0017) | 1.1509 | 1.0147 (0.0072) | 0.0945 (0.0463) |
| 3 | 930,186 | 0.011 (0.0015) | 1.0885 | 1.0124 (0.0075) | 0.1319 (0.0792) |
| 4 | 930,186 | 0.0034 (0.0015) | 1.0108 | 0.9947 (0.0067) | <0 |
| 5 | 930,186 | 0.0414 (0.0032) | 1.224 | 1.0322 (0.0092) | 0.0945 (0.0271) |
| 6 | 930,186 | 0.0078 (0.0014) | 1.0556 | 1.0022 (0.0067) | 0.0358 (0.1091) |
| 7 | 930,186 | 0.0268 (0.0021) | 1.1904 | 1.0206 (0.0075) | 0.0912 (0.0332) |
| 8 | 930,186 | 0.0065 (0.0014) | 1.0409 | 1.0005 (0.0062) | 0.0096 (0.1262) |
| 9 | 930,186 | 0.009 (0.0016) | 1.0555 | 0.9968 (0.0075) | <0 |
| 10 | 930,186 | 0.0182 (0.0015) | 1.123 | 1.0029 (0.0067) | 0.0206 (0.0484) |
| 11 | 930,186 | 0.0042 (0.0013) | 1.0303 | 0.9967 (0.0065) | <0 |
| 12 | 930,186 | 0.0094 (0.0016) | 1.0673 | 0.9996 (0.0071) | <0 |
| 13 | 930,186 | 0.0129 (0.0016) | 1.0952 | 1.0128 (0.0068) | 0.1165 (0.0622) |
| 14 | 930,186 | 0.0081 (0.0016) | 1.0591 | 1.004 (0.0077) | 0.0627 (0.1205) |
| 15 | 930,186 | 0.0093 (0.0016) | 1.0742 | 1.011 (0.0074) | 0.1325 (0.0896) |
| 16 | 930,186 | 0.0041 (0.0014) | 1.0388 | 1.0072 (0.0066) | 0.1867 (0.1721) |
| 17 | 930,186 | 0.0242 (0.0021) | 1.1598 | 1.0165 (0.008) | 0.0825 (0.0397) |
| 18 | 930,186 | 0.0087 (0.0021) | 1.0642 | 1.0098 (0.0087) | 0.1281 (0.1142) |
| 19 | 930,186 | 0.0165 (0.0021) | 1.1261 | 1.0247 (0.0081) | 0.1655 (0.054) |
| 20 | 930,186 | 0.0142 (0.0016) | 1.1099 | 1.0064 (0.0074) | 0.0561 (0.0645) |

**Table S12:** LDSC heritability estimates for component CVD traits on both observed and liability scales. **Key point:** Because our signatures are continuous phenotypes, they can only be measured on the observed scale. Therefore, the observed-scale heritabilities 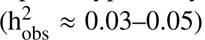 shown here are the appropriate comparison for Signature 5 (*ℎ*^2^ = 0.0414)—and Signature 5 meets or exceeds these values. Liability-scale estimates are provided solely for comparison with literature values, which typically report heritability on this scale (e.g., Neale Lab reports coronary atherosclerosis 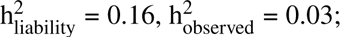 https://nealelab.github.io/UKBB_ldsc/h2_summary_I9_CORATHER.html). Conversion uses the formula from Lee et al. 2011: 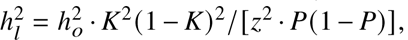 where *K* is population prevalence, *P* is sample prevalence, and *z* is the standard normal density at the liability threshold. Following standard practice when population prevalence is unknown, we assume *K* = *P*.

| Component CVD Trait | $h^2_{\text{obs}}$ (SE) | Prevalence | $h^2_{\text{liability}}$ | Intercept (SE) |
| --- | --- | --- | --- | --- |
| Angina Pectoris | 0.0340 (0.0024) | 0.072 | 0.121 | 1.033 (0.008) |
| Coronary Atherosclerosis | 0.0477 (0.0035) | 0.089 | 0.149 | 1.043 (0.009) |
| Hypercholesterolemia | 0.0444 (0.0032) | 0.178 | 0.096 | 1.050 (0.009) |
| Myocardial Infarction | 0.0316 (0.0024) | 0.062 | 0.123 | 1.024 (0.009) |
| Other Acute IHD | 0.0033 (0.0013) | 0.009 | 0.049 | 1.019 (0.007) |
| Other Chronic IHD | 0.0339 (0.0023) | 0.077 | 0.116 | 1.037 (0.009) |
| Unstable Angina | 0.0117 (0.0015) | 0.020 | 0.098 | 1.013 (0.007) |

**Table S13:** Significant rare variant associations: Mask1 (LoF variants only). Gene-based rare variant association results for Mask1, showing the best MAF threshold per gene-signature pair. LOG10P = − log_10_(p-value). Genome-wide significance threshold: p *<* 2.5 × 10^−6^ (LOG10P ≥ 5.6).

| Sig | Gene | MAF | LOG10P | Beta | SE | Chr |
| --- | --- | --- | --- | --- | --- | --- |
| 0 | TTN | 0.0001 | 20.49 | 0.1877 | 0.0198 | 2 |
| 3 | MSH2 | 0.001 | 5.96 | -0.4047 | 0.0830 | 2 |
| 4 | C10orf67 | singleton | 5.93 | 3.4043 | 0.7002 | 10 |
| 5 | LDLR | 0.0001 | 17.54 | 0.9228 | 0.1059 | 19 |
| 5 | APOB | 0.0001 | 8.28 | -0.2520 | 0.0432 | 2 |
| 5 | LPA | 0.1 | 7.14 | -0.0325 | 0.0060 | 6 |
| 5 | CDH26 | 0.001 | 6.11 | -0.1283 | 0.0260 | 20 |
| 6 | BRCA2 | 0.0001 | 9.14 | 0.1920 | 0.0312 | 13 |
| 8 | PKD1 | $1 \times 10^{-5}$ | 6.62 | -0.6135 | 0.1187 | 16 |
| 16 | PKD1 | $1 \times 10^{-5}$ | 13.73 | 0.9337 | 0.1219 | 16 |
| 16 | TET2 | singleton | 6.73 | 0.4271 | 0.0820 | 4 |
| 16 | BRCA2 | 0.0001 | 5.62 | 0.1481 | 0.0314 | 13 |
| 17 | MSH2 | $1 \times 10^{-5}$ | 5.94 | 0.6478 | 0.1332 | 2 |
| 19 | GJB2 | $1 \times 10^{-5}$ | 5.97 | 0.7586 | 0.1556 | 13 |
| 19 | RAD52 | 0.1 | 5.94 | -0.0377 | 0.0077 | 12 |
| 20 | DEFB1 | 0.0001 | 6.05 | 0.7003 | 0.1425 | 8 |

**Table S14:** Significant rare variant associations: Mask2 (LoF and DelMis09 variants). Gene-based rare variant association results for Mask2, showing the best MAF threshold per gene-signature pair. LOG10P = – log_10_(p-value). Genome-wide significance threshold: p *<* 2.5 × 10^−6^ (LOG10P ≥ 5.6).

| Sig | Gene | MAF | LOG10P | Beta | SE | Chr |
| --- | --- | --- | --- | --- | --- | --- |
| 0 | TTN | 0.0001 | 20.49 | 0.1877 | 0.0198 | 2 |
| 4 | C10orf67 | singleton | 5.93 | 3.4043 | 0.7002 | 10 |
| 5 | LDLR | 0.0001 | 18.28 | 0.8885 | 0.0998 | 19 |
| 5 | APOB | 0.0001 | 8.28 | -0.2520 | 0.0432 | 2 |
| 5 | LPA | 0.1 | 7.14 | -0.0325 | 0.0060 | 6 |
| 5 | CDH26 | 0.001 | 6.11 | -0.1283 | 0.0260 | 20 |
| 6 | BRCA2 | 0.0001 | 9.14 | 0.1920 | 0.0312 | 13 |
| 8 | PKD1 | $1 \times 10^{-5}$ | 6.62 | -0.6135 | 0.1187 | 16 |
| 10 | MIP | 0.0001 | 6.29 | 0.4722 | 0.0940 | 12 |
| 16 | PKD1 | $1 \times 10^{-5}$ | 13.73 | 0.9337 | 0.1219 | 16 |
| 16 | TET2 | singleton | 6.73 | 0.4271 | 0.0820 | 4 |
| 16 | BRCA2 | 0.0001 | 5.62 | 0.1481 | 0.0314 | 13 |
| 19 | PTPN14 | 0.0001 | 6.18 | 0.4725 | 0.0950 | 1 |
| 19 | RAD52 | 0.1 | 5.94 | -0.0377 | 0.0077 | 12 |
| 20 | DEFB1 | 0.0001 | 6.05 | 0.7003 | 0.1425 | 8 |

**Table S15:** Significant rare variant associations: Mask3 (LoF, DelMis09, DelMis08 variants). Gene-based rare variant association results for Mask3, showing the best MAF threshold per gene-signature pair. LOG10P = − log_10_(p-value). Genome-wide significance threshold: p *<* 2.5 × 10^−6^ (LOG10P ≥ 5.6).

| Sig | Gene | MAF | LOG10P | Beta | SE | Chr |
| --- | --- | --- | --- | --- | --- | --- |
| 0 | TTN | 0.0001 | 20.97 | 0.1896 | 0.0198 | 2 |
| 0 | EEF1A1 | singleton | 5.89 | -2.1361 | 0.4410 | 6 |
| 3 | ADGRG7 | 0.001 | 5.65 | -0.1780 | 0.0376 | 3 |
| 4 | C10orf67 | singleton | 5.93 | 3.4043 | 0.7002 | 10 |
| 5 | LDLR | 0.001 | 32.98 | 0.4656 | 0.0385 | 19 |
| 5 | APOB | 0.0001 | 8.28 | -0.2520 | 0.0432 | 2 |
| 5 | LPA | 0.1 | 7.14 | -0.0325 | 0.0060 | 6 |
| 5 | CDH26 | 0.001 | 6.11 | -0.1283 | 0.0260 | 20 |
| 6 | BRCA2 | 0.0001 | 9.14 | 0.1920 | 0.0312 | 13 |
| 7 | GNB2 | 0.0001 | 5.84 | 0.8585 | 0.1781 | 7 |
| 8 | RNF216 | 0.0001 | 6.28 | -0.3578 | 0.0713 | 7 |
| 10 | MIP | 0.0001 | 6.30 | 0.3502 | 0.0697 | 12 |
| 11 | CLPTM1L | 0.0001 | 6.23 | 0.4448 | 0.0890 | 5 |
| 16 | PKD1 | $1 \times 10^{-5}$ | 11.63 | 0.6068 | 0.0865 | 16 |
| 16 | TET2 | singleton | 6.73 | 0.4271 | 0.0820 | 4 |
| 16 | BRCA2 | 0.0001 | 5.62 | 0.1481 | 0.0314 | 13 |
| 19 | OCA2 | 0.01 | 6.45 | 0.1232 | 0.0242 | 15 |
| 19 | RAD52 | 0.1 | 5.99 | -0.0378 | 0.0077 | 12 |
| 20 | DEFB1 | 0.0001 | 6.05 | 0.7003 | 0.1425 | 8 |

**Table S16:** Significant rare variant associations: Mask4 (LoF, DelMis09, DelMis08, DelMis07 variants). Gene-based rare variant association results for Mask4, showing the best MAF threshold per gene-signature pair. LOG10P = − log_10_(p-value). Genome-wide significance threshold: p *<* 2.5 × 10^−6^ (LOG10P ≥ 5.6).

| Sig | Gene | MAF | LOG10P | Beta | SE | Chr |
| --- | --- | --- | --- | --- | --- | --- |
| 0 | TTN | 0.0001 | 17.96 | 0.1501 | 0.0170 | 2 |
| 4 | C10orf67 | singleton | 5.93 | 3.4043 | 0.7002 | 10 |
| 5 | LDLR | 0.001 | 26.58 | 0.3037 | 0.0281 | 19 |
| 5 | APOB | 0.0001 | 7.59 | -0.2349 | 0.0422 | 2 |
| 5 | LPA | 0.1 | 7.14 | -0.0325 | 0.0060 | 6 |
| 5 | CDH26 | 0.001 | 6.11 | -0.1283 | 0.0260 | 20 |
| 6 | BRCA2 | 0.0001 | 10.41 | 0.2017 | 0.0305 | 13 |
| 7 | GNB2 | 0.0001 | 6.03 | 0.6442 | 0.1314 | 7 |
| 10 | MIP | 0.0001 | 6.91 | 0.3361 | 0.0636 | 12 |
| 16 | PKD1 | singleton | 7.91 | 0.4927 | 0.0865 | 16 |
| 16 | TET2 | singleton | 6.23 | 0.3985 | 0.0798 | 4 |
| 16 | BRCA2 | 0.0001 | 5.92 | 0.1494 | 0.0308 | 13 |
| 19 | OCA2 | 0.01 | 10.22 | 0.0738 | 0.0113 | 15 |
| 19 | RAD52 | 0.1 | 6.11 | -0.0381 | 0.0077 | 12 |
| 20 | DEFB1 | 0.0001 | 6.05 | 0.7003 | 0.1425 | 8 |

**Table S17:** Significant rare variant associations: Mask5 (LoF, DelMis09, DelMis08, DelMis07, DelMis06 variants). Gene-based rare variant association results for Mask5, showing the best MAF threshold per gene-signature pair. LOG10P = − log_10_(p-value). Genome-wide significance threshold: p *<* 2.5 × 10^−6^ (LOG10P ≥ 5.6).

| Sig | Gene | MAF | LOG10P | Beta | SE | Chr |
| --- | --- | --- | --- | --- | --- | --- |
| 0 | TTN | 0.0001 | 8.42 | 0.0735 | 0.0125 | 2 |
| 4 | C10orf67 | singleton | 5.93 | 3.4043 | 0.7002 | 10 |
| 5 | LDLR | 0.001 | 24.30 | 0.2529 | 0.0245 | 19 |
| 5 | APOB | 0.001 | 9.15 | -0.2112 | 0.0343 | 2 |
| 5 | LPA | 0.1 | 7.14 | -0.0325 | 0.0060 | 6 |
| 5 | CDH26 | 0.001 | 6.97 | -0.1362 | 0.0256 | 20 |
| 5 | ABCG5 | 0.01 | 6.23 | 0.0850 | 0.0170 | 2 |
| 5 | HFE | 0.1 | 5.95 | -0.0218 | 0.0045 | 6 |
| 5 | CHEK2 | 0.01 | 5.69 | -0.0782 | 0.0165 | 22 |
| 5 | PDX1 | 0.01 | 5.62 | 0.1015 | 0.0215 | 13 |
| 6 | BRCA2 | 0.0001 | 5.95 | 0.1168 | 0.0240 | 13 |
| 8 | AFF1 | 0.0001 | 5.91 | -0.2546 | 0.0525 | 4 |
| 10 | MIP | 0.0001 | 6.46 | 0.3161 | 0.0620 | 12 |
| 10 | CASP7 | 0.01 | 5.91 | 0.0946 | 0.0195 | 10 |
| 16 | PKD1 | singleton | 6.68 | 0.3730 | 0.0719 | 16 |
| 16 | TET2 | 0.0001 | 6.10 | 0.2166 | 0.0439 | 4 |
| 19 | OCA2 | 0.01 | 10.35 | 0.0716 | 0.0109 | 15 |
| 19 | RAD52 | 0.1 | 6.29 | -0.0387 | 0.0077 | 12 |
| 20 | DEFB1 | 0.0001 | 6.05 | 0.7003 | 0.1425 | 8 |

**Table S18:** Significant rare variant associations: Mask6 (LoF, DelMis09, DelMis08, DelMis07, DelMis06, DelMis05 variants (most inclusive)). Gene-based rare variant association results for Mask6, showing the best MAF threshold per gene-signature pair. LOG10P = − log_10_(p-value). Genome-wide significance threshold: p *<* 2.5 × 10^−6^ (LOG10P ≥ 5.6).

| Sig | Gene | MAF | LOG10P | Beta | SE | Chr |
| --- | --- | --- | --- | --- | --- | --- |
| 0 | TTN | 0.0001 | 6.95 | 0.0491 | 0.0092 | 2 |
| 0 | RBM4 | singleton | 6.45 | 0.8613 | 0.1691 | 11 |
| 3 | ZNF577 | 0.001 | 5.65 | -0.2374 | 0.0502 | 19 |
| 5 | LDLR | 0.0001 | 20.63 | 0.2296 | 0.0242 | 19 |
| 5 | LPA | 0.1 | 7.13 | -0.0323 | 0.0060 | 6 |
| 5 | APOB | 0.0001 | 7.01 | -0.1653 | 0.0310 | 2 |
| 5 | CDH26 | 0.001 | 7.00 | -0.1348 | 0.0253 | 20 |
| 5 | ABCG5 | 0.01 | 5.86 | 0.0804 | 0.0167 | 2 |
| 9 | HCN2 | 0.0001 | 5.68 | -0.1916 | 0.0404 | 19 |
| 10 | MIP | 0.0001 | 6.05 | 0.2957 | 0.0602 | 12 |
| 10 | CASP7 | 0.01 | 5.95 | 0.0941 | 0.0193 | 10 |
| 16 | SRSF2 | 0.0001 | 7.09 | 1.0040 | 0.1871 | 17 |
| 16 | PKD1 | singleton | 6.43 | 0.3025 | 0.0595 | 16 |
| 18 | ZNF521 | singleton | 6.25 | 0.5693 | 0.1137 | 18 |
| 19 | OCA2 | 0.01 | 9.87 | 0.0676 | 0.0105 | 15 |
| 19 | RAD52 | 0.1 | 6.32 | -0.0388 | 0.0077 | 12 |

## Footnotes

1 See R3 Population Stratification Ancestry.ipynb.

2 See R3 AvoidingReverseCausation.ipynb.

## References and Notes

1. D. A. Berry, Bayesian clinical trials. Nature Reviews Drug Discovery 5 (1), 27–36 (2006), number: 1 Publisher: Nature Publishing Group, doi:10.1038/nrd1927, https://www.nature.com/articles/nrd1927.

2. A. Bellot, M. V. D. Schaar, Flexible Modelling of Longitudinal Medical Data: A Bayesian Nonparametric Approach. ACM Transactions on Computing for Healthcare 1 (1), 1–15 (2020), doi:10.1145/3377164, https://dl.acm.org/doi/10.1145/3377164.

3. D. C. Angus, C.-C. H. Chang, Heterogeneity of Treatment Effect: Estimating How the Effects of Interventions Vary Across Individuals. JAMA 326 (22), 2312–2313 (2021), doi:10.1001/jama.2021.20552, 10.1001/jama.2021.20552.

4. C. Sudlow, et al., UK Biobank: An Open Access Resource for Identifying the Causes of a Wide Range of Complex Diseases of Middle and Old Age. PLOS Medicine 12 (3), e1001779 (2015), doi: 10.1371/journal.pmed.1001779, https://dx.plos.org/10.1371/journal.pmed.1001779.

5. E. M. Pedersen, et al., ADuLT: An efficient and robust time-to-event GWAS. Nature Communications 14 (1), 5553 (2023), publisher: Nature Publishing Group, doi:10.1038/s41467-023-41210-z, https://www.nature.com/articles/s41467-023-41210-z.

6. S. M. Urbut, et al., MSGene: a multistate model using genetic risk and the electronic health record applied to lifetime risk of coronary artery disease. Nature Communications 15 (1), 4884 (2024), publisher: Nature Publishing Group, doi:10.1038/s41467-024-49296-9, https://www.nature.com/articles/s41467-024-49296-9.

7. J. Zhao, et al., Using topic modeling via non-negative matrix factorization to identify relationships between genetic variants and disease phenotypes: A case study of Lipoprotein(a) (LPA). PLOS ONE 14 (2), e0212112 (2019), publisher: Public Library of Science, doi:10.1371/journal.pone.0212112, https://journals.plos.org/plosone/article?id=10.1371/journal.pone.0212112.

8. X. Jiang, et al., Age-dependent topic modeling of comorbidities in UK Biobank identifies disease subtypes with differential genetic risk. Nature Genetics 55 (11), 1854–1865 (2023), doi: 10.1038/s41588-023-01522-8, https://www.nature.com/articles/s41588-023-01522-8.

9. S. M. Urbut, et al., Dynamic Importance of Genomic and Clinical Risk for Coronary Artery Disease Over the Life Course. medRxiv (2023), publisher: Cold Spring Harbor Laboratory Preprints.

10. V. Hyttinen, J. Kaprio, L. Kinnunen, M. Koskenvuo, J. Tuomilehto, Genetic liability of type 1 diabetes and the onset age among 22,650 young Finnish twin pairs: a nationwide follow-up study. Diabetes 52 (4), 1052–1055 (2003), doi:10.2337/diabetes.52.4.1052.

11. R. Caruana, Multitask Learning. *Machine Learning* **28** (1), 41–75 (1997), publisher: Springer Science and Business Media LLC, doi:10.1023/a:1007379606734, https://link.springer.com/10.1023/A:1007379606734.

12. C. E. Rasmussen, C. K. I. Williams, Gaussian Processes for Machine Learning (The MIT Press) (2006).

13. W. Wang, M. Stephens, Empirical Bayes Matrix Factorization. arXiv:1802.06931 [stat] (2021), arXiv: 1802.06931, http://arxiv.org/abs/1802.06931.

14. B. E. Engelhardt, M. Stephens, Analysis of Population Structure: A Unifying Framework and Novel Methods Based on Sparse Factor Analysis. PLoS Genet 6 (9), e1001117 (2010), doi:10.1371/journal.pgen.1001117, 10.1371/journal.pgen.1001117.

15. E. W. Steyerberg, Y. Vergouwe, Towards better clinical prediction models: seven steps for development and an ABCD for validation. European Heart Journal 35 (29), 1925–1931 (2014), doi:10.1093/eurheartj/ehu207, 10.1093/eurheartj/ehu207.

16. H. Putter, H. C. van Houwelingen, Understanding Landmarking and Its Relation with Time-Dependent Cox Regression. Stat Biosci 9 (2), 489–503 (2017), doi:10.1007/s12561-016-9157-9.

17. D. R. Cox, Regression Models and Life-Tables. Journal of the Royal Statistical Society. Series B (Methodological) 34 (2), 187–220 (1972), publisher: [Royal Statistical Society, Wiley], https://www.jstor.org/stable/2985181.

18. S. Koyama, et al., Decoding Genetics, Ancestry, and Geospatial Context for Precision Health. medRxiv (2023), publisher: Cold Spring Harbor Laboratory Preprints.

19. L. Bastarache, Using Phecodes for Research with the Electronic Health Record: From PheWAS to PheRS. Annual review of biomedical data science 4, 1–19 (2021), doi: 10.1146/annurev-biodatasci-122320-112352, https://www.ncbi.nlm.nih.gov/pmc/articles/PMC9307256/.

20. G. Hripcsak, D. J. Albers, Next-generation phenotyping of electronic health records. Journal of the American Medical Informatics Association 20 (1), 117–121 (2013), publisher: Oxford University Press (OUP), doi:10.1136/amiajnl-2012-001145, https://academic.oup.com/jamia/article-lookup/doi/10.1136/amiajnl-2012-001145.

21. M. W. Yeung, P. Van Der Harst, N. Verweij, ukbpheno v1.0: An R package for phenotyping health-related outcomes in the UK Biobank. STAR Protocols 3 (3), 101471 (2022), doi:10.1016/j.xpro.2022.101471, https://linkinghub.elsevier.com/retrieve/pii/S2666166722003513.

22. J. Cohen, Statistical Power Analysis for the Behavioral Sciences (Routledge), 0 ed. (2013), doi:10.4324/9780203771587, https://www.taylorfrancis.com/books/9781134742707.

23. D. J. Thompson, et al., UK Biobank release and systematic evaluation of optimised polygenic risk scores for 53 diseases and quantitative traits (2022), doi:10.1101/2022.06.16.22276246, https://www.medrxiv.org/content/10.1101/2022.06.16.22276246v2, iSSN: 2227-6246 Pages: 2022.06.16.22276246.

24. B. K. Bulik-Sullivan, et al., LD Score regression distinguishes confounding from polygenicity in genomewide association studies. Nature Genetics 47 (3), 291–295 (2015), doi:10.1038/ng.3211, http://www.nature.com.proxy.uchicago.edu/ng/journal/v47/n3/full/ng.3211.html.

25. C. J. Willer, et al., Discovery and refinement of loci associated with lipid levels. Nature genetics 45 (11), 1274–1283 (2013).

26. S. Kathiresan, D. Srivastava, Genetics of human cardiovascular disease. Cell 148 (6), 1242–1257 (2012).

27. S. F. Grant, et al., Variant of transcription factor 7-like 2 (TCF7L2) gene confers risk of type 2 diabetes. Nature genetics 38 (3), 320–323 (2006).

28. D. M. Evans, et al., Meta-analysis of genome-wide association studies confirms a susceptibility locus for knee osteoarthritis on chromosome 7q22. Annals of the rheumatic diseases 70 (2), 349–355 (2011).

29. C. Tcheandjieu, et al., Large-scale genome-wide association study of coronary artery disease in genetically diverse populations. Nature Medicine 28 (8), 1679–1692 (2022), publisher: Nature Publishing Group, doi:10.1038/s41591-022-01891-3, https://www.nature.com/articles/s41591-022-01891-3.

30. M. Abifadel, et al., Mutations in PCSK9 cause autosomal dominant hypercholesterolemia. Nature genetics 34 (2), 154–156 (2003).

31. M. Benn, A. Tybjærg-Hansen, S. Stender, R. Frikke-Schmidt, B. G. Nordestgaard, Genetic and environmental influences on premature death in adult adoptees. New England Journal of Medicine 358 (13), 1360–1368 (2008).

32. D. S. Herman, et al., Truncations of titin causing dilated cardiomyopathy. New England Journal of Medicine 366 (7), 619–628 (2012).

33. S. Jaiswal, et al., Age-related clonal hematopoiesis associated with adverse outcomes. New England Journal of Medicine 371 (25), 2488–2498 (2014).

34. P. C. Harris, V. E. Torres, Autosomal dominant polycystic kidney disease: the last 3 years. Kidney international 76 (2), 149–168 (2009).

35. A. Shiels, S. Bassnett, Mutations in the founder of the MIP gene family underlie cataract development in the mouse. Nature Genetics 12 (2), 212–215 (1996), doi:10.1038/ng0296-212.

36. B. G. Nordestgaard, et al., Familial hypercholesterolaemia is underdiagnosed and undertreated in the general population: guidance for clinicians to prevent coronary heart disease: consensus statement of the European Atherosclerosis Society. European heart journal 34 (45), 3478–3490 (2013).

37. S. Jaiswal, et al., Clonal hematopoiesis and risk of atherosclerotic cardiovascular disease. New England Journal of Medicine 377 (2), 111–121 (2017).

38. T. Schoeler, J.-B. Pingault, Z. Kutalik, The impact of self-report inaccuracy in the UK Biobank and its interplay with selective participation. Nature Human Behaviour 9 (3), 584–594 (2025), epub 2024 Dec 18, doi:10.1038/s41562-024-02061-w.

39. S. van Alten, B. W. Domingue, J. Faul, T. Galama, A. T. Marees, Reweighting UK Biobank corrects for pervasive selection bias due to volunteering. International Journal of Epidemiology 53 (3), dyae054 (2024), doi:10.1093/ije/dyae054, 10.1093/ije/dyae054.

40. M. H. Gail, et al., Projecting Individualized Probabilities of Developing Breast Cancer for White Females Who Are Being Examined Annually. JNCI: Journal of the National Cancer Institute 81 (24), 1879–1886 (1989), doi:10.1093/jnci/81.24.1879, 10.1093/jnci/81.24.1879.

41. E. A. Ashley, Towards precision medicine. *Nature Reviews Genetics* **17** (9), 507–522 (2016), publisher: Springer Science and Business Media LLC, doi:10.1038/nrg.2016.86, https://www.nature.com/articles/nrg.2016.86.

42. F. S. Collins, H. Varmus, A New Initiative on Precision Medicine. New England Journal of Medicine 372 (9), 793–795 (2015), publisher: Massachusetts Medical Society, doi:10.1056/nejmp1500523, http://www.nejm.org/doi/10.1056/NEJMp1500523.

43. M. Martínez-Garćıa, E. Hernández-Lemus, Data integration challenges for machine learning in precision medicine. Frontiers in medicine 8, 784455 (2022).

44. N. J. Schork, Personalized medicine: Time for one-person trials. Nature 520 (7549), 609–611 (2015), publisher: Springer Science and Business Media LLC, doi:10.1038/520609a, https://www.nature.com/articles/520609a.

45. A. L. Price, C. C. A. Spencer, P. Donnelly, Progress and promise in understanding the genetic basis of common diseases. Proceedings of the Royal Society B: Biological Sciences 282 (1821), 20151684 (2015), publisher: The Royal Society, doi:10.1098/rspb.2015.1684, https://royalsocietypublishing.org/doi/10.1098/rspb.2015.1684.

46. M. J. Joyner, N. Paneth, Promises, promises, and precision medicine. Journal of Clinical Investigation 129 (3), 946–948 (2019), publisher: American Society for Clinical Investigation, doi:10.1172/jci126119, https://www.jci.org/articles/view/126119.

47. S. R. Steinhubl, E. J. Topol, Digital medicine, on its way to being just plain medicine. npj Digital Medicine 1 (1) (2018), publisher: Springer Science and Business Media LLC, doi:10.1038/s41746-017-0005-1, https://www.nature.com/articles/s41746-017-0005-1.

48. J. Tyler, S. W. Choi, M. Tewari, Real-time, personalized medicine through wearable sensors and dynamic predictive modeling: A new paradigm for clinical medicine. Current Opinion in Systems Biology 20, 17–25 (2020), 10.1016/j.coisb.2020.07.001, https://www.sciencedirect.com/science/article/pii/S2452310020300068.

49. N. Simon, R. Simon, Adaptive enrichment designs for clinical trials. Biostatistics 14 (4), 613–625 (2013), publisher: Oxford University Press (OUP), doi:10.1093/biostatistics/kxt010, https://academic.oup.com/biostatistics/article-lookup/doi/10.1093/biostatistics/kxt010.

50. A. Shmatko, et al., Learning the natural history of human disease with generative transformers. Nature 647 (8088), 248–256 (2025), doi:10.1038/s41586-025-09529-3, 10.1038/ s41586-025-09529-3.

51. J. Amar, et al., Integrating Genomics into Multimodal EHR Foundation Models (2025), https://arxiv.org/abs/2510.23639.

52. Z. D. Bailey, et al., Structural racism and health inequities in the USA: evidence and interventions. The Lancet 389 (10077), 1453–1463 (2017), publisher: Elsevier BV, doi:10.1016/s0140-6736(17)30569-x, https://linkinghub.elsevier.com/retrieve/pii/S014067361730569X.

53. L. L. Guo, et al., A multi-center study on the adaptability of a shared foundation model for electronic health records. NPJ digital medicine 7 (1), 171 (2024).

54. J. D. Kalbfleisch, R. L. Prentice, The Statistical Analysis of Failure Time Data (John Wiley & Sons) (2011).

55. S. M. Urbut, et al., MSGene: Derivation and validation of a multistate model for lifetime risk of coronary artery disease using genetic risk and the electronic health record. medRxiv (2023), publisher: Cold Spring Harbor Laboratory Preprints.

56. M. W. Yeung, N. VERWEIJ, niekverw/ukbpheno: v1.0.0 (2022), doi:10.5281/ZENODO.6557829, https://zenodo.org/record/6557829.

57. C. Bycroft, et al., The UK Biobank resource with deep phenotyping and genomic data. Nature 562 (7726), 203–209 (2018), doi:10.1038/s41586-018-0579-z, https://www.nature.com/articles/s41586-018-0579-z.

58. S. Urbut, et al., MS Gene: Multistate Modeling of Dynamic Lifetime Risk of Coronary Artery Disease Using Electronic Health Records in the UK Biobank. Circulation 148 (Suppl 1), A14747–A14747 (2023), publisher: Lippincott Williams & Wilkins Hagerstown, MD.

59. The All of Us Research Program Investigators, The “All of Us” Research Program 381 (7), 668–676, doi:10.1056/NEJMsr1809937, http://www.nejm.org/doi/10.1056/NEJMsr1809937.

60. J. Mbatchou, et al., Computationally efficient whole-genome regression for quantitative and binary traits. Nature Genetics 53 (7), 1097–1103 (2021), doi:10.1038/s41588-021-00870-7, 10.1038/s41588-021-00870-7.

61. S. S. Khan, et al., Development and Validation of the American Heart Association’s PREVENT Equations 149 (6), 430–449, eprint: https://www.ahajournals.org/doi/pdf/10.1161/CIRCULATIONAHA.123.067626, doi:10.1161/CIRCULATIONAHA.123.067626, https://www.ahajournals.org/doi/abs/10.1161/CIRCULATIONAHA.123.067626.

62. J. D. M. Lloyd, et al., Estimating Longitudinal Risks and Benefits From Cardiovascular Preventive Therapies Among Medicare Patients. Journal of the American College of Cardiology 69 (12), 1617– 1636 (2017), publisher: American College of Cardiology Foundation, doi:10.1016/j.jacc.2016.10.018, https://www.jacc.org/doi/10.1016/j.jacc.2016.10.018.

63. M. Ambrosio, et al., Performance of PREVENT and pooled cohort equations for predicting 10-Year ASCVD risk in the UK Biobank. American Journal of Preventive Cardiology 22, 101009 (2025), publisher: Elsevier BV, doi:10.1016/j.ajpc.2025.101009, https://linkinghub.elsevier.com/retrieve/pii/S2666667725000844.

